# A cost consequence analysis of six diagnostic strategies for ovarian cancer: A model-based economic evaluation

**DOI:** 10.1101/2025.10.17.25338224

**Authors:** Samuel J. Perry, Donal Griffin, Eleanor V. Williams, Tracy E. Roberts, Fong Lien Kwong, Sarah Williams, Jon Deeks, Katie Scandrett, Ridhi Agarwal, Sudha S. Sundar, Mark Monahan

**Affiliations:** Department of Applied Health Sciences, College of Medicine and Health, University of Birmingham.; Pan Birmingham Gynaecological Cancer Centre, Midlands Metropolitan University Hospital, Birmingham, UK; Department of Cancer and Genomic Sciences, University of Birmingham, Birmingham, UK; University Hospitals Birmingham, UK; NIHR Birmingham Biomedical Research Centre, University Hospitals Birmingham NHS Foundation Trust, University of Birmingham, Birmingham, UK

**Keywords:** Gynaecologic cancer, economic evaluation, ultrasound, cost-effectiveness, menopause

## Abstract

**Objective:** To assess the costs and consequences of six diagnostic strategies for ovarian cancer in pre-menopausal and post-menopausal women with symptoms in secondary care.

**Design:** Economic evaluation alongside a prospective single-arm diagnostic accuracy study.

**Setting:** NHS secondary care outpatients (2-week referrals, clinics, GP referrals, cross-specialty referrals) and inpatients (emergency presentations to secondary care).

**Sample:** Two cohorts of 857 pre-menopausal and 1,242 post-menopausal newly presenting to secondary care with symptoms of suspected ovarian cancer.

**Methods:** A model-based cost-consequence analysis (CCA) was conducted using a decision tree simulating patient pathways over 12-months. Diagnostic accuracy data were sourced from the ROCkeTS study and supplemented by literature.

**Main outcome measures:** Cancer deaths, correct diagnosis proportion, and diagnostic yield.

**Results:** No diagnostic strategy was optimal across all outcomes. Across both cohorts, the Risk of Malignancy Index (RMI) 200 was least expensive but had poor cancer death and diagnostic yield outcomes. The ADNEX 3% strategy had the highest diagnostic yield and lowest cancer mortality but was the most expensive.

For pre-menopausal women, the IOTA ADNEX 10% strategy outperformed ORADS, ROMA, and CA125 in cost and outcomes. For post-menopausal women, the high cancer prevalence required a trade-off. In sensitivity analysis a two-step IOTA ADNEX 10% strategy outperformed ORADS, ROMA, and CA125 across all three outcomes, making the strategy a more balanced choice in both cohorts.

**Conclusion:** At 12 months, no single diagnostic strategy was superior. Early diagnosis requires balancing cancer mortality, diagnostic yield, and cost. The IOTA ADNEX two-step strategy at 10% threshold provided the best trade-off across these factors and is recommended for practice.

**Funding:** This study is funded by a grant from National Institute of Heath Research, Health Technology assessment HTA 13/13/01.

## Introduction

Ovarian cancer remains a significant health concern worldwide, with over 300,000 new cases diagnosed annually [1]. In the UK, approximately 7,500 new cases are diagnosed each year [2], with around 70% diagnosed at advanced stages [3] necessitating extensive treatment including maximum cytoreductive surgery, chemotherapy, and targeted therapy [4]. Due to the high proportion of late-stage diagnoses, ovarian cancer has a substantial mortality burden, with approximately 4,100 deaths occurring annually in the UK [2]. Whilst most cases occur post-menopause, some pre-menopausal women are diagnosed each year [5].

Women presenting to general practitioners in the UK are tested with cancer antigen 125 (CA125) and ultrasound and referred to secondary care if these are abnormal[4]. However, adherence to this guidance varies in clinical practice [7]. In hospital, women receive a triage test to assess their risk of ovarian cancer. Professional societies endorse different risk prediction models for ovarian cancer. The Risk of Malignancy Index (RMI) is the UK standard of care [4], whilst IOTA ADNEX [8], ORADS [9] and Risk of ovarian malignancy algorithm (ROMA) [10] are used internationally. Effective triage is critical to ensure high-risk patients receive appropriate surgery with the best outcomes [11].

Current UK guidance endorses the RMI 200 strategy, which has 70% sensitivity and 90% specificity [12,13]. To compensate for poor sensitivity, surgery is recommended irrespective of diagnostic outcome: women with RMI≥200 undergo surgery at a designated gynaecological cancer centre by a specialist gynaecological surgical oncologist, whereas women with RMI<200 undergo surgery at a cancer unit by a gynaecologist. This disadvantages women with early-stage cancer who undergo surgery by non-cancer specialists and those without cancer who may have avoided unnecessary surgery with a more accurate test. The high rate of surgical intervention under the current pathway (70% in the ROCkeTS study) demonstrates the need for refined tools to improve cancer detection while minimising unnecessary surgeries and associated morbidity.

The Refining Ovarian Cancer Test Accuracy Scores (ROCkeTS) study externally validated risk prediction models estimating the probability of ovarian cancer for women with non-specific symptoms in pre- and post-menopausal cohorts [6]. ROCkeTS was a prospective single-arm diagnostic accuracy study where all patients received all commonly used tests and diagnostic accuracy was evaluated against gold standard histology or 12-month outcome [6]. The test accuracy results are published elsewhere [14]. Here, we report the economic evaluation conducted alongside ROCkeTs, assessing the costs and consequences of six different ovarian cancer diagnostic testing strategies in two cohorts (pre- and post-menopausal women) using ROCkeTS data.

## Methods

### Study design of ROCkeTS prospective study

Participants were eligible for inclusion in the ROCkeTS study if they were aged 16-90 years and newly presenting to secondary care with symptoms and either an abnormal CA125 test, abnormal imaging, or both. Participants donated a sample of blood and underwent a transvaginal and transabdominal ultrasound performed mainly by NHS sonographers with certified training in the use of IOTA ultrasound models [6]. The study recruited women at ‘low’ or unknown genetic risk of ovarian cancer, i.e., women not known to have a BRCA or other germline mutation predisposing to ovarian cancer. Menopause was defined as more than 12 months without menstruation. Women who had not menstruated for more than 12 months due to contraception or hysterectomy were categorised by age: those under 50 as pre-menopausal and those 51 and over as post-menopausal. Women who were still menstruating and over 50 years old were considered pre-menopausal. The ovarian cancer definition used in this study is primary invasive ovarian malignant neoplasm or diagnosed with ovarian cancer in the last 12 months.

### Current standard of care diagnostic pathway

When a patient presents with symptoms of ovarian cancer in secondary care, usual practice involves a series of diagnostic tests and procedures. The RMI I score is calculated which combines CA125, limited ultrasound variables, and menopausal status. Women with an RMI I score ≥200 are referred to a cancer specialist multidisciplinary team and a CT scan of the pelvis and abdomen is also performed to establish the extent of disease [4].

Women with CT scan findings of disseminated cancer in the abdomen undergo image guided biopsy to confirm malignancy, others will undergo surgery[12,13]. Patients with RMI 25-200 undergo surgery in a cancer unit and may also undergo an MRI scan. Those considered at low risk of cancer (RMI<25) will be managed under surveillance with follow-up scans [13].

### Cost consequence modelling methods

A model-based cost-consequence analysis (CCA) was undertaken using a decision tree developed in TreeAge Pro 2024 [15]. A UK National Health Service (NHS) perspective was adopted [16]. The model structure and methods followed the Consolidated Health Economic Evaluation Reporting Standards (CHEERS) checklist [17] (Supporting Information Table S6). The modelled diagnostic strategies considered where a patient was identified as having ovarian cancer were as follows:

- RMI I test score ≥ 200

- RMI is a scoring system used to estimate the likelihood that an ovarian mass is malignant. A score ≥200 indicates a high risk of ovarian malignancy [18].
- ROMA test result of manufacturer recommended thresholds (>29.9% for post-menopausal cohort; >11.4% for pre-menopausal cohort).

- ROMA uses blood markers CA125 and HE4 (human epididymis protein 4) along with menopausal status to estimate cancer risk. Results above the thresholds indicate a high risk of epithelial ovarian cancer [10].
- IOTA ADNEX test result

- IOTA ADNEX model is used to differentiate between benign and malignant ovarian tumours [19].The model uses six ultrasound variables and three clinical parameters (age, optional CA125 level and setting) [20]. Two different thresholds are considered, at 10% and at 3%.
- CA125 test result ≥ 35 u/ml

- CA125 is a blood test measuring a protein linked to ovarian cancer. A level above 35 u/ml is considered elevated [21].
- Ovarian-ADNEXal Reporting and Data System (ORADS) test result ≥4

- ORADS is a standardised imaging classification for ovarian masses based on pattern recognition in ultrasound. A score ≥4 means there is a moderate-to-high suspicion of malignancy (>10%). [9]

The outcomes considered in the CCA were cancer death, proportion of correct diagnosis and diagnostic yield. A correct diagnosis was those who were correctly identified as having ovarian cancer (true positive) or correctly ruled out (true negative). An incorrect diagnosis was those incorrectly identified as having ovarian cancer (false positive) or incorrectly ruled out (false negative). Diagnostic yield is the percentage of cancers detected out of the population screened. The pathways were the same for both cohorts and differed only by the test characteristics and underlying ovarian cancer prevalence in each population. Model inputs are listed in Tables S1-S3.

A 12-month end point was used as the time horizon for this model. Discounting was not undertaken due to the time horizon length. A cost year of 2022-2023 was used for the model. Costs were inflated where applicable using the PSSRU inflation index [22].

It was decided a CCA best represented the different outcome measures and trade-offs associated with diagnosis. When considering alternative approaches such as a cost-effectiveness analysis we found no one outcome measure would sufficiently represent patients with and without ovarian cancer. Furthermore, the decision against cost-utility analysis (CUA) was due to ROCkeTS not measuring health related quality of life (HRQoL) data as this was a diagnostic test accuracy study. We also found insufficient utility data available from previous studies and databases to accurately reflect all outcomes when using a comprehensive patient pathway mapping approach.

### Model Assumptions

Several simplifying assumptions were required to develop a workable model given data limitations and variations in care recommendations. These were agreed upon by the clinical research team prior to conducting the analysis.

All ovarian cancer patients were assumed to receive treatment, meaning best supportive care was not an option. All patients were assumed to respond to first-line platinum-based chemotherapy, therefore second-line treatment was not considered within the 12-month timeline. First-line treatment was assumed to take 6 months from test result, and maintenance therapy (where applicable) was for the remaining 6 months. All those receiving cancer treatment were assumed to undergo either germline BRCA testing (FIGO Stage I) or MyChoice homologous recombination deficiency (HRD) testing (FIGO Stage II-IV). No distinction was made between those who are BRCA germline mutation carriers and HRD groups. Treatment or tumour testing was not differentiated by ovarian cancer histology types. All chemotherapy patients had a computed tomography (CT) scan before the first cycle, and after the third and sixth cycles. All follow-ups up to 12 months consisted of CA125 tests and CT scans with a gynaecologist in secondary care. Stage-specific cancer deaths were taken from NHS digital and comprise the 12-month age standardised cancer deaths of adult patient cancers diagnosed 2016 to 2020 in England [23].

Key assumptions are provided in Table 1 for the TP, FN, FP, and FN patient pathways, with further details provided in Supporting Information (p.18-21)

**Table 1.**
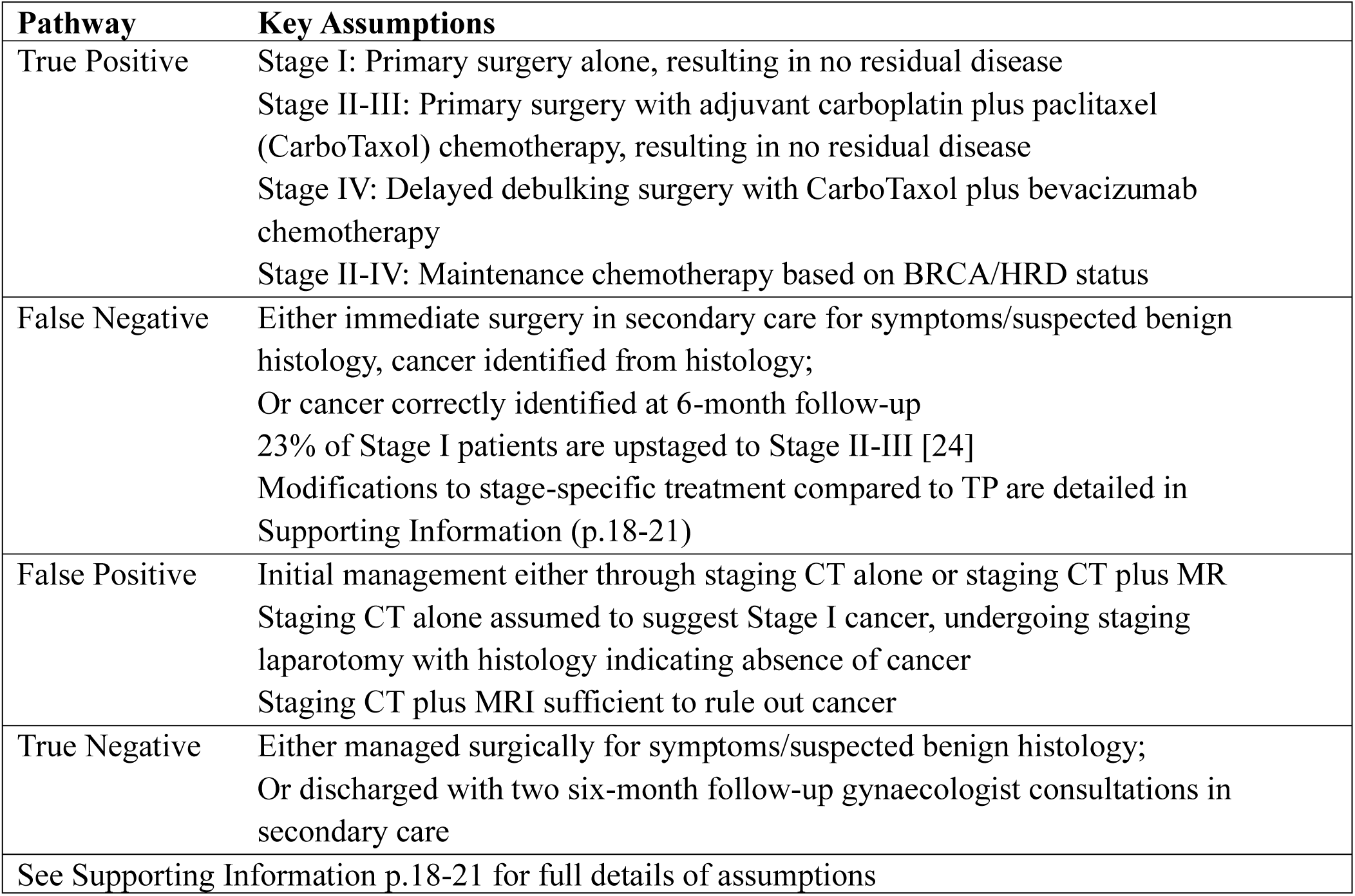
Key Test Result Patient Pathway Assumptions thway Key Assumptions.

**Table 2.** Base case deterministic results.

| Strategy | Cost (£) | Cancer deaths (%) | Proportion of correct diagnosis (%) | Diagnostic yield (%) |
| --- | --- | --- | --- | --- |
| **Pre** |  |  |  |  |
| RMI 200 | 5,283 | 1.17 | 92.51 | 3.00 |
| ADNEX<br>10% | 6,823 | 1.06 | 75.99 | 5.47 |
| ORADS | 6,964 | 1.12 | 84.59 | 4.86 |
| CA125 | 7,609 | 1.43 | 65.15 | 4.76 |
| ADNEX 3% | 7,795 | 1.05 | 48.47 | 5.73 |
| ROMA<br>11.4% | 8,056 | 1.13 | 73.45 | 4.86 |
| **Post** |  |  |  |  |
| RMI 200 | 13,471 | 3.84 | 84.52 | 16.94 |
| ORADS | 14,243 | 4.02 | 77.92 | 15.22 |
| CA125 | 14,414 | 4.04 | 79.27 | 17.14 |
| ADNEX<br>10% | 14,471 | 3.73 | 65.99 | 19.15 |
| ROMA<br>29.9% | 14,494 | 3.96 | 81.51 | 17.53 |
| ADNEX 3% | 15,119 | 3.70 | 44.59 | 19.93 |

### Analysis

The analysis was stratified by the pre- and post-menopausal cohorts due to a higher ovarian cancer prevalence amongst post-menopausal women, and the potential adverse fertility impact of surgical intervention on pre-menopausal women [5]. Strategies were ranked by costs, and an incremental approach was taken whereby strategies were compared against the nearest alternative in cost. A strategy was considered dominated if it was more expensive and less effective on all outcome measures compared to a strategy.

### Deterministic sensitivity analysis

The following deterministic sensitivity analyses (DSAs) were conducted to assess the impact on the base case results for both cohorts:

1. The prevalence of ovarian cancer was varied ± 25%.
2. The lower and upper confidence intervals of the upstaging variable from Van de Vorst (2021) [24]
3. The proportion of FPs that have additional MRI was varied ± 25%.
4. Proportion of TPs with Stage I Ovarian Cancer was varied ± 25%.
5. Proportion of correctly staged resectable patients who have all listed surgery procedures was varied ± 25%.

Additionally, the ROCkeTS study assessed the performance of ultrasound models when used by NHS sonographers with varying levels of expertise, ensuring they had received appropriate training, certification and quality assurance. To objectively evaluate these models, the ROCkeTS diagnostic accuracy analysis excluded subjective assessments of ovarian masses during ultrasound examinations. This approach isolated the performance of ultrasound-based models and minimised bias linked to individual operator expertise. In clinical practice, an alternative approach could triage out approximately 36% of women with benign-appearing tumours during the same ultrasound examination using a two-step strategy [25]. To account for this, we conducted a sixth DSA:

6. For the ADNEX 10% strategy, the sensitivity and specificity of a two-step strategy was used [25] with an absolute reduction of 17% in surgeries for FPs [26]

### Probabilistic Sensitivity Analysis

Probabilistic sensitivity analysis (PSA) was conducted to explore model input uncertainty. The distributions and parameters used are shown in Tables S2-3. Beta distributions were attached to the test proportions and count data. No distribution was attached where there were zero count data. Gamma distributions were attached to cost data and, with a 20% standard error assumed where information was unavailable. 10,000 iterations were run with a seeded random generator. Mean costs and outcomes were calculated across the iterations along with their 95% prediction intervals. Incremental cost-consequences were presented to illustrate parameter uncertainty with separate scatterplots for each outcome measure A 95% confidence ellipse is also presented within the incremental cost-consequence scatterplot to convey the region where 95% of the incremental iterations resided [27].

## Results

The base case deterministic results for the pre-menopausal cohort (N=857) and post-menopausal cohorts (N=1,242) are shown in Table 1. In the pre-menopausal cohort, 5.7% were diagnosed with ovarian cancer, 87.5% had no ovarian cancer, and 6.8% fell into other categories. In the post-menopausal cohort 17.3% were diagnosed with ovarian cancer, 69.3% had no ovarian cancer and 13.4% were classified as other.

For both cohorts, the RMI 200 strategy was the least costly and had the highest proportion of correct diagnosis but performed poorly on diagnostic yield relative to other strategies. The ADNEX 3% strategy was associated with the lowest number of cancer deaths and highest diagnostic yield in both cohorts. For the pre-menopausal cohort, the ORADS, ROMA 11.4% and CA125 strategies were dominated by the ADNEX 10% strategy. Given the high prevalence of ovarian cancer in post-menopausal women, selecting a diagnostic strategy involves a trade-off. Costs were higher in the post-menopausal cohort compared to the pre-menopausal cohort, reflecting the increased prevalence of the disease.

The DSA results are shown in Tables S4-5. Decreasing the ovarian cancer prevalence improved the proportion of correct diagnosis for strategies with higher specificity such as RMI 200 while increasing ovarian cancer prevalence improved the proportion of correct diagnosis for strategies with higher sensitivity such as ADNEX 3%. Changing the ovarian cancer prevalence had the greatest impact on the expected cost per strategy, although the order of strategies by cost remained consistent with the base case analysis.

For the two-step ADNEX 10% strategy the expected costs and diagnostic yield decreased while cancer deaths and proportion of correct diagnosis increased relative to baseline in both cohorts. In the post-menopausal cohort, this reduction in cost meant the two-step ADNEX 10% category now had the second lowest cost after RMI 200, whilst it retained this ordering from base case in the pre-menopausal cohort. As such, for both cohorts the ORADS, ROMA, and CA125 strategies were dominated by the two-step strategy. Furthermore, the two-step strategy continued to outperform RMI 200 on cancer deaths and diagnostic yield, whilst the disparity in proportion of correct diagnosis narrowed.

The mean PSA iterations for the two cohorts are shown in Tables 3–4 with associated 95% prediction intervals for the costs and consequences. The average costs and outcome measure effectiveness were similar to the deterministic base case results.

**Table 3.**
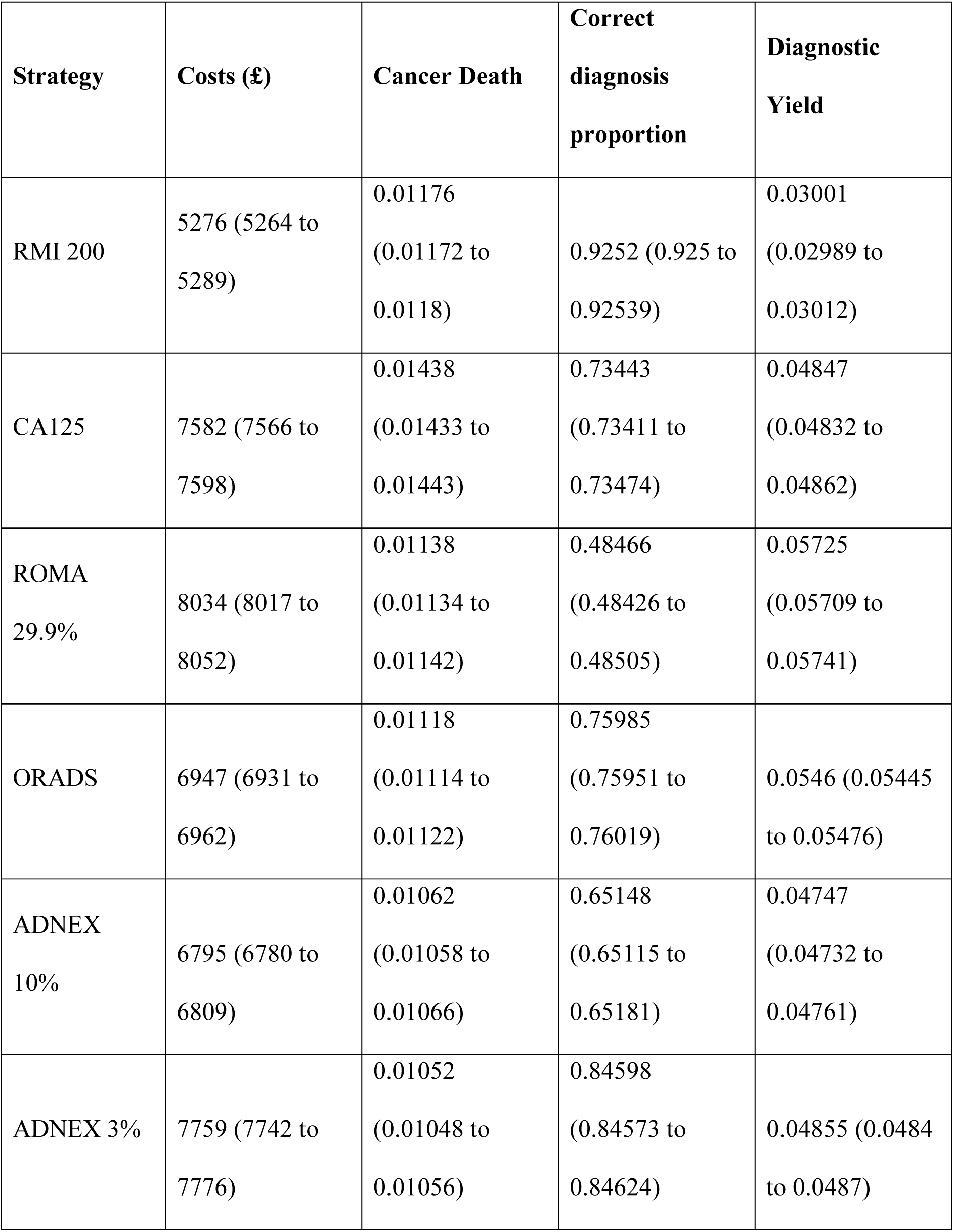
PSA mean output premenopausal cohort.

| **Strategy** | **Costs (£)** | **Cancer Death** | **Correct**<br>**diagnosis**<br>**proportion** | **Diagnostic**<br>**Yield** |
| --- | --- | --- | --- | --- |
| RMI 200 | 5276 (5264 to 5289) | 0.01176<br>(0.01172 to 0.0118) | 0.9252 (0.925 to 0.92539) | 0.03001<br>(0.02989 to 0.03012) |
| CA125 | 7582 (7566 to 7598) | 0.01438<br>(0.01433 to 0.01443) | 0.73443<br>(0.73411 to 0.73474) | 0.04847<br>(0.04832 to 0.04862) |
| ROMA<br>29.9% | 8034 (8017 to 8052) | 0.01138<br>(0.01134 to 0.01142) | 0.48466<br>(0.48426 to 0.48505) | 0.05725<br>(0.05709 to 0.05741) |
| ORADS | 6947 (6931 to 6962) | 0.01118<br>(0.01114 to 0.01122) | 0.75985<br>(0.75951 to 0.76019) | 0.0546 (0.05445 to 0.05476) |
| ADNEX<br>10% | 6795 (6780 to 6809) | 0.01062<br>(0.01058 to 0.01066) | 0.65148<br>(0.65115 to 0.65181) | 0.04747<br>(0.04732 to 0.04761) |
| ADNEX 3% | 7759 (7742 to 7776) | 0.01052<br>(0.01048 to 0.01056) | 0.84598<br>(0.84573 to 0.84624) | 0.04855 (0.0484 to 0.0487) |

**Table 4.** PSA mean output postmenopausal cohort.

| Strategy | Costs | Cancer Death | Correct diagnosis proportion | Diagnostic Yield |
| --- | --- | --- | --- | --- |
| RMI 200 | 13460 (13436 to 13485) | 0.03842<br>(0.03836 to 0.03848) | 0.84484<br>(0.84461 to 0.84507) | 0.16924<br>(0.16902 to 0.16947) |
| CA125 | 14417 (14391 to 14442) | 0.04034<br>(0.04028 to 0.04041) | 0.81502<br>(0.81477 to 0.81526) | 0.17512<br>(0.17489 to 0.17535) |
| ROMA 29.9% | 14482 (14456 to 14507) | 0.03957<br>(0.03951 to 0.03964) | 0.44622<br>(0.44591 to 0.44654) | 0.19908<br>(0.19884 to 0.19932) |
| ORADS | 14237 (14212 to 14262) | 0.04014<br>(0.04008 to 0.0402) | 0.65945<br>(0.65913 to 0.65976) | 0.19136<br>(0.19112 to 0.1916) |
| ADNEX 10% | 14478 (14452 to 14503) | 0.03728<br>(0.03722 to 0.03734) | 0.79244 (0.7922 to 0.79268) | 0.17127<br>(0.17104 to 0.17149) |
| ADNEX 3% | 15113 (15085 to 15140) | 0.03696 (0.0369 to 0.03702) | 0.7789 (0.77865 to 0.77915) | 0.15221<br>(0.15199 to 0.15243) |

The incremental cost-consequence scatterplots for the three outcome measures for the pre-menopausal cohort and post-menopausal cohort are shown in Figures S8-22 and Figures S23-37, respectively. All scatterplots used RMI 200 as a baseline comparator.

For the pre-menopausal cohort, the RMI 200 strategy was less expensive in the majority of the iterations relative to the other comparators. ROMA 11.4% was consistently more expensive than RMI 200 across iterations. For cancer deaths, the PSA output indicates an inverse relationship between cost and deaths for the incremental iterations. For correct diagnosis proportion, no strategy had a better correct diagnosis proportion than the RMI 200 strategy across the PSA iterations. Furthermore, for the diagnostic yield, all the other strategies performed better on diagnostic yield compared to RMI 200.

For the post-menopausal cohort, the RMI 200 strategy was generally less expensive in the majority of the iterations relative to the other comparators. While the inverse relationship between cost and deaths was seen again in the post-menopausal cohort, the cancer death incremental iterations tended to be less dispersed compared to the pre-menopausal cohort ones. For correct diagnosis proportion, RMI 200 had a higher correct diagnosis proportion than ORADS, ADNEX 10% and ADNEX 3%. The RMI 200 strategy had a much higher correct diagnosis proportion than ROMA 29.9% and CA125 strategies. Furthermore, as with the pre-menopausal cohort, all the other strategies performed better on diagnostic yield compared to the RMI 200 strategy.

## Discussion

### Principal Findings

This study assessed the costs and consequences of six diagnostic strategies in ovarian cancer screening. Decision tree modelling comprehensively mapped the patient pathway for pre- and post-menopausal women with suspected ovarian cancer, capturing key costs and clinical consequences during the first 12 months after diagnostic testing. No single strategy outperformed all others across all outcome measures: cost, diagnostic yield, cancer deaths and proportion correctly diagnosed. While RMI 200 was the cheapest in both cohorts, its lower sensitivity resulted in more cancer-related deaths and a lower diagnostic yield compared to ADNEX-based strategies. Sensitivity analysis supported the base-case findings across both cohorts.

The two ADNEX-based strategies modelled in the analysis using ROCkeTS data (where the risk prediction models were tested in isolation) were characterised by high sensitivity but lower specificity, resulting in potentially unnecessary costs associated with the management of false positive results. Recently, Landolfo et al. [25] evaluated a two-step ultrasound IOTA ADNEX strategy in 4,905 patients, first applying benign descriptors for initial triage, followed by the IOTA ADNEX model. At a 10% risk threshold, they reported a sensitivity of 91% and a specificity of 85%. In a sensitivity analysis, this two-step strategy with IOTA ADNEX 10% was modelled alongside a reduction in surgery rate of 17%, extrapolated from Nunes et al. [26] findings for ultrasound-based models. This reduced false positives and presented the best balance across all outcome measures, making it suitable for clinical implementation. However, a two-step strategy also slightly underestimated the risk of malignancy compared to a strategy of IOTA ADNEX alone.

### Strengths and Limitations

A key strength of this analysis its use of prospectively collected data from the ROCkeTS study to facilitate the comparisons across numerous diagnostic strategies. ROCkeTS is the first head-to head comparison prospective study published investigating all currently used diagnostic models and tests. Additionally, the analysis stratified pre- and post-menopausal cohorts to reflect key differences in prevalence of cancer, performance of diagnostic models, histology types and consequences of treatment in both groups [28–30]. Another key strength is the incorporation of up-to-date treatment modalities, including germline testing for inherited cancer, tumour testing for HRD, and appropriate treatment with targeted therapies such as Bevacizumab and PARP inhibitors. Furthermore, the present analysis is also the first to incorporate upstaging within the model and incorporate the impact of different diagnostic strategies on surgery rates.

This study is not without limitations. Although it employed a 12-month time horizon to comprehensively examine how a patient would transition through treatment pathways and their associated costs within the first year from diagnosis, a longer time horizon may have given a better reflection of important differences in costs and outcomes between the alternative diagnostic strategies. For instance, differences in cancer deaths would likely increase over time between strategies with varying levels of sensitivity. Additionally, within the 12-month horizon considered, costs associated with a strategy improving early accurate ovarian cancer diagnosis were higher due to longer durations of maintenance therapies.

A further limitation was the inability to model the impacts of ovarian cancer on fertility and related complications in pre-menopausal women. Ovarian cancer treatment in pre-menopausal women frequently results in infertility due to the aggressive nature of the surgical procedures and the effects of chemotherapy [5]. While fertility preservation techniques exist, their applicability is constrained by factors like cancer stage and patient health [5]. Survivors of pre-menopausal ovarian cancer also face challenges including premature menopause, reduced ovarian reserve, and increased risk of longer term cardiovascular complications [31].

Finally, this study did not include a CUA, limiting a full assessment of cost-effectiveness and the impact of timely diagnosis on HRQoL and HRQoL decrements for misdiagnosed patients. However, no appropriate literature sources were identified to provide information on the disutility of screening for all types of patients with suspected ovarian cancer. Previous economic evaluations conducting CUA for ovarian cancer diagnostic testing have relied on sources limited by a small sample size and health state descriptions that do not generalise to current management of suspected ovarian cancer patients [4,32].

### Interpretation

Existing economic evaluations of ovarian cancer testing have examined fewer diagnostic strategies than the present study. For example, Forde et al. [33] compared multivariate index assay and CA125 testing; Kearns et al. [34] compared multimodal screening, ultrasound screening and no screening; Havrilesky et al. [35] looked at annual screening versus no screening.

Westwood et al. [21] assessed various diagnostic tests including RMI, ROMA, and ADNEX risk scores. Their economic evaluation found the ADNEX 10% strategy to be most cost-effective option at the £30,000 per QALY threshold in a lifetime Markov model. In contrast, we chose a CCA as a robust approach to representing the trade-offs involved in decision making, given no single outcome measure sufficiently captures patients both with and without ovarian cancer. This approach also allowed us to comprehensively map patient pathways rather than using a simplified version of treatment pathways following an ovarian cancer diagnosis.

## Conclusion

Earlier and better diagnostic strategies are essential to improving ovarian cancer outcomes. This study compared six of the available diagnostic strategies. A trade-off involved was found: the usual care strategy, RMI 200, was least costly in both cohorts, but its lower sensitivity meant more cancer deaths and lower diagnostic yield than an ADNEX 3% strategy. The results suggest a two-step strategy IOTA ADNEX 10% offers the best balance with near-comparable expected costs to RMI 200 whilst retaining significantly higher diagnostic yield and lower cancer deaths, making it highly suitable for implementation in practice.

The findings demonstrate that modifications to clinical pathways extend beyond diagnostic test accuracy; they fundamentally influence clinical decision-making, manifesting in differential costs and consequences across distinct patient cohorts. This is particularly evident in surgical intervention rates, specialist referral patterns, and long-term patient outcomes including cancer death.

Future economic evaluations should incorporate longer time horizons and account for broader quality-of-life impacts to enable further assessment of cost-effectiveness. Pertinent to this is future research into the quality-of-life impact of timely diagnosis, misdiagnosis, and the impact on pre-menopausal women facing infertility resulting from surgery and chemotherapy.

## Data Availability

The data that support the findings of this study are available from the corresponding author upon reasonable request.

## Acknowledgements

We thank the ROCkeTS project oversight committee (Chair— Peter Sasieni, members—Andy Nordin, Michael Weston, and Annwen Jones [Target Ovarian Cancer]) for their kind input and guidance. We acknowledge our gratitude to the patients who generously participated in our study. This study is funded by a grant from the NIHR Health Technology Assessment programme (HTA 13/13/01). The views expressed are those of the authors and not necessarily those of the NIHR or the Department of Health and Social Care.

## Contribution to Authorship

SS, TER, MM conceptualised and designed the study. SJP, MM, DG, EVW and TER were responsible for the economic analysis: SJP and MM did the economic analysis and received advice from DG, EVW and TER. MM supervised the economic analysis. SS contributed to the planning and interpretation of analysis. FLK and SW provided clinical input into the pathways. All authors reviewed the results and manuscript. JD, RA, and KS have directly accessed and verified the underlying data reported in the manuscript. All authors had full access to all the data in the study and accept responsibility to submit for publication.

## Details of Ethics Approval

ROCkeTS received ethical approval from NHS West Midlands REC (14/WM/1241) and is registered on the controlled trials website (ISRCTN17160843). Written informed consent was obtained from participants before participation.

## References

[1] Cancer Tomorrow. Global Cancer Observatory n.d. https://gco.iarc.who.int/today/ (accessed February 4, 2025).

[2] CRUK. What is ovarian cancer? 2024. https://www.cancerresearchuk.org/about-cancer/ovarian-cancer/what-is-ovarian-cancer (accessed December 18, 2024).

[3] Gaitskell K, Hermon C, Barnes I, Pirie K, Floud S, Green J, et al. Ovarian cancer survival by stage, histotype, and pre-diagnostic lifestyle factors, in the prospective UK Million Women Study. Cancer Epidemiology 2022;76:102074. 10.1016/j.canep.2021.102074.

[4] NICE. Ovarian cancer: recognition and initial management. Ovarian Cancer 2023. https://www.nice.org.uk/guidance/cg122/resources/ovarian-cancer-recognition-and-initial-management-35109446543557 (accessed February 10, 2025).

[5] Zapardiel I, Diestro MD, Aletti G. Conservative treatment of early stage ovarian cancer: Oncological and fertility outcomes. European Journal of Surgical Oncology (EJSO) 2014;40:387–93. 10.1016/j.ejso.2013.11.028.

[6] Sundar S, Rick C, Dowling F, Au P, Snell K, Rai N, et al. Refining Ovarian Cancer Test accuracy Scores (ROCkeTS): protocol for a prospective longitudinal test accuracy study to validate new risk scores in women with symptoms of suspected ovarian cancer. BMJ Open 2016;6:e010333. 10.1136/bmjopen-2015-010333.

[7] Target Ovarian Cancer. Pathfinder 2022: Faster, further, and fairer. London: n.d.

[8] Timmerman D, Planchamp F, Bourne T, Landolfo C, du Bois A, Chiva L, et al. ESGO/ISUOG/IOTA/ESGE Consensus Statement on preoperative diagnosis of ovarian tumors. Ultrasound in Obstetrics & Gynecology 2021;58:148–68. 10.1002/uog.23635.

[9] Strachowski LM, Jha P, Phillips CH, Blanchette Porter MM, Froyman W, Glanc P, et al. O-RADS US v2022: An Update from the American College of Radiology’s Ovarian-Adnexal Reporting and Data System US Committee. Radiology 2023;308:e230685. 10.1148/radiol.230685.

[10] Montagnana M, Danese E, Ruzzenente O, Bresciani V, Nuzzo T, Gelati M, et al. The ROMA (Risk of Ovarian Malignancy Algorithm) for estimating the risk of epithelial ovarian cancer in women presenting with pelvic mass: is it really useful? Clin Chem Lab Med 2011;49:521–5. 10.1515/CCLM.2011.075.

[11] Cummins C, Kumar S, Long J, Balega J, Broadhead T, Duncan T, et al. Investigating the Impact of Ultra-Radical Surgery on Survival in Advanced Ovarian Cancer Using Population-Based Data in a Multicentre UK Study. Cancers 2022;14:4362. 10.3390/cancers14184362.

[12] Royal College of Obstetricians and Gynaecologists. Ovarian Cysts in Postmenopausal Women. London: Royal College of Obstetricians and Gynaecologists; 2016.

[13] Royal College of Obstetricians and Gynaecologists, British Society for Gynaecological Endoscopy. Management of Suspected Ovarian Masses in Pre-menopausal Women. London: RCOG; 2011.

[14] Sundar S, Agarwal R, Davenport C, Scandrett K, Johnson S, Sengupta P, et al. Risk-prediction models in postmenopausal patients with symptoms of suspected ovarian cancer in the UK (ROCkeTS): a multicentre, prospective diagnostic accuracy study. The Lancet Oncology 2024;25:1371–86. 10.1016/S1470-2045(24)00406-6.

[15] Munzer A. TreeAge Software 2024.

[16] NICE. NICE health technology evaluations: the manual [PMG36]. Process and Methods [PMG36] 2022.

[17] Husereau D, Drummond M, Augustovski F, Bekker-Grob E de, Briggs AH, Carswell C, et al. Consolidated Health Economic Evaluation Reporting Standards 2022 (CHEERS 2022) statement: updated reporting guidance for health economic evaluations. BMJ 2022;376:e067975. 10.1136/bmj-2021-067975.

[18] NICE. Tests in secondary care to identify people at high risk of ovarian cancer. Diagnostic guidance DG31 2017. https://www.nice.org.uk/guidance/dg31/chapter/3-The-diagnostic-tests (accessed February 4, 2025).

[19] Van Calster B, Van Hoorde K, Froyman W, Kaijser J, Wynants L, Landolfo C, et al. Practical guidance for applying the ADNEX model from the IOTA group to discriminate between different subtypes of adnexal tumors. Facts Views Vis Obgyn 2015;7:32–41.

[20] Yang S, Tang J, Rong Y, Wang M, Long J, Chen C, et al. Performance of the IOTA ADNEX model combined with HE4 for identifying early-stage ovarian cancer. Front Oncol 2022;12:949766. 10.3389/fonc.2022.949766.

[21] Westwood M, Ramaekers B, Lang S, Grimm S, Deshpande S, de Kock S, et al. Risk scores to guide referral decisions for people with suspected ovarian cancer in secondary care: a systematic review and cost-effectiveness analysis. Health Technol Assess 2018;22:1–264. 10.3310/hta22440.

[22] Jones KC, Weatherly H, Birch S, Castelli A, Chalkley M, Dargan A, et al. Unit Costs of Health and Social Care 2023 Manual 2024. 10.22024/UniKent/01.02.100519.

[23] NHS England Digital,. Cancer Survival in England, cancers diagnosed 2016 to 2020, followed up to 2021. NHS England Digital 2023. https://digital.nhs.uk/data-and-information/publications/statistical/cancer-survival-in-england/cancers-diagnosed-2016-to-2020-followed-up-to-2021 (accessed December 13, 2024).

[24] Vorst REWM van de, Hoogendam JP, Aa MA van der, Witteveen PO, Zweemer RP, Gerestein CG. The attributive value of comprehensive surgical staging in clinically early-stage epithelial ovarian carcinoma: A systematic review and meta-analysis. Gynecologic Oncology 2021;161:876–83. 10.1016/j.ygyno.2021.04.007.

[25] Landolfo C, Bourne T, Froyman W, Van Calster B, Ceusters J, Testa AC, et al. Benign descriptors and ADNEX in two-step strategy to estimate risk of malignancy in ovarian tumors: retrospective validation in IOTA5 multicenter cohort. Ultrasound in Obstetrics & Gynecology 2023;61:231–42. 10.1002/uog.26080.

[26] Nunes N, Ambler G, Foo X, Naftalin J, Derdelis G, Widschwendter M, et al. Comparison of two protocols for the management of asymptomatic postmenopausal women with adnexal tumours - a randomised controlled trial of RMI/RCOG vs Simple Rules. Br J Cancer 2017;116:584–91. 10.1038/bjc.2017.17.

[27] Briggs A, Fenn P. Confidence intervals or surfaces? Uncertainty on the cost-effectiveness plane. Health Economics 1998;7:723–40. 10.1002/(SICI)1099-1050(199812)7:8<723::AID-HEC392>3.0.CO;2-O.

[28] Kwong FLA, Kristunas C, Davenport C, Deeks J, Mallett S, Agarwal R, et al. Symptom-triggered testing detects early stage and low volume resectable advanced stage ovarian cancer. International Journal of Gynecologic Cancer 2024.

[29] Kwong FL, Kristunas C, Davenport C, Aggarwal R, Deeks J, Mallett S, et al. Investigating harms of testing for ovarian cancer – psychological outcomes and cancer conversion rates in women with symptoms of ovarian cancer: A cohort study embedded in the multicentre ROCkeTS prospective diagnostic study. BJOG: An International Journal of Obstetrics & Gynaecology 2024;131:1400–10. 10.1111/1471-0528.17813.

[30] Davenport CF, Rai N, Sharma P, Deeks J, Berhane S, Mallett S, et al. Diagnostic Models Combining Clinical Information, Ultrasound and Biochemical Markers for Ovarian Cancer: Cochrane Systematic Review and Meta-Analysis. Cancers 2022;14:3621. 10.3390/cancers14153621.

[31] Roof KA, Andre KE, Modesitt SC, Schirmer DA. Maximizing ovarian function and fertility following chemotherapy in premenopausal patients: Is there a role for ovarian suppression? Gynecologic Oncology Reports 2024;53:101383. 10.1016/j.gore.2024.101383.

[32] Havrilesky LJ, Broadwater G, Davis DM, Nolte KC, Barnett JC, Myers ER, et al. Determination of quality of life-related utilities for health states relevant to ovarian cancer diagnosis and treatment. Gynecologic Oncology 2009;113:216–20. 10.1016/j.ygyno.2008.12.026.

[33] Forde GK, Hornberger J, Michalopoulos S, Bristow RE. Cost-effectiveness analysis of a multivariate index assay compared to modified American College of Obstetricians and Gynecologists criteria and CA-125 in the triage of women with adnexal masses. Curr Med Res Opin 2016;32:321–9. 10.1185/03007995.2015.1123679.

[34] Kearns B, Chilcott J, Whyte S, Preston L, Sadler S. Cost-effectiveness of screening for ovarian cancer amongst postmenopausal women: a model-based economic evaluation. BMC Medicine 2016;14:200. 10.1186/s12916-016-0743-y.

[35] Havrilesky LJ, Sanders GD, Kulasingam S, Myers ER. Reducing ovarian cancer mortality through screening: Is it possible, and can we afford it? Gynecol Oncol 2008;111:179–87. 10.1016/j.ygyno.2008.07.006.

[36] NHS Improvement. 2022/23 National Cost Collection Data Publication. 2022/23 National Cost Collection Data Publication 2024. https://www.england.nhs.uk/publication/2022-23-national-cost-collection-data-publication/ (accessed October 28, 2024).

[37] Joint Formulary Committee,. British National Formulary (BNF). British National Formulary (Online) 2024. http://www.medicinescomplete.com (accessed October 28, 2024).

[38] Eccleston A, Bentley A, Dyer M, Strydom A, Vereecken W, George A, et al. A Cost-Effectiveness Evaluation of Germline BRCA1 and BRCA2 Testing in UK Women with Ovarian Cancer. Value Health 2017;20:567–76. 10.1016/j.jval.2017.01.004.

[39] Elsea D, Muston D, Fan L, Mihai A, Meng Y, Kasle A, et al. Cost-Effectiveness Analysis of Biomarker Testing to Guide First-Line PARP Inhibitor Maintenance for Patients with Advanced Ovarian Cancer After Response to First-Line Platinum Chemotherapy in the USA. Targ Oncol 2023;18:531–41. 10.1007/s11523-023-00966-6.

[40] National Cancer Registration and Analysis Service. Ovarian Cancer Audit Feasibility Pilot (OCAFP) geographic variation in ovarian, fallopian tube and primary peritoneal cancer treatment in England 2023. https://digital.nhs.uk/ndrs/data/data-outputs/cancer-publications-and-tools/ovarian-cancer-audit-feasibility-pilot---geographic-variation (accessed October 28, 2024).

