## Supporting Information for "A cost consequence analysis of six diagnostic strategies for ovarian cancer: A model-based economic evaluation"

### **Table S1 Cost inputs**

| **Variable** | **Value (£)** | **PSA Distribution (parameters)** | **Source** |
| --- | --- | --- | --- |
| Cost of 'all' surgery | 12675.70 | Gamma (α=25;λ=  0.00197) | NHS 2022/23 reference costs [8], weighted average of currency codes F50A to F50C, Elective Inpatients |
| Cost of omentectomy for 'some' surgery | 8365.08 | Gamma (α=25; λ=0.00299) | NHS 2022/23 reference costs [8], weighted average of currency codes FF51A to FF51J, Elective Inpatients |
| Per cycle cost of bevacizumab drug | 1140.27 | Gamma (α=25; λ=0.02192) | Weighted average cost of 1.313 Bevacizumab vials per cycle[9] |
| Cost of administration of bevacizumab as maintenance therapy | 253 | Gamma (α=25; λ=0.09881) | NHS 2022/23 reference costs [8], currency code SB15Z |
| Cost of anterior resection of rectum and exteriorisation of the bowel for 'all' surgery | 13205.48 | Gamma (α=25; λ=0.00189) | NHS reference costs 2022/23[8], weighted average of currency codes FF31A to FF31D, Elective Inpatient |
| Cost of serum CA125 for diagnostic testing (Roche) | 5.62 | Gamma (α=25; λ=4.44840) | Personal communication from Roche diagnostics |
| Cost of one cycle of carboplatin for chemotherapy (drug only) | 336.13 | Gamma (α=25; λ=0.07438) | Weighted average cost of carboplatin 1.25 vials of 60 ml [9] |
| Delivery of CarboTaxol regimen (first) | 277.00 | Gamma (α=25; λ=0.09025) | NHS reference costs 2022/23[8], currency code SB13Z (“Deliver more complex perenteral chemotherapy at first attendance (outpatient”), Outpatient |
| Cost of subsequent elements of chemotherapy cycle | 249 | Gamma (α=25; λ=0.09881) | NHS reference costs 2022/23[8], currency code SB15Z |
| Delivery of complex chemotherapy admin (cost per cycle) | 361.00 | Gamma (α=25; λ= 0.10040) | NHS reference costs 2022/23[8], currency code SB14Z |
| CT scan of three areas (thorax, abdomen, pelvis) with contrast - for chemo | 167.63 | Gamma (α=25; λ= 0.14914) | NHS reference costs 2022/23[8], currency code RD26Z |
| Per cycle cost of dexamethasone (all regimens) | 18.52 | Gamma (α=25; λ=1.34989) | Weighted average cost of 6 tablets of dexamethasone [9] |
| Cost of diaphragm resection (Other specified operations on diaphragm) for 'all' surgery | 8309.30 | Gamma (α=25; λ=0.00301) | NHS reference costs 2022/23[8], weighted average of currency codes DZ63A to DZ63C |
| Major, laparoscopic or endoscopic, upper genital tract procedures (bilateral salpingo-oopherectomy) | 6275.10 | Gamma (α=25; λ=0.00398) | NHS 2022/23 reference costs [8], weighted average of currency codes MA08A and MA08B |
| Total abdominal hysterectomy and bilateral salpingo-oopherectomy for 'all' surgery | 9,952.00 | Gamma (α=25; λ=0.00251) | NHS 2022/23 reference costs [8], weighted average of currency codes MA26A to MA26C |
| Total abdominal hysterectomy plus BSO for 'some' surgery | 7054.12 | Gamma (α=25; λ=0.00354) | NHS 2022/23 reference costs [8], weighted average of currency codes MA06A to MA06 |
| Cost of germline BRCA test | 370.76 | Gamma (α=25; λ=0.06743) | Test and 2 counselling sessions: Ecclestone et al. (2017)[10], PSSRU band 7 hospital-based scientific and professional staff [7] |
| Cost of gyno (non-consultant) follow-up for TNs | 108.28 | Gamma (α=25; λ= 0.23088) | NHS 2022/23 reference costs [8], currency code WF01A |
| Cost per consultant-led gynaecological oncology service follow-up | 168.20 | Gamma (α=25; λ= 0.14863) | NHS 2022/23 reference costs [8], currency code WF01A |
| Cost of HE4 test for ROMA | 31.39 | Gamma (α=25; λ=0.79643) | Test kit: Roche personal communication; Quality control, Calibration, Maintenance, Capital, and Personnel: Westwood (2018)[11] |
| Cost of histology | 82.06 | Gamma (α=25; λ=0.30466) | NHS 2022/23 reference costs [8], currency code DAPS02 |
| Cost of MyChoice® CDx HRD test | 3561.46 | Gamma (α=25; λ=0.00702) | Elsea (2023)[12] converted to GBP using average 2019 exchange rate of 0.7835 and inflated |
| Cost of image-guided biopsy for NACT | 925.17 | Gamma (α=25; λ=0.02702) | NHS 2022/23 reference costs [8], currency code YF05Z |
| Cost of gynaecological oncology MDT review | 478.66 | Gamma (α=25; λ=0.05223) | NHS 2022/23 reference costs [8], currency code CMDTSpG |
| Cost per cycle of metoclopramide (all regimens) | 0.42 | Gamma (α=25; λ=59.52381) | Weighted average cost of ½ packet of metoclopramide [9] |
| MRI scan of one area, w/o contrast | 175.93 | Gamma (α=25; λ=0.06745) | NHS 2022/23 reference costs [8], currency code RD01A |
| Cost per 28-day cycle of Niraparib | 6750 | Gamma (α=25; λ=0.00370) | 1 pack of Niraparib [9] |
| Cost of 28-day cycle of Olaparib | 2317.50 | Gamma (α=25; λ=0.01079) | 4 x 150mg tablets of Olaparib, taken in sets of two every day[9] |
| Splenectomy | 7719.28 | Gamma (α=25; λ=0.00324) | NHS 2022/23 reference costs [8], weighted average of currency code GA07C to GA07E |
| CT scan of three areas (thorax, abdomen, pelvis) with contrast | 167.63 | Gamma (α=25; λ=0.14914) | NHS 2022/23 reference costs [8], currency code RD26Z |
| Cost of staging laparotomy | 7054.12 | Gamma (α=25; λ=0.00354) | NHS 2022/23 reference costs [8], weighted average of currency codes MA06A to MA06 |
| Cost per cycle of paclitaxel | 1028.04 | Gamma (α=25; λ=0.02432) | Weighted average cost of 1 vial per cycle of paclitaxel [9] |
| Transvaginal pelvic ultrasound | 242.65 | Gamma (α=25; λ= 0.10303) | NHS 2022/23 reference costs [8], currency code MA36Z. |

### **Table S2 Clinical model inputs pre-menopausal cohort**

| **Variable** | **Value** | **PSA Distribution (parameters)** | **Source** |
| --- | --- | --- | --- |
| Ovarian cancer proportion | 0.0613 | Beta (α=49, β=750) | ROCkeTS dataset |
| ADNEX 10% strategy:  Proportion of FNs with ADNEX scores within 1-10% range that have Stage I OC | 0.667 | Beta (α=2, β=1) | ROCkeTS dataset |
| ADNEX 10% strategy:  Proportion of FNs with ADNEX scores within 1-10% range that have Stage II/III OC out of those with Stage II-IV OC | 0 | Fixed | ROCkeTS dataset |
| ADNEX 10% strategy:  Proportion of FNs with ADNEX scores < 1% out of those with ADNEX scores <10% | 0.4 | Beta (α=2, β=3) | ROCkeTS dataset |
| ADNEX 10% strategy; ADNEX 3% strategy:  Proportion of FNs with ADNEX scores <1% that have Stage I OC | 0 | Fixed | ROCkeTS dataset |
| ADNEX 10% strategy; ADNEX 3% strategy:  Proportion of FNs with ADNEX scores <1% that have Stage II/III OC relative to those with Stages II-IV OC | 0 | Fixed | ROCkeTS dataset |
| ADNEX 3% strategy: Proportion of FNs with ADNEX scores within 1-3% range that have Stage I OC | 0 | Fixed | ROCkeTS dataset |
| ADNEX 3% strategy: Proportion of FNs with ADNEX scores within 1-3% range that have Stage II/III OC relative to those with Stages II-IV OC | 0 | Fixed | ROCkeTS dataset |
| ADNEX 3% strategy:  Proportion of FNs with ADNEX scores < 1% out of those with ADNEX scores <3% | 0.667 | Beta (α=2, β=1) | ROCkeTS dataset |
| ADNEX 10% strategy:  Proportion of FPs relative to population with no OC | 0.249 | Beta (α=142, β=429) | ROCkeTS dataset |
| ADNEX 3% strategy:  Proportion of FPs relative to population with no OC | 0.545 | Beta (α=311, β=260) | ROCkeTS dataset |
| ADNEX 10% strategy:  Proportion of ADNEX 10% TNs with ADNEX scores <1% | 0.249 | Beta (α=62, β=187) | ROCkeTS dataset |
| ADNEX 3% strategy:  Proportion of ADNEX 3% TNs with ADNEX scores <1% | 0.238 | Beta (α=62, β=198) | ROCkeTS dataset |
| ADNEX 3% strategy:  Proportion of TPs relative to population with OC | 0.935 | Beta (α=43, β=3) | ROCkeTS dataset |
| Proportion of Stages II-IV Cancer patients having primary surgery | 1 | Fixed | ROCkeTS dataset |
| ADNEX 10% strategy:  Proportion of TPs relative to population with OC | 0.891 | Beta (α=41, β=5) | ROCkeTS dataset |
| ADNEX 10% strategy:  Proportion of FPs having surgery for likely benign histology | 0.796 | Beta (α=113, β=29) | ROCkeTS dataset |
| ADNEX 3% strategy:  Proportion of FPs having surgery for likely benign histology | 0.745 | Beta (α=231, β=79) | ROCkeTS dataset |
| ROMA 11.4%strategy:  Proportion of FPs having surgery for likely benign histology | 0.650 | Beta (α=13, β=7) | ROCkeTS dataset |
| CA 125 strategy:  Proportion of FPs having surgery for likely benign histology | 0.608 | Beta (α=161, β=104) | ROCkeTS dataset |
| ORADS strategy:  Proportion of FPs having surgery for likely benign histology | 0.798 | Beta (α=87, β=22) | ROCkeTS dataset |
| RMI 200 strategy:  Proportion of FPs having surgery for likely benign histology | 0.759 | Beta (α=22, β=7) | ROCkeTS dataset |
| ORADS strategy:  Proportion of FNs with Stage I OC | 0.4 | Beta (α=4, β=6) | ROCkeTS dataset |
| CA 125 strategy:  Proportion of FNs with Stage I OC | 0.636 | Beta (α=7, β=4) | ROCkeTS dataset |
| ORADS strategy:  Proportion of FNs that have Stage II/III OC relative to those with Stages II-IV OC | 0.667 | Beta (α=4, β=2) | ROCkeTS dataset |
| ORADS strategy:  Proportion of FPs relative to population with no OC | 0.151 | Beta (α=109, β=615) | ROCkeTS dataset |
| ORADS strategy:  Proportion of TPs relative to population with OC | 0.792 | Beta (α=38, β=10) | ROCkeTS dataset |
| RMI 200 strategy:  Proportion of FNs with RMI score of 25-200 with Stage I OC | 0.579 | Beta (α=11, β=8) | ROCkeTS dataset |
| RMI 200 strategy:  Proportion of FNs with RMI score of 25-200 that have Stage II/III OC relative to those with Stages II-IV OC | 0.5 | Beta (α=1, β=1) | ROCkeTS dataset |
| RMI 200 strategy:  Proportion of RMI 200 FNs with score < 25 | 0.208 | Beta (α=5, β=19) | ROCkeTS dataset |
| RMI 200 strategy:  Proportion of FNs with RMI score <25 with Stage I OC | 0.6 | Beta (α=3, β=2) | ROCkeTS dataset |
| RMI 200 strategy  Proportion of FNs with RMI score <25 that have Stage II/III OC relative to those with Stages II-IV OC | 1 | Fixed | ROCkeTS dataset |
| RMI 200 strategy:  Proportion of TNs with RMI score <25 | 0.607 | Beta (α=362, β=234) | ROCkeTS dataset |
| RMI 200 strategy:  Proportion of FPs relative to population with no OC | 0.046 | Beta (α=29, β=596) | ROCkeTS dataset |
| RMI 200 strategy:  Proportion of TPs relative to population with OC | 0.489 | Beta (α=23, β=24) | ROCkeTS dataset |
| ROMA 11.4% strategy:  Proportion of FNs with Stage I OC | 0.7 | Beta (α=7, β=3) | ROCkeTS dataset |
| ROMA 11.4%strategy:  Proportion of FNs that have Stage II/III OC relative to those with Stages II-IV OC | 0% | Fixed | ROCkeTS dataset |
| ROMA 11.4% strategy: Proportion of FPs relative to population with no OC | 0.028 | Beta (α=20, β=688) | ROCkeTS dataset |
| ROMA 11.4% strategy:  Proportion of TPs relative to population with OC | 0.583 | Beta (α=28, β=20) | ROCkeTS dataset |
| ADNEX 10% strategy:  Proportion of ADNEX 1-10% TNs having surgery for benign/symptoms | 0.529 | Beta (α=193, β=172) | ROCkeTS dataset |
| ADNEX 3% strategy:  Proportion of 1-3% ADNEX TNs having surgery for benign/symptoms | 0.452 | Beta (α=89, β=108) | ROCkeTS dataset |
| CA 125 strategy:  Proportion of TNs that have surgery for benign/symptoms | 0.479 | Beta (α=229, β=249) | ROCkeTS dataset |
| ORADS strategy:  Proportion of TNs that have surgery for benign/symptoms | 0.491 | Beta (α=300, β=311) | ROCkeTS dataset |
| RMI 200 strategy:  Proportion of RMI 25-200 TNs having surgery for benign/symptoms | 0.564 | Beta (α=132, β=102) | ROCkeTS dataset |
| ROMA 11.4%strategy:  Proportion of TNs that have surgery for benign/symptoms | 0.482 | Beta (α=328, β=352) | ROCkeTS dataset |
| ADNEX 10% strategy:  Proportion of TPs with Stage I OC | 0.439 | Beta (α=18, β=23) | ROCkeTS dataset |
| ADNEX 3% strategy:  Proportion of TPs with Stage I OC | 0.465 | Beta (α=20, β=23) | ROCkeTS dataset |
| CA 125 strategy:  Proportion of TPs with Stage I OC | 0.342 | Beta (α=13, β=25) | ROCkeTS dataset |
| ORADS strategy:  Proportion of TPs with Stage I OC | 0.421 | Beta (α=16, β=22) | ROCkeTS dataset |
| RMI 200 strategy:  Proportion of TPs with Stage I OC | 0.261 | Beta (α=6, β=17) | ROCkeTS dataset |
| ROMA 11.4%strategy:  Proportion of TPs with Stage I OC | 0.342 | Beta (α=13, β=25) | ROCkeTS dataset |
| ADNEX 10% strategy:  Proportion of TPs that have Stage II/III OC relative to those with Stages II-IV OC | 0.826 | Beta (α=19, β=4) | ROCkeTS dataset |
| ADNEX 3% strategy:  Proportion of TPs that have Stage II/III OC relative to those with Stages II-IV OC | 0.826 | Beta (α=19, β=4) | ROCkeTS dataset |
| CA 125 strategy:  Proportion of TPs that have Stage II/III OC relative to those with Stages II-IV OC | 0.8 | Beta (α=20, β=5) | ROCkeTS dataset |
| ORADS strategy:  Proportion of TPs that have Stage II/III OC relative to those with Stages II-IV OC | 0.727 | Beta (α=16, β=6) | ROCkeTS dataset |
| RMI 200 strategy:  Proportion of TPs that have Stage II/III OC relative to those with Stages II-IV OC | 0.824 | Beta (α=14, β=3) | ROCkeTS dataset |
| ROMA 11.4%strategy:  Proportion of TPs that have Stage II/III OC relative to those with Stages II-IV OC | 0.8 | Beta (α=20, β=5) | ROCkeTS dataset |
| Proportion of Stage II-IV patients that get all listed surgery procedures (for primary surgery, interval debulking, delayed debulking) | 0.35 | Beta (α=3.5, β=6.5) | Assumption |
| Stage I OC death at 12 months | 0.022 | Beta (α=192, β= 8,526) | NHS digital[13] |
| Stage II and III OC death at 12 months | 0.232 | Beta (α=2483, β=8,230) | NHS digital[13] |
| Stage IV OC death at 12 months | 0.437 | Beta (α=2635, β= 3394) | NHS digital[13] |
| Proportion upstaged | 0.23 | Beta (α= 27.59, β= 92.36) | Van de Vorst (2021)[14] |
| Stage I follow-up after staging laparotomy | 0.57 | Beta (α=1763, β=1328) | OCAFP[15] |
| Proportion of FPs that have additional MRI | 0.35 | Beta (α=3.5, β=6.5) | Assumption from local clinic audit data |
| OC: Ovarian Cancer; FP: False Positive; TP: True Positive; FN: False Negative; TN: True Negative; HRD: homologous recombination deficiency; MRI: Magnetic resonance imaging; | | | |

### **Table S3 Clinical model inputs post-menopausal cohort**

| **Variable** | **Value** | **PSA Distribution (parameters)** | **Source** |
| --- | --- | --- | --- |
| Ovarian cancer proportion | 0.199 | Beta (α=214, β=860) | ROCkeTS dataset |
| ADNEX 10% strategy:  Proportion of FNs with ADNEX scores within 1-10% range that have Stage I OC | 0.714 | Beta (α=5, β=2) | ROCkeTS dataset |
| ADNEX 10% strategy:  Proportion of FNs with ADNEX scores within 1-10% range that have Stage II/III OC out of those with Stage II-IV OC | 0.500 | Beta (α=1, β=1) | ROCkeTS dataset |
| ADNEX 10% strategy:  Proportion of FNs with ADNEX scores < 1% out of those with ADNEX scores <10% | 0 | Fixed | ROCkeTS dataset |
| ADNEX 10% strategy; ADNEX 3% strategy:  Proportion of FNs with ADNEX scores <1% that have Stage I OC | 0 | Fixed | ROCkeTS dataset |
| ADNEX 10% strategy; ADNEX 3% strategy:  Proportion of FNs with ADNEX scores <1% that have Stage II/III OC relative to those with Stages II-IV OC | 0 | Fixed | ROCkeTS dataset |
| ADNEX 3% strategy:  Proportion of FNs with ADNEX scores within 1-3% range that have Stage I OC | 0 | Fixed | ROCkeTS dataset |
| ADNEX 3% strategy:  Proportion of FNs with ADNEX scores within 1-3% range that have Stage II/III OC relative to those with Stages II-IV OC | 0 | Fixed | ROCkeTS dataset |
| ADNEX 3% strategy:  Proportion of FNs with ADNEX scores < 1% out of those with ADNEX scores <3% | 0 | Fixed | ROCkeTS dataset |
| ADNEX 10% strategy:  Proportion of FPs relative to population with no OC | 0.415 | Beta (α=295, β=415) | ROCkeTS dataset |
| ADNEX 3% strategy:  Proportion of FPs relative to population with no OC | 0.692 | Beta (α=491, β=219) | ROCkeTS dataset |
| ADNEX 10% strategy:  Proportion of ADNEX 10% TNs with ADNEX scores <1% | 0.145 | Beta (α=60, β=355) | ROCkeTS dataset |
| ADNEX 3% strategy:  Proportion of ADNEX 3% TNs with ADNEX scores <1% | 0.274 | Beta (α=60, β=159) | ROCkeTS dataset |
| ADNEX 3% strategy:  Proportion of TPs relative to population with OC | 1 | Fixed | ROCkeTS dataset |
| Proportion of Stages II-IV Cancer patients having primary surgery | 1 | Fixed | ROCkeTS dataset |
| ADNEX 10% strategy:  Proportion of TPs relative to population with OC | 0.961 | Beta (α=173, β=7) | ROCkeTS dataset |
| ADNEX 10% strategy:  Proportion of FPs having surgery for likely benign histology | 0.416 | Beta (α=121, β=170) | ROCkeTS dataset |
| ADNEX 3% strategy:  Proportion of FPs having surgery for likely benign histology | 0.440 | Beta (α=211, β=269) | ROCkeTS dataset |
| ROMA 29.9% strategy:  Proportion of FPs having surgery for likely benign histology | 0.245 | Beta (α=37, β=114) | ROCkeTS dataset |
| CA 125 strategy:  Proportion of FPs having surgery for likely benign histology | 0.268 | Beta (α=51, β=139) | ROCkeTS dataset |
| ORADS strategy:  Proportion of FPs having surgery for likely benign histology | 0.335 | Beta (α=61, β=121) | ROCkeTS dataset |
| RMI 200 strategy:  Proportion of FPs having surgery for likely benign histology | 0.303 | Beta (α=37, β=85) | ROCkeTS dataset |
| ORADS strategy:  Proportion of FNs with Stage I OC | 0.245 | Beta (α=12, β=37) | ROCkeTS dataset |
| CA 125 strategy:  Proportion of FNs with Stage I OC | 0.567 | Beta (α=17, β=13) | ROCkeTS dataset |
| ORADS strategy:  Proportion of FNs that have Stage II/III OC relative to those with Stages II-IV OC | 0.703 | Beta (α=26, β=11) | ROCkeTS dataset |
| ORADS strategy:  Proportion of FPs relative to population with no OC | 0.217 | Beta (α=182, β=655) | ROCkeTS dataset |
| ORADS strategy:  Proportion of TPs relative to population with OC | 0.764 | Beta (α=159, β=49) | ROCkeTS dataset |
| RMI 200 strategy:  Proportion of FNs with RMI score of 25-200 with Stage I OC | 0.565 | Beta (α=13, β=10) | ROCkeTS dataset |
| RMI 200 strategy:  Proportion of FNs with RMI score of 25-200 that have Stage II/III OC relative to those with Stages II-IV OC | 0.800 | Beta (α=8, β=2) | ROCkeTS dataset |
| RMI 200 strategy:  Proportion of RMI 200 FNs with score < 25 | 0.179 | Beta (α=5, β=23) | ROCkeTS dataset |
| RMI 200 strategy:  Proportion of FNs with RMI score <25 with Stage I OC | 0.600 | Beta (α=3, β=2) | ROCkeTS dataset |
| RMI 200 strategy  Proportion of FNs with RMI score <25 that have Stage II/III OC relative to those with Stages II-IV OC | 1 | Fixed | ROCkeTS dataset |
| RMI 200 strategy:  Proportion of TNs with RMI score <25 | 0.368 | Beta (α=362, β=234) | ROCkeTS dataset |
| RMI 200 strategy:  Proportion of FPs relative to population with no OC | 0.156 | Beta (α=29, β=596) | ROCkeTS dataset |
| RMI 200 strategy:  Proportion of TPs relative to population with OC | 0.850 | Beta (α=23, β=24) | ROCkeTS dataset |
| ROMA 29.9% strategy:  Proportion of FNs with Stage I OC | 0.696 | Beta (α=16, β=7) | ROCkeTS dataset |
| ROMA 29.9% strategy:  Proportion of FNs with that have Stage II/III OC relative to those with Stages II-IV OC | 0.714 | Beta (α=5, β=2) | ROCkeTS dataset |
| ROMA 29.9% strategy: Proportion of FPs relative to population with no OC | 0.201 | Beta (α=154, β=612) | ROCkeTS dataset |
| ROMA 29.9% strategy:  Proportion of TPs relative to population with OC | 0.880 | Beta (α=168, β=23) | ROCkeTS dataset |
| ADNEX 10% strategy:  Proportion of ADNEX 1-10% TNs having surgery for benign/symptoms | 0.425 | Beta (α=151, β=204) | ROCkeTS dataset |
| ADNEX 3% strategy:  Proportion of 1-3% ADNEX TNs having surgery for benign/symptoms | 0.388 | Beta (α=85, β=134) | ROCkeTS dataset |
| CA 125 strategy:  Proportion of TNs that have surgery for benign/symptoms | 0.427 | Beta (α=286, β=382) | ROCkeTS dataset |
| ORADS strategy:  Proportion of TNs that have surgery for benign/symptoms | 0.408 | Beta (α=271, β=394) | ROCkeTS dataset |
| RMI 200 strategy:  Proportion of RMI 25-200 TNs having surgery for benign/symptoms | 0.447 | Beta (α=189, β=234) | ROCkeTS dataset |
| ROMA 29.9% strategy:  Proportion of TNs that have surgery for benign/symptoms | 0.426 | Beta (α=261, β=351) | ROCkeTS dataset |
| Proportion of Stage II-IV patients that get all listed surgery procedures (for primary surgery, interval debulking, delayed debulking) | 0.35 | Beta (α=3.5, β=6.5) | Assumption |
| ADNEX 10% strategy:  Proportion of TPs with Stage I OC | 0.324 | Beta (α=56, β=117) | ROCkeTS dataset |
| ADNEX 3% strategy:  Proportion of TPs with Stage I OC | 0.339 | Beta (α=61, β=119) | ROCkeTS dataset |
| CA 125 strategy:  Proportion of TPs with Stage I OC | 0.261 | Beta (α=48, β=136) | ROCkeTS dataset |
| ORADS strategy:  Proportion of TPs with Stage I OC | 0.321 | Beta (α=51, β=159) | ROCkeTS dataset |
| RMI 200 strategy:  Proportion of TPs with Stage I OC | 0.268 | Beta (α=45, β=168) | ROCkeTS dataset |
| ROMA 29.9% strategy:  Proportion of TPs with Stage I OC | 0.289 | Beta (α=46, β=113) | ROCkeTS dataset |
| ADNEX 10% strategy:  Proportion of TPs that have Stage II/III OC relative to those with Stages II-IV OC | 0.821 | Beta (α=96, β=21) | ROCkeTS dataset |
| ADNEX 3% strategy:  Proportion of TPs that have Stage II/III OC relative to those with Stages II-IV OC | 0.815 | Beta (α=97, β=22) | ROCkeTS dataset |
| CA 125 strategy:  Proportion of TPs that have Stage II/III OC relative to those with Stages II-IV OC | 0.787 | Beta (α=107, β=29) | ROCkeTS dataset |
| ORADS strategy:  Proportion of TPs that have Stage II/III OC relative to those with Stages II-IV OC | 0.815 | Beta (α=88, β=20) | ROCkeTS dataset |
| RMI 200 strategy:  Proportion of TPs that have Stage II/III OC relative to those with Stages II-IV OC | 0.805 | Beta (α=91, β=22) | ROCkeTS dataset |
| ROMA 29.9% strategy:  Proportion of TPs that have Stage II/III OC relative to those with Stages II-IV OC | 0.8 | Beta (α=97, β=26) | ROCkeTS dataset |
| Stage I OC death at 12 months | 0.022 | Beta (α=192, β= 8,526) | NHS digital[13] |
| Stage II and III OC death at 12 months | 0.232 | Beta (α=2483, β=8,230) | NHS digital[13] |
| Stage IV OC death at 12 months | 0.437 | Beta (α=2635, β= 3394) | NHS digital[13] |
| FN Stage 1 proportion upstaged | 0.230 | Beta (α= 27.59, β= 92.36) | Van de Vorst (2021)[14] |
| Proportion of True positives with Stage I OC | 0.302 | Beta (α=65, β=150) | ROCkeTS dataset |
| Stage I follow-up after staging laparotomy | 0.57 | Beta (α=1763, β=1328) | OCAFP[15] |
| Proportion of FPs that have additional MRI | 0.35 | Beta (α=3.5, β=6.5) | Assumption from local clinic audit data |
| OC: Ovarian Cancer; FP: False Positive; TP: True Positive; FN: False Negative; TN: True Negative; HRD: homologous recombination deficiency; MRI: Magnetic resonance imaging; | | | |

**Description of Patient Pathway Assumptions by Diagnostic Result**

True Positive

All TP cases undergo a staging CT to determine disease stage. Based on results, patients were classified into Stage I or Stage II-IV. Proportions for these stages were assumed from ROCkeTS data, with patients missing FIGO stage being categorised and treated as Stage IV. All TP diagnosis surgery was conducted in tertiary care setting.

For Stage I patients, all undergo staging laparotomy, which includes total abdominal hysterectomy, bilateral salpingo-oophorectomy, omental biopsy, and pelvic and para-aortic node sampling. Additionally, all Stage I patients receive germline testing for BRCA mutations. Following surgery, 57.0% of Stage I patients proceed directly to follow-up care involving two visits, as reported in the OCAFP second report [15]. The remaining 43.0% receive adjuvant chemotherapy of CarboTaxol, followed by two follow-up visits. All Stage I patients are assumed to be disease-free at 12 months.

For patients with Stage II-IV disease, all undergo the MyChoice HRD test, with those testing positive either having a BRCA mutation or being HRD-positive. These patients are assumed to have similar survival outcomes given the 12-month timeframe of the model. All patients have germline testing for BRCA mutation, those positive for BRCA mutation are treated the same as those who are somatic (tumour) BRCA mutation positive.

Treatment for Stage II-IV cases involves either primary surgery plus adjuvant chemotherapy (CarboTaxol) or primary chemotherapy with planned interval debulking for those deemed unresectable. It was assumed that all Stage II-III patients undergo primary surgery, while all Stage IV patients have interval debulking surgery. Those having primary surgery can either have ‘partial’ procedures (total abdominal hysterectomy, bilateral salpingo-oopherectomy, omentectomy) or ‘extensive’ procedures (some plus bowel resection, cholecystectomy, splenectomy, peritonectomy, diaphragmatic resection, pleural draining, excision of enlarged lymph nodes). We have assumed that 35% have ‘extensive’ procedures. The rest of the patients have ‘partial’ procedures. Following surgery, patients receive six cycles of CarboTaxol, followed by six months of maintenance therapy. Maintenance therapy was Olaparib for HRD-positive patients and Niraparib for HRD-negative patients along with two follow-up visits for both types of patients.

For Stage IV patients receiving primary chemotherapy, an image-guided biopsy is performed prior to treatment, with no surgery initially. These patients receive three cycles of CarboTaxol plus bevacizumab, followed by interval debulking involving all surgical procedures. After surgery, patients receive an additional three cycles of CarboTaxol plus bevacizumab, followed by maintenance therapy. HRD-positive patients continue with Olaparib, while HRD-negative patients receive bevacizumab.

False Negatives

FN cases either undergo surgery in secondary care (Supporting Information Figure S4) or are diagnosed after a six-month follow-up (Supporting Information Figure S5). In this analysis, only those who had surgery for suspected benign histology or based on symptoms indicating the need for suspected benign histology or based on symptoms indicating the need for surgery are considered. Data for FNs are drawn from the true negative (TN) cases in ROCkeTS, as FNs and TNs are treated as a homogenous group in this context. Given the hypothetical nature of the test results, it was assumed that a proportion of FNs may have undergone surgery for suspected malignancy, as indicated by the test results. In ROCkeTS, these surgeries were often carried out due to suspicion of cancer because in ROCkeTS there was no change of care mandated by diagnostic test. Rates of surgery are high because patients were managed by RMI. RMI compensates for poor sensitivity by mandating that almost all patients receive surgery. FN surgery for suspected benign histology and/or symptoms were conducted in secondary care. Further surgery after correct diagnosis were conducted in tertiary care.

Test-specific figures were used to differentiate between TNs and FNs. For FNs, surgery was assumed to be bilateral salpingo-oopherectomy in accordance with the Royal College of Obstetricians and Gynaecologists (RCOG) guidelines[16]. A histology was expected to immediately identify cancer, prompting triage to tertiary care where a staging CT was performed to determine if the cancer was Stage I or Stage II-IV. Stage distributions are derived from ROCkeTS data and are test specific. As with TP cases, patients with missing FIGO stage were treated as Stage IV.

For Stage I FN cases, additional completion surgeries such as total abdominal hysterectomy and omentectomy are performed. These patients either proceed to follow-up or receive six cycles of CarboTaxol and then a follow-up. All Stage I patients are assumed to be disease-free at 12 months and undergo germline testing. Stage II-IV patients are assessed for resectability. Resectable patients (Stage II-III) undergo either partial or extensive surgery, followed by adjuvant chemotherapy (CarboTaxol) and maintenance therapy based on homologous recombination deficiency (HRD) status. Unresectable patients (Stage IV) receive interval debulking surgery after three cycles of CarboTaxol plus Bevacizumab, followed by three additional cycles of CarboTaxol plus Bevacizumab.

During the completion surgery, 23% of Stage I patients are upstaged [14]. Based on the studies included in their meta-analysis, it was assumed that 100% of these upstaged patients moved to stages 2 and 3. These patients undergo extensive surgery, followed by six cycles of CarboTaxol plus Bevacizumab and maintenance therapy (Olaparib plus Bevacizumab) and two follow up appointments. For FNs who were initially discharged to follow-up, it was assumed that suspected cancer is identified during the six-month follow-up with a gynaecologist in secondary care, after which they are triaged to tertiary care for treatment. Survival outcomes are based on the equivalent ones for TP cases due to lack of data on FN survival outcomes.

FN cases diagnosed at the six-month follow up appointment follow the same treatment pathway as TPs, but without maintenance therapy as the first-line treatment takes six months. Additionally, FN cases with ADNEX scores below 1% (Supporting Information Figure S3) [ADNEX diagnostic strategies only] or RMI scores < 25 (Supporting Information Figure S2) [RMI 200 strategy only] were deemed to be at very low risk and are discharged immediately without surgery. These individuals were identified at the six- or twelve-month follow-up and follow the TP treatment pathway.

False Positives

FP cases are managed through either staging CT alone or staging CT combined with additional MRI imaging (Supporting Information Figure S7). 35% of FP cases were assumed to have both a staging CT and MRI imaging patients. For patients who receive only a staging CT, it was assumed that the results suggest Stage I cancer. Consequently, all these patients undergo a staging laparotomy. However, a histology following the procedure reveals a false diagnosis, confirming the absence of cancer. These patients are then discharged and require no further intervention.

In contrast, for those who undergo both staging CT and additional MRI imaging it was assumed that the results are sufficient to rule out cancer, and these patients do not proceed to staging laparotomy. Instead, they were discharged directly and require no further intervention.

True Negatives

TN cases are managed by either undergoing surgery for suspected benign histology and/or symptoms, or by being discharged and followed up with two six-month consultations with a gynaecologist in secondary care (Supporting Information Figure S6). For TNs who undergo surgery, histology confirms the correct diagnosis, and these patients are then discharged without the need for follow up, exiting the model.

For TNs with ADNEX scores < 1% [ADNEX diagnostic strategies only] or RMI scores < 25 [RMI 200 strategy only], it was assumed that they are at very low risk, and thus they are discharged immediately without surgery. These patients are followed up with two six-month consultations in secondary care, involving CA125 testing and CT scans to ensure no progression of symptoms.

##### Pathway Endpoints General Assumptions

Each terminal node, or endpoint, in the model is coded for the three outcome measures accordingly. The proportions of patients reaching each endpoint are derived from the treatment pathways within the model.

For patients with ovarian cancer, the outcome at the end of the 12-month treatment pathway is categorised as either survival or cancer death. For costing purposes, this approach implicitly assumes that death occurs at the 12-month mark, ensuring that full treatment resources are utilised up until these points. For patients without ovarian cancer, the model ends with either discharge or follow-up care, reflecting the completion of their pathway.

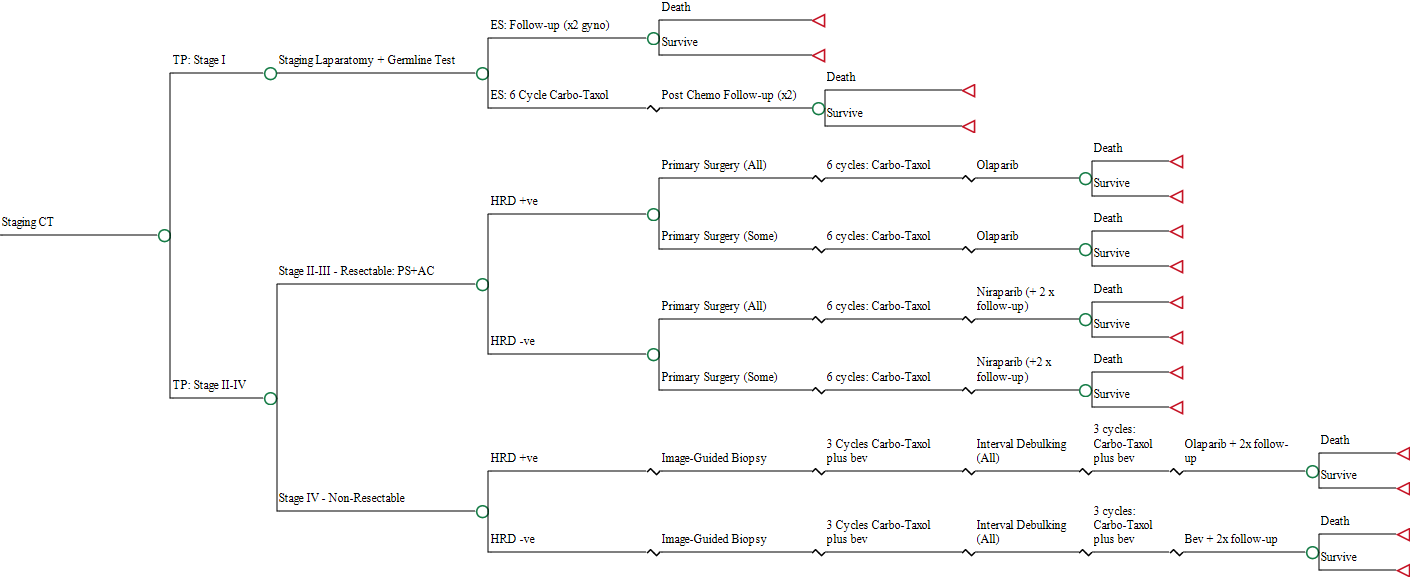

#### **Supporting Information Figure S1 True Positive Pathway**

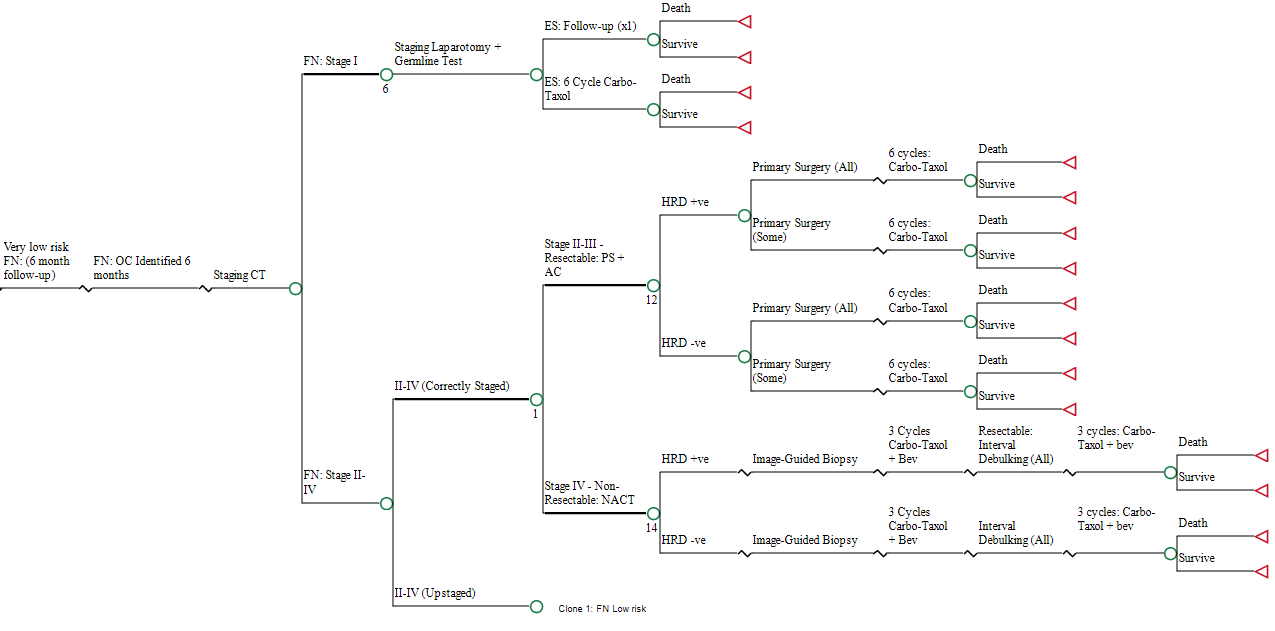

#### **Supporting Information Figure S2 False Negative Pathway for very low risk patients in RMI 200 strategy**

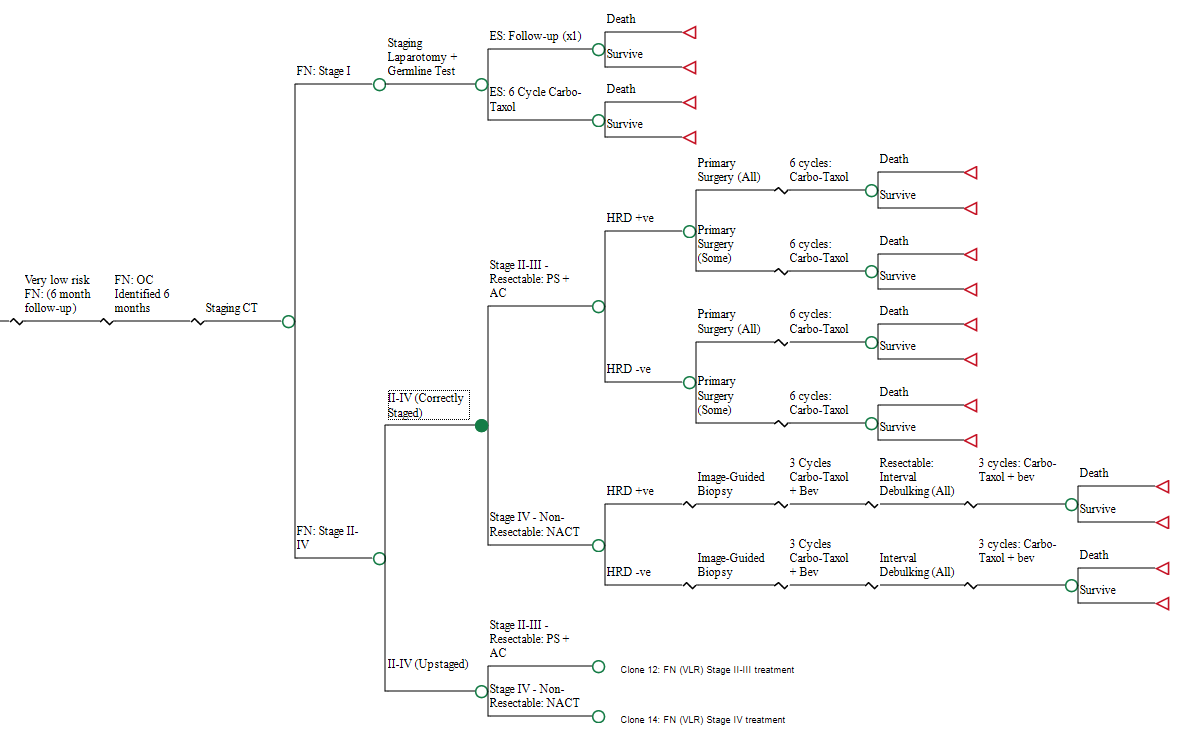

#### **Supporting Information Figure S3 False Negative Pathway for very low risk patients in ADNEX strategies**

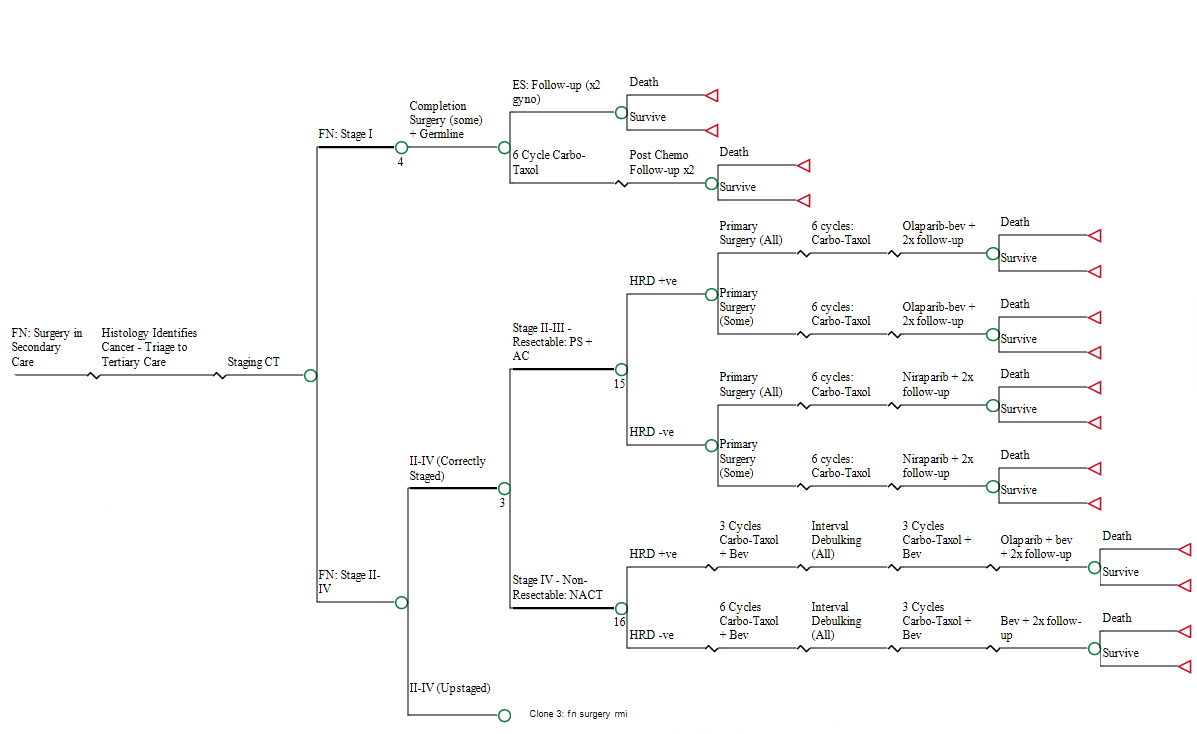

#### **Supporting Information Figure S4 Pathway for False Negative patients who have surgery.**

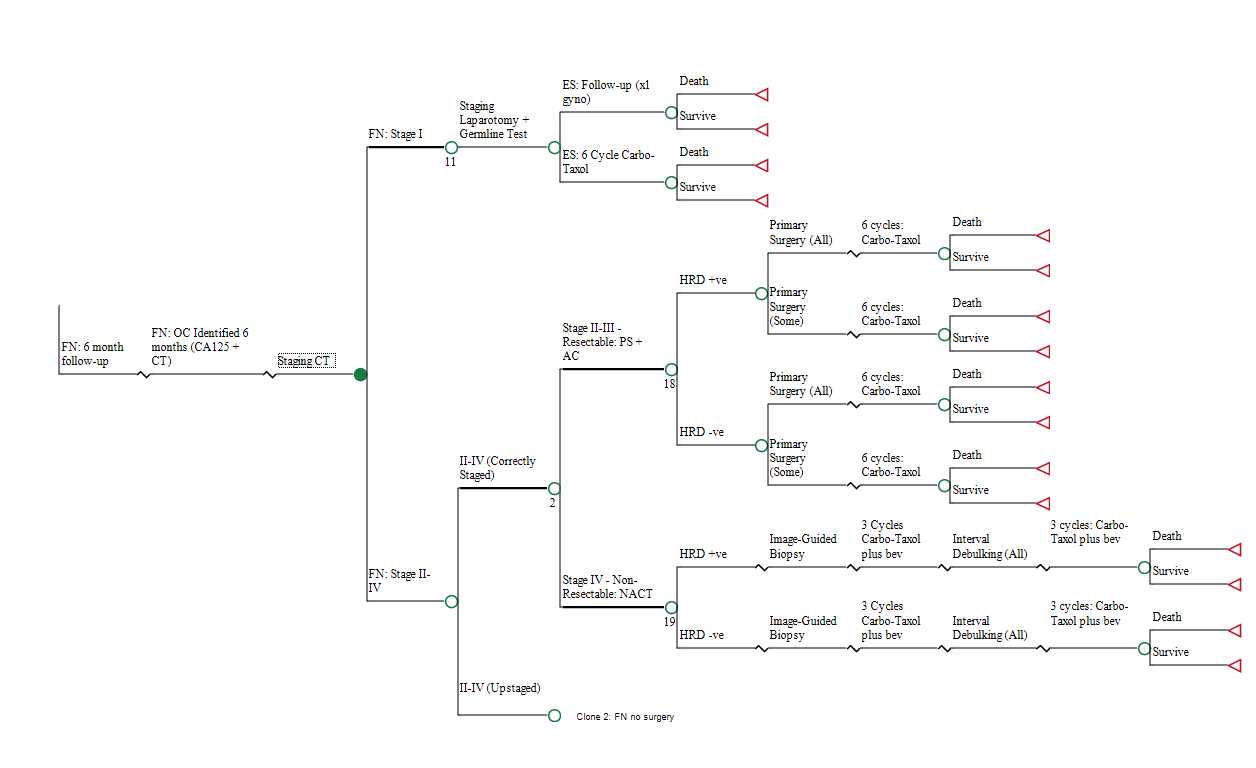

#### **Supporting Information Figure S5 Pathway for False Negative patients who have 6 months follow up**

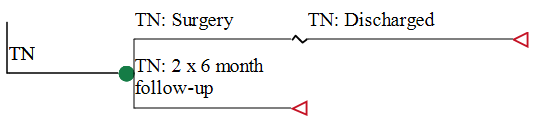

#### **Supporting Information Figure S6 True Negative Pathway**

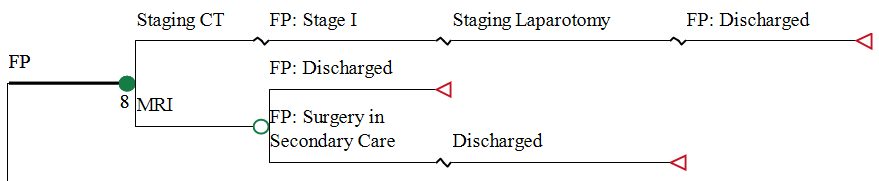

#### **Supporting Information Figure S7 False Positive Pathway**

### **Table S4 Deterministic Sensitivity Analysis output premenopausal cohort**

| ***Proportion of Ovarian Cancer changed to 0.0460*** | | | | |
| --- | --- | --- | --- | --- |
| **Strategy** | **Cost** | **Cancer deaths** | **Proportion of correct diagnosis** | **Diagnostic Yield** |
| RMI 200 | £4,483 | 0.88% | 93.22% | 2.25% |
| ADNEX 10% | £6,118 | 0.80% | 75.78% | 4.10% |
| ORADS | £6,237 | 0.84% | 84.68% | 3.64% |
| CA125 | £6,815 | 1.08% | 64.95% | 3.57% |
| ADNEX 3% | £7,120 | 0.79% | 47.74% | 4.30% |
| ROMA 11.4% | £7,305 | 0.85% | 73.36% | 3.64% |
| ***Proportion of Ovarian Cancer changed to 0.0767*** | | | | |
| **Strategy** | **Cost** | **Cancer deaths** | **Proportion of correct diagnosis** | **Diagnostic Yield** |
| RMI 200 | £6,083 | 1.47% | 91.80% | 3.75% |
| ADNEX 10% | £7,528 | 1.33% | 76.20% | 6.83% |
| ORADS | £7,691 | 1.40% | 84.50% | 6.07% |
| CA125 | £8,403 | 1.80% | 65.36% | 5.94% |
| ADNEX 3% | £8,469 | 1.32% | 49.21% | 7.17% |
| ROMA 11.4% | £8,806 | 1.42% | 73.54% | 6.07% |
| ***The lower Confidence Interval of the upstaging variable was used (15.5%)*** | | | | |
| **Strategy** | **Cost** | **Cancer deaths** | **Proportion of correct diagnosis** | **Diagnostic Yield** |
| RMI 200 | £5,188 | 1.15% | 92.51% | 3.00% |
| ADNEX 10% | £6,808 | 1.06% | 75.99% | 5.47% |
| ORADS | £6,937 | 1.11% | 84.59% | 4.86% |
| CA125 | £7,563 | 1.42% | 65.15% | 4.76% |
| ADNEX 3% | £7,795 | 1.05% | 48.47% | 5.73% |
| ROMA 11.4% | £8,005 | 1.12% | 73.45% | 4.86% |
| ***The upper Confidence Interval of the upstaging variable was used (30.5%)*** | | | | |
| **Strategy** | **Cost** | **Cancer deaths** | **Proportion of correct diagnosis** | **Diagnostic Yield** |
| RMI 200 | £5,378 | 1.21% | 92.51% | 3.00% |
| ADNEX 10% | £6,837 | 1.07% | 75.99% | 5.47% |
| ORADS | £6,991 | 1.13% | 84.59% | 4.86% |
| CA125 | £7,655 | 1.45% | 65.15% | 4.76% |
| ADNEX 3% | £7,795 | 1.05% | 48.47% | 5.73% |
| ROMA 11.4% | £8,106 | 1.15% | 73.45% | 4.86% |
| ***Proportion of False Positives that have additional MRI was varied to 0.2625*** | | | | |
| **Strategy** | **Cost** | **Cancer deaths** | **Proportion of correct diagnosis** | **Diagnostic Yield** |
| RMI 200 | £5,292 | 1.18% | 92.51% | 3.00% |
| ADNEX 10% | £6,865 | 1.07% | 75.99% | 5.47% |
| ORADS | £6,989 | 1.12% | 84.59% | 4.86% |
| CA125 | £7,699 | 1.44% | 65.15% | 4.76% |
| ADNEX 3% | £7,901 | 1.05% | 48.47% | 5.73% |
| ROMA 11.4% | £8,129 | 1.14% | 73.45% | 4.86% |
| ***Proportion of False Positives that have additional MRI was varied to 0.4375*** | | | | |
| **Strategy** | **Cost** | **Cancer deaths** | **Proportion of correct diagnosis** | **Diagnostic Yield** |
| RMI 200 | £5,274 | 1.18% | 92.51% | 3.00% |
| ADNEX 10% | £6,780 | 1.07% | 75.99% | 5.47% |
| ORADS | £6,939 | 1.12% | 84.59% | 4.86% |
| CA125 | £7,519 | 1.44% | 65.15% | 4.76% |
| ADNEX 3% | £7,688 | 1.05% | 48.47% | 5.73% |
| ROMA 11.4% | £7,982 | 1.14% | 73.45% | 4.86% |
| ***Proportion of True positives with Stage I Ovarian Cancer was reduced by 25%*** | | | | |
| **Strategy** | **Cost** | **Cancer deaths** | **Proportion of correct diagnosis** | **Diagnostic Yield** |
| RMI 200 | £5,407 | 1.22% | 92.51% | 3.00% |
| ADNEX 10% | £7,202 | 1.21% | 75.99% | 5.47% |
| ORADS | £7,293 | 1.26% | 84.59% | 4.86% |
| CA125 | £7,889 | 1.58% | 65.15% | 4.76% |
| ADNEX 3% | £8,217 | 1.22% | 48.47% | 5.73% |
| ROMA 11.4% | £8,320 | 1.24% | 73.45% | 4.86% |
| ***Proportion of True positives with Stage I Ovarian Cancer was increased by 25%*** | | | | |
| **Strategy** | **Cost** | **Cancer deaths** | **Proportion of correct diagnosis** | **Diagnostic Yield** |
| RMI 200 | £5,159 | 1.13% | 92.51% | 3.00% |
| ADNEX 10% | £6,443 | 0.92% | 75.99% | 5.47% |
| ORADS | £6,634 | 0.98% | 84.59% | 4.86% |
| CA125 | £7,328 | 1.30% | 65.15% | 4.76% |
| ADNEX 3% | £7,373 | 0.89% | 48.47% | 5.73% |
| ROMA 11.4% | £7,791 | 1.03% | 73.45% | 4.86% |
| ***Proportion of correctly staged resectable patients who have all listed surgery procedures was varied to 0.2625*** | | | | |
| **Strategy** | **Cost** | **Cancer deaths** | **Proportion of correct diagnosis** | **Diagnostic Yield** |
| RMI 200 | £5,091 | 1.18% | 92.51% | 3.00% |
| ADNEX 10% | £6,725 | 1.07% | 75.99% | 5.47% |
| ORADS | £6,851 | 1.12% | 84.59% | 4.86% |
| CA125 | £7,516 | 1.44% | 65.15% | 4.76% |
| ADNEX 3% | £7,714 | 1.05% | 48.47% | 5.73% |
| ROMA 11.4% | £7,919 | 1.14% | 73.45% | 4.86% |
| ***Proportion of correctly staged resectable patients who have all listed surgery procedures was varied to 0.4375*** | | | | |
| **Strategy** | **Cost** | **Cancer deaths** | **Proportion of correct diagnosis** | **Diagnostic Yield** |
| RMI 200 | £5,276 | 1.18% | 92.51% | 3.00% |
| ADNEX 10% | £6,891 | 1.07% | 75.99% | 5.47% |
| ORADS | £7,021 | 1.12% | 84.59% | 4.86% |
| CA125 | £7,606 | 1.44% | 65.15% | 4.76% |
| ADNEX 3% | £7,876 | 1.05% | 48.47% | 5.73% |
| ROMA 11.4% | £8,095 | 1.14% | 73.45% | 4.86% |
| ***For the ADNEX 10 strategy, the sensitivity and specificity of a two-step strategy was used*** | | | | |
| **Strategy** | **Cost** | **Cancer deaths** | **Proportion of correct diagnosis** | **Diagnostic Yield** |
| RMI 200 | £5,283 | 1.18% | 92.51% | 3.00% |
| ADNEX 10% | £6,372 | 1.05% | 84.90% | 5.59% |
| ORADS | £6,964 | 1.12% | 84.59% | 4.86% |
| CA125 | £7,609 | 1.44% | 65.15% | 4.76% |
| ADNEX 3% | £7,795 | 1.05% | 48.47% | 5.73% |
| ROMA 11.4% | £8,056 | 1.14% | 73.45% | 4.86% |

### **Table S5 Deterministic Sensitivity Analysis output postmenopausal cohort**

| ***Proportion of Ovarian Cancer changed to 0.1494*** | | | | |
| --- | --- | --- | --- | --- |
| **Strategy** | **Cost** | **Cancer deaths** | **Proportion of correct diagnosis** | **Diagnostic Yield** |
| RMI 200 | £10,810 | 2.88% | 84.49% | 12.70% |
| ORADS | £11,603 | 3.01% | 78.02% | 11.42% |
| CA125 | £11,694 | 3.03% | 78.86% | 12.85% |
| ROMA 29.9% | £11,799 | 2.97% | 81.11% | 13.15% |
| ADNEX 10% | £11,915 | 2.80% | 64.12% | 14.36% |
| ADNEX 3% | £12,629 | 2.78% | 41.14% | 14.94% |
| ***Proportion of Ovarian Cancer changed to 0.2491*** | | | | |
| **Strategy** | **Cost** | **Cancer deaths** | **Proportion of correct diagnosis** | **Diagnostic Yield** |
| RMI 200 | £16,132 | 4.80% | 84.55% | 21.17% |
| ORADS | £16,884 | 5.02% | 77.83% | 19.03% |
| ADNEX 10% | £17,027 | 4.66% | 67.86% | 23.94% |
| CA125 | £17,135 | 5.05% | 79.69% | 21.42% |
| ROMA 29.9% | £17,188 | 4.96% | 81.92% | 21.92% |
| ADNEX 3% | £17,609 | 4.63% | 48.04% | 24.91% |
| ***The lower Confidence Interval of the upstaging variable was used (15.5%)*** | | | | |
| **Strategy** | **Cost** | **Cancer deaths** | **Proportion of correct diagnosis** | **Diagnostic Yield** |
| RMI 200 | £13,386 | 3.81% | 84.52% | 16.94% |
| ORADS | £14,185 | 4.00% | 77.92% | 15.22% |
| CA125 | £14,333 | 4.01% | 79.27% | 17.14% |
| ROMA 29.9% | £14,408 | 3.94% | 81.51% | 17.53% |
| ADNEX 10% | £14,443 | 3.72% | 65.99% | 19.15% |
| ADNEX 3% | £15,119 | 3.70% | 44.59% | 19.93% |
| ***The upper Confidence Interval of the upstaging variable was used (30.5%)*** | | | | |
| **Strategy** | **Cost** | **Cancer deaths** | **Proportion of correct diagnosis** | **Diagnostic Yield** |
| RMI 200 | £13,556 | 3.87% | 84.52% | 16.94% |
| ORADS | £14,302 | 4.04% | 77.92% | 15.22% |
| CA125 | £14,496 | 4.06% | 79.27% | 17.14% |
| ADNEX 10% | £14,500 | 3.74% | 65.99% | 19.15% |
| ROMA 29.9% | £14,579 | 3.99% | 81.51% | 17.53% |
| ADNEX 3% | £15,119 | 3.70% | 44.59% | 19.93% |
| ***Proportion of False Positives that have additional MRI was varied to 0.2625*** | | | | |
| **Strategy** | **Cost** | **Cancer deaths** | **Proportion of correct diagnosis** | **Diagnostic Yield** |
| RMI 200 | £13,529 | 3.84% | 84.52% | 16.94% |
| ORADS | £14,321 | 4.02% | 77.92% | 15.22% |
| CA125 | £14,499 | 4.04% | 79.27% | 17.14% |
| ROMA 29.9% | £14,573 | 3.96% | 81.51% | 17.53% |
| ADNEX 10% | £14,605 | 3.73% | 65.99% | 19.15% |
| ADNEX 3% | £15,335 | 3.70% | 44.59% | 19.93% |
| ***Proportion of False Positives that have additional MRI was varied to 0.4375*** | | | | |
| **Strategy** | **Cost** | **Cancer deaths** | **Proportion of correct diagnosis** | **Diagnostic Yield** |
| RMI 200 | £13,413 | 3.84% | 84.52% | 16.94% |
| ORADS | £14,166 | 4.02% | 77.92% | 15.22% |
| CA125 | £14,329 | 4.04% | 79.27% | 17.14% |
| ADNEX 10% | £14,338 | 3.73% | 65.99% | 19.15% |
| ROMA 29.9% | £14,414 | 3.96% | 81.51% | 17.53% |
| ADNEX 3% | £14,904 | 3.70% | 44.59% | 19.93% |
| ***Proportion of True positives with Stage I Ovarian Cancer was reduced by 25%*** | | | | |
| **Strategy** | **Cost** | **Cancer deaths** | **Proportion of correct diagnosis** | **Diagnostic Yield** |
| RMI 200 | £14,250 | 4.15% | 84.52% | 16.94% |
| ORADS | £15,018 | 4.32% | 77.92% | 15.22% |
| CA125 | £15,127 | 4.32% | 79.27% | 17.14% |
| ROMA 29.9% | £15,242 | 4.26% | 81.51% | 17.53% |
| ADNEX 10% | £15,453 | 4.11% | 65.99% | 19.15% |
| ADNEX 3% | £16,190 | 4.12% | 44.59% | 19.93% |
| ***Proportion of True positives with Stage I Ovarian Cancer was increased by 25%*** | | | | |
| **Strategy** | **Cost** | **Cancer deaths** | **Proportion of correct diagnosis** | **Diagnostic Yield** |
| RMI 200 | £12,693 | 3.53% | 84.52% | 16.94% |
| ORADS | £13,469 | 3.72% | 77.92% | 15.22% |
| ADNEX 10% | £13,489 | 3.35% | 65.99% | 19.15% |
| CA125 | £13,702 | 3.75% | 79.27% | 17.14% |
| ROMA 29.9% | £13,745 | 3.67% | 81.51% | 17.53% |
| ADNEX 3% | £14,048 | 3.28% | 44.59% | 19.93% |
| ***Proportion of correctly staged resectable patients who have all listed surgery procedures was varied to 0.2625*** | | | | |
| **Strategy** | **Cost** | **Cancer deaths** | **Proportion of correct diagnosis** | **Diagnostic Yield** |
| RMI 200 | £13,022 | 3.84% | 84.52% | 16.94% |
| ORADS | £13,824 | 4.02% | 77.92% | 15.22% |
| CA125 | £13,969 | 4.04% | 79.27% | 17.14% |
| ROMA 29.9% | £14,051 | 3.96% | 81.51% | 17.53% |
| ADNEX 10% | £14,095 | 3.73% | 65.99% | 19.15% |
| ADNEX 3% | £14,777 | 3.70% | 44.59% | 19.93% |
| ***Proportion of correctly staged resectable patients who have all listed surgery procedures was varied to 0.4375*** | | | | |
| **Strategy** | **Cost** | **Cancer deaths** | **Proportion of correct diagnosis** | **Diagnostic Yield** |
| RMI 200 | £13,734 | 3.84% | 84.52% | 16.94% |
| ORADS | £14,537 | 4.02% | 77.92% | 15.22% |
| CA125 | £14,687 | 4.04% | 79.27% | 17.14% |
| ROMA 29.9% | £14,755 | 3.96% | 81.51% | 17.53% |
| ADNEX 10% | £14,787 | 3.73% | 65.99% | 19.15% |
| ADNEX 3% | £15,462 | 3.70% | 44.59% | 19.93% |
| ***For the ADNEX 10 strategy, the sensitivity and specificity of a two-step strategy was used*** | | | | |
| **Strategy** | **Cost** | **Cancer deaths** | **Proportion of correct diagnosis** | **Diagnostic Yield** |
| RMI 200 | £13,471 | 3.84% | 84.52% | 16.94% |
| ADNEX 10% | £13,524 | 3.69% | 85.82% | 18.15% |
| ORADS | £14,243 | 4.02% | 77.92% | 15.22% |
| CA125 | £14,414 | 4.04% | 79.27% | 17.14% |
| ROMA 29.9% | £14,494 | 3.96% | 81.51% | 17.53% |
| ADNEX 3% | £15,119 | 3.70% | 44.59% | 19.93% |

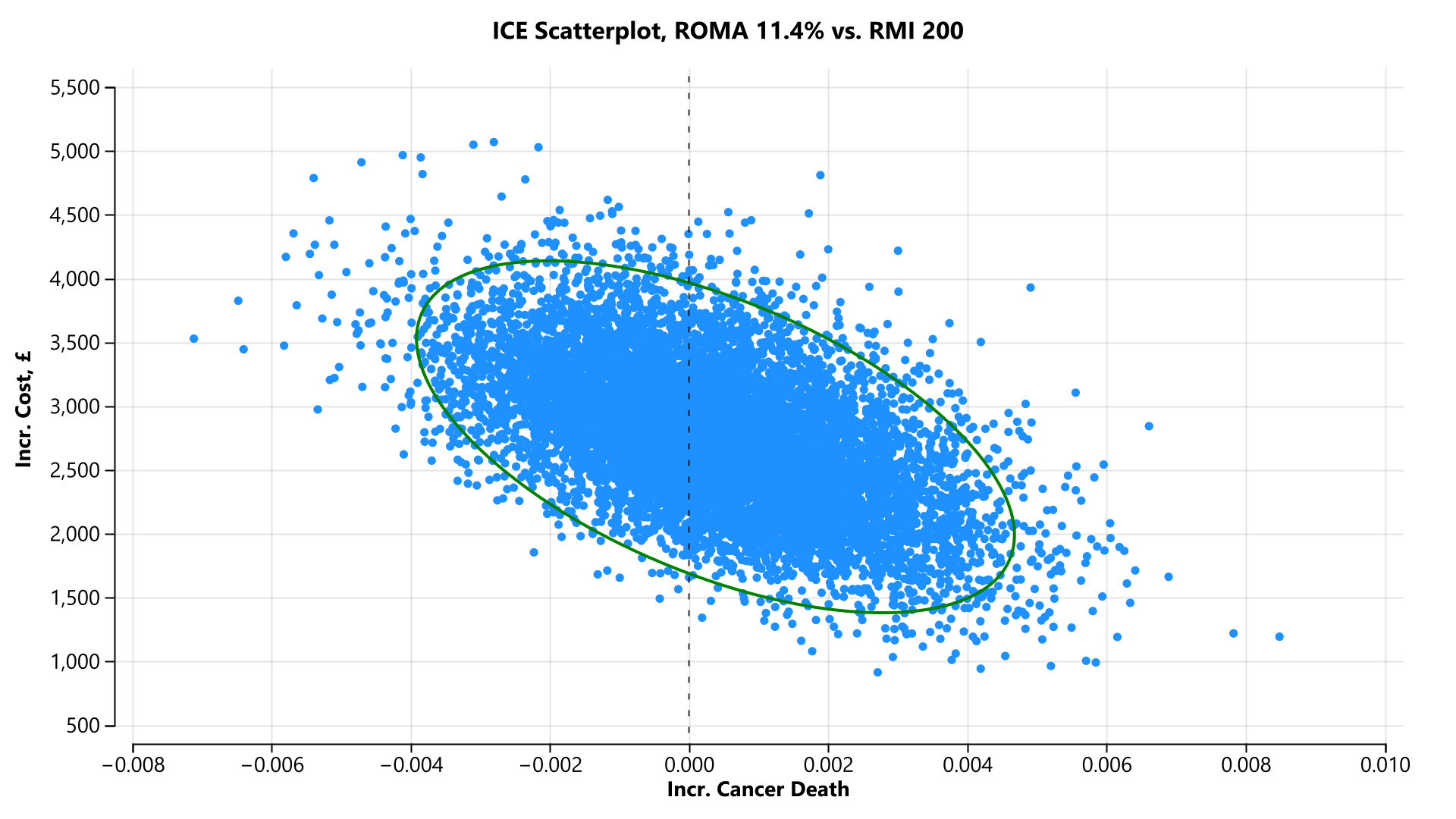

#### **Supporting Information Figure S8 Pre-menopausal cohort incremental cost-effectiveness scatterplot (ROMA 11.4% versus RMI 200) [cancer death]**

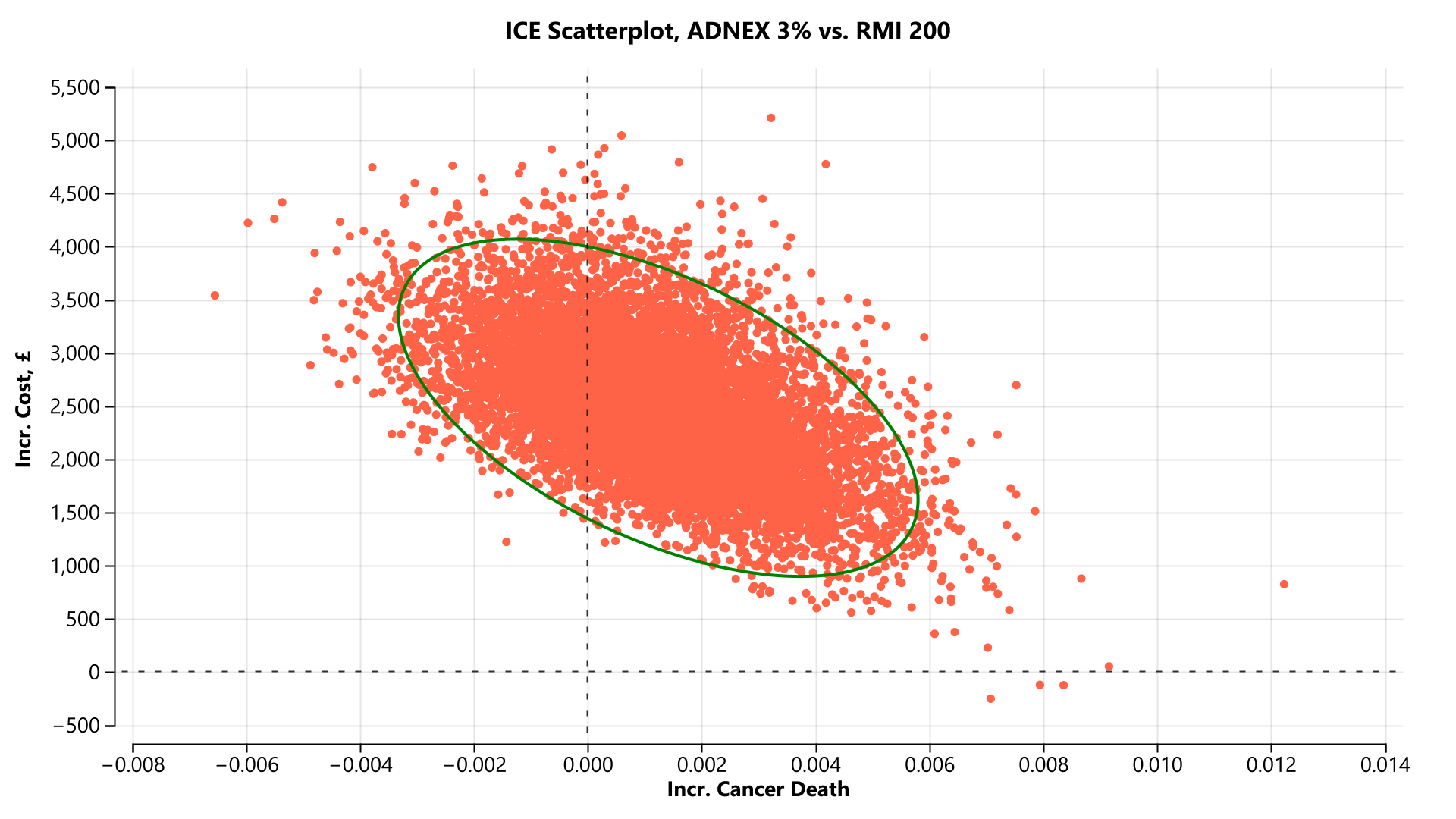

#### **Supporting Information Figure S9 Pre-menopausal cohort incremental cost-effectiveness scatterplot (ADNEX 3% versus RMI 200) [cancer death]**

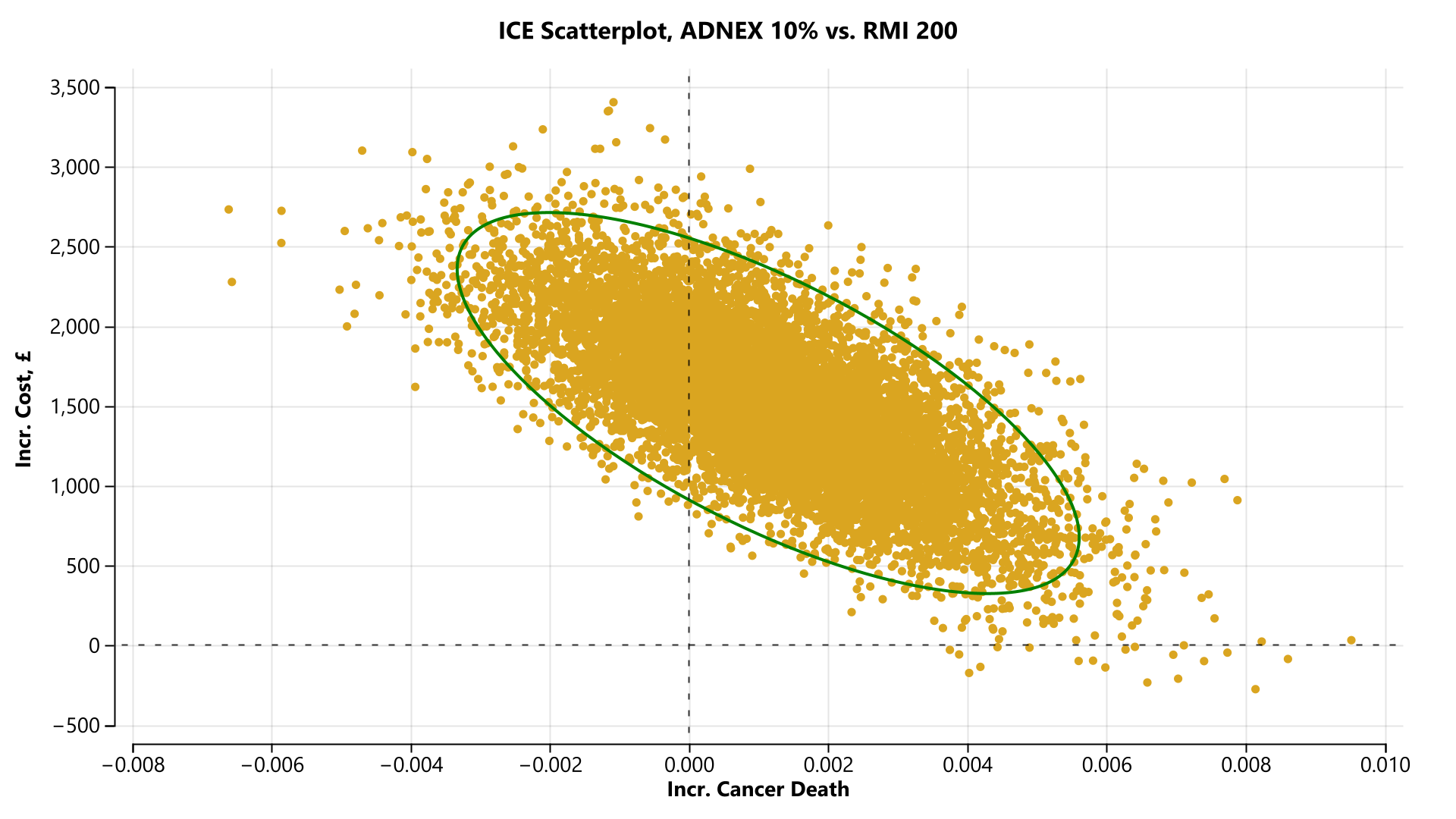

#### **Supporting Information Figure S10 Pre-menopausal cohort incremental cost-effectiveness scatterplot (ADNEX 10% versus RMI 200) [cancer death]**

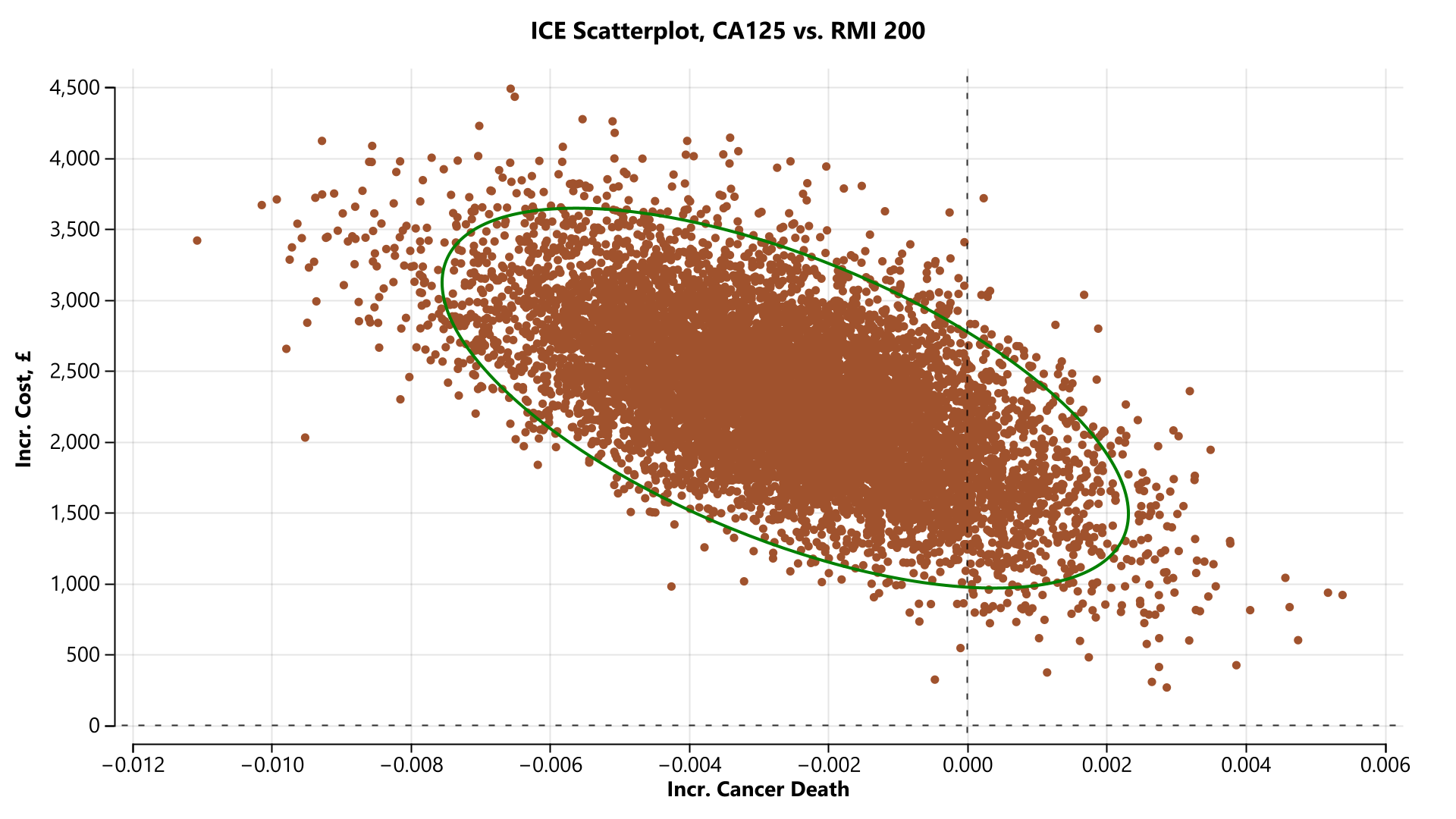

#### **Supporting Information Figure S11 Pre-menopausal cohort incremental cost-effectiveness scatterplot (CA 125 versus RMI 200) [cancer death]**

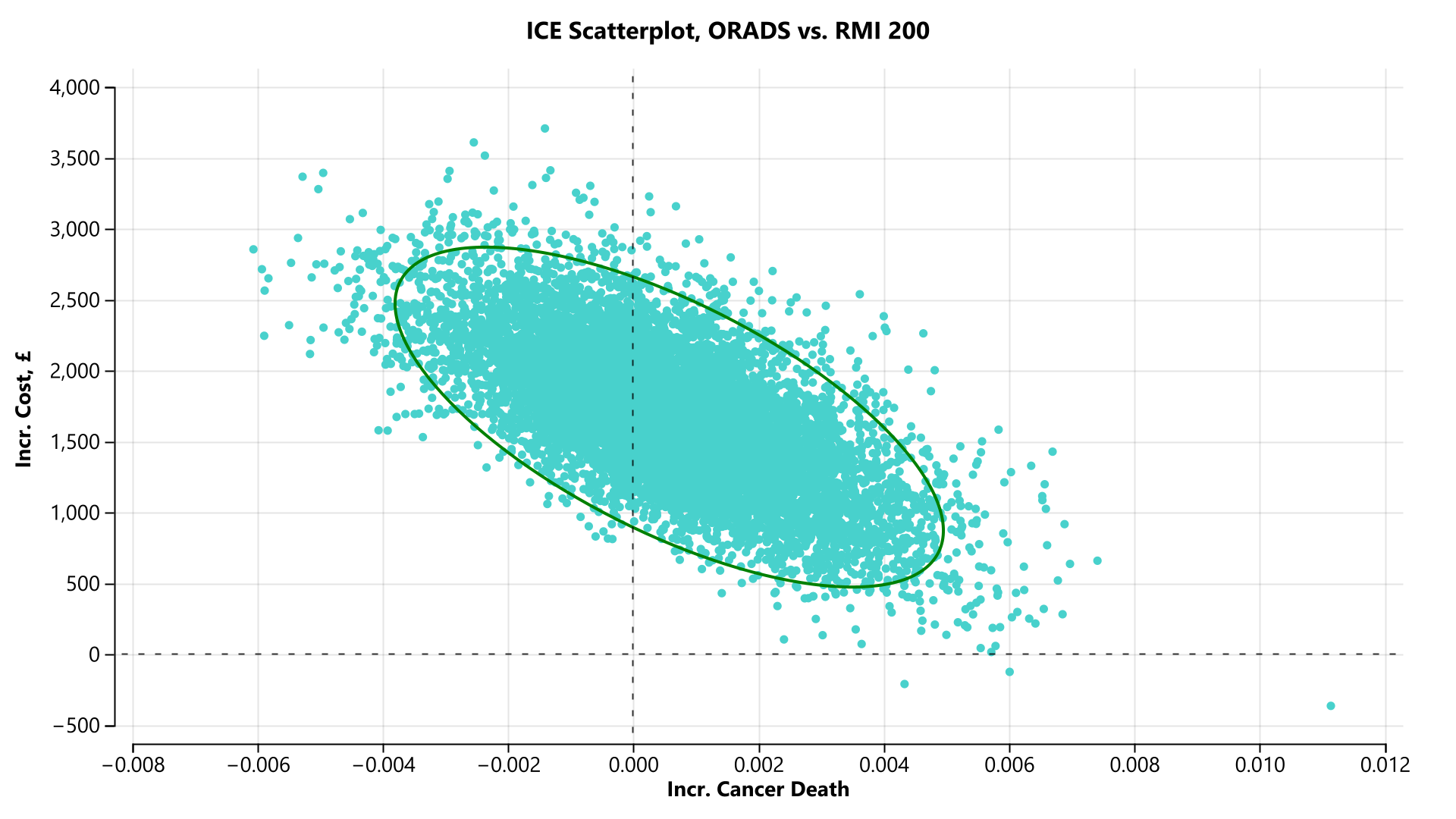

#### **Supporting Information Figure S12 Pre-menopausal cohort incremental cost-effectiveness scatterplot (ORADS versus RMI 200) [cancer death]**

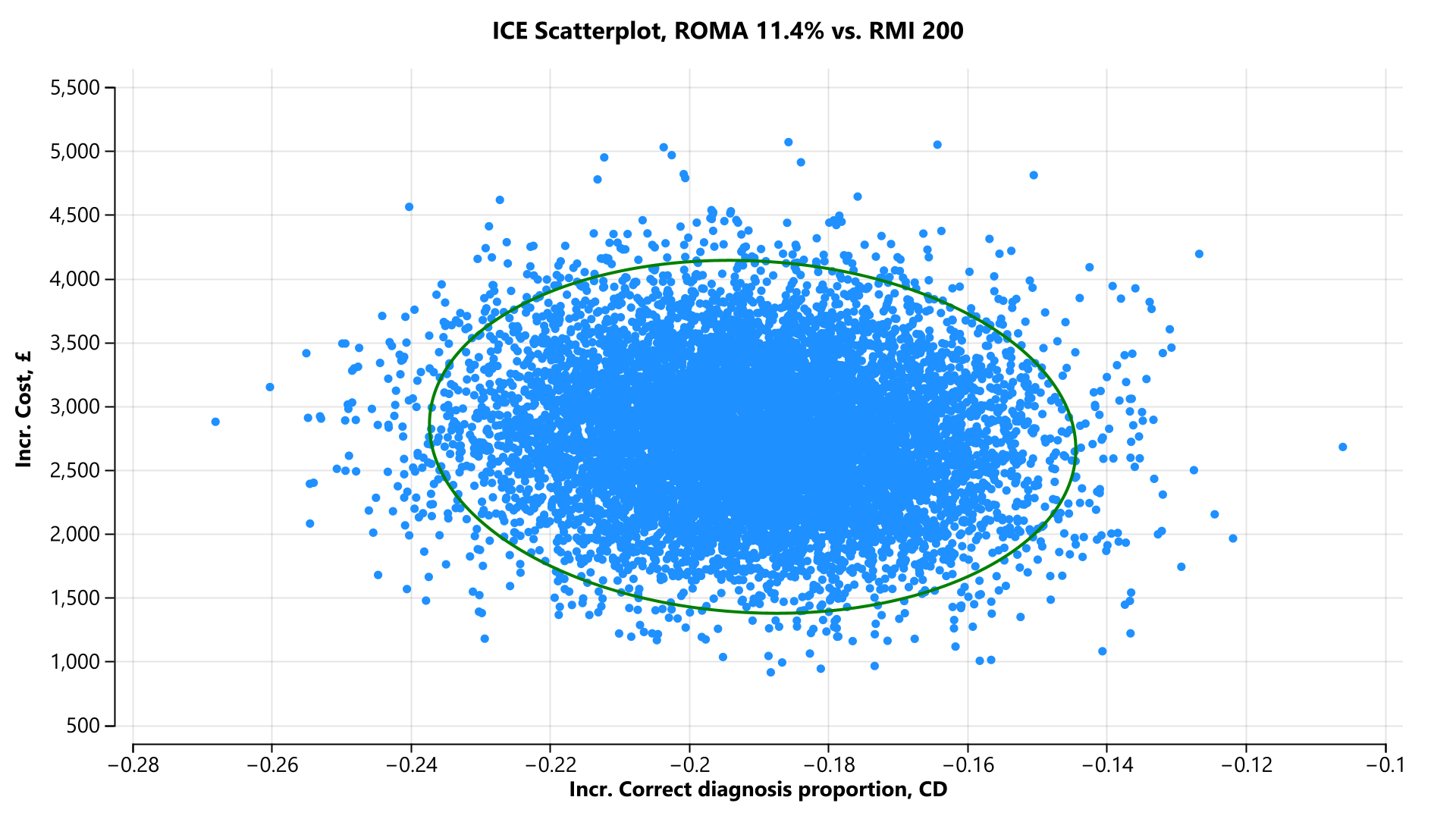
 **Supporting Information Figure S13 Pre-menopausal cohort incremental cost-effectiveness scatterplot (ROMA 11.4% versus RMI 200) [correct diagnosis proportion]**

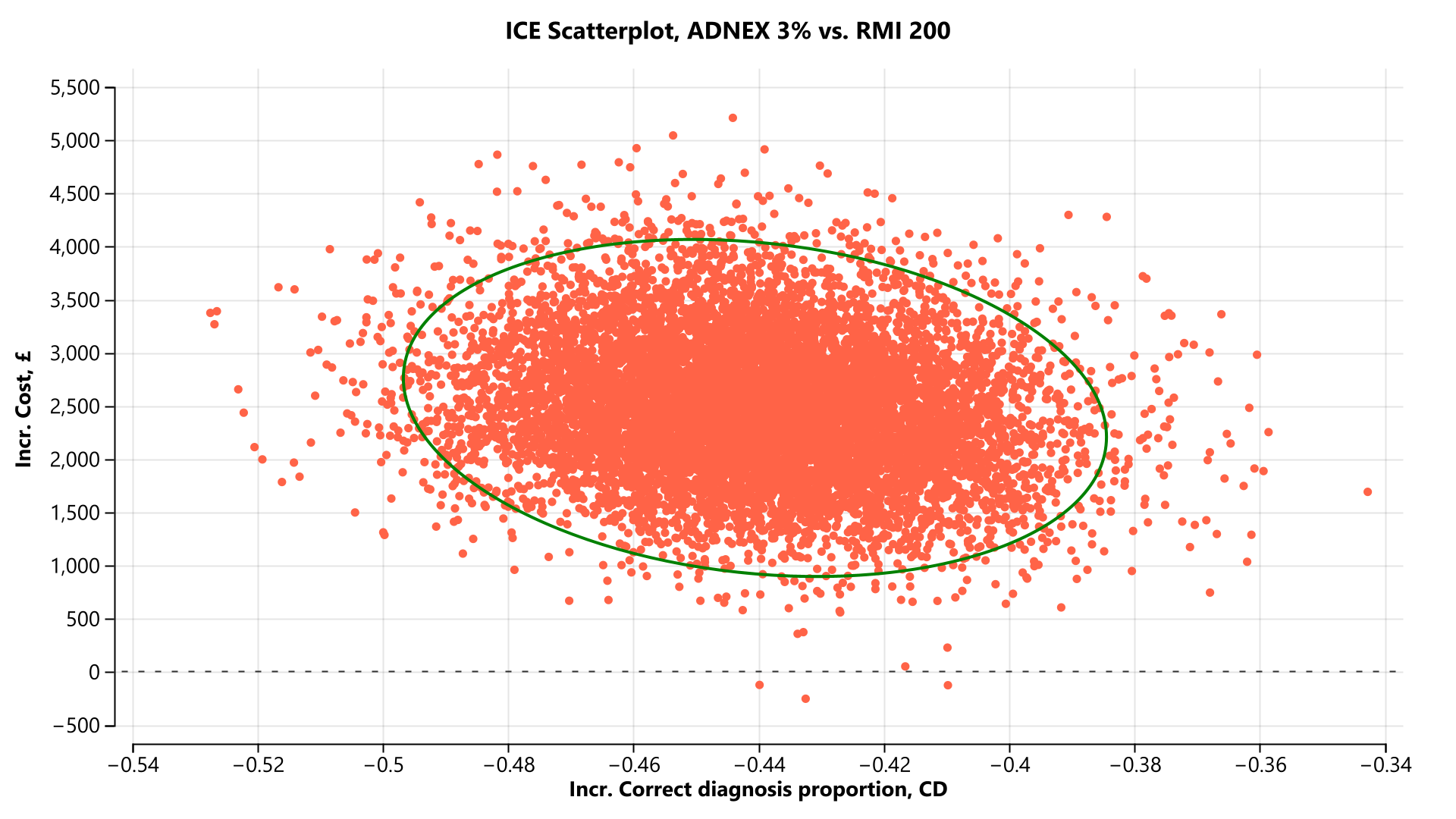
 **Supporting Information Figure S14 Pre-menopausal cohort incremental cost-effectiveness scatterplot (ADNEX 3% versus RMI 200) [correct diagnosis proportion]**

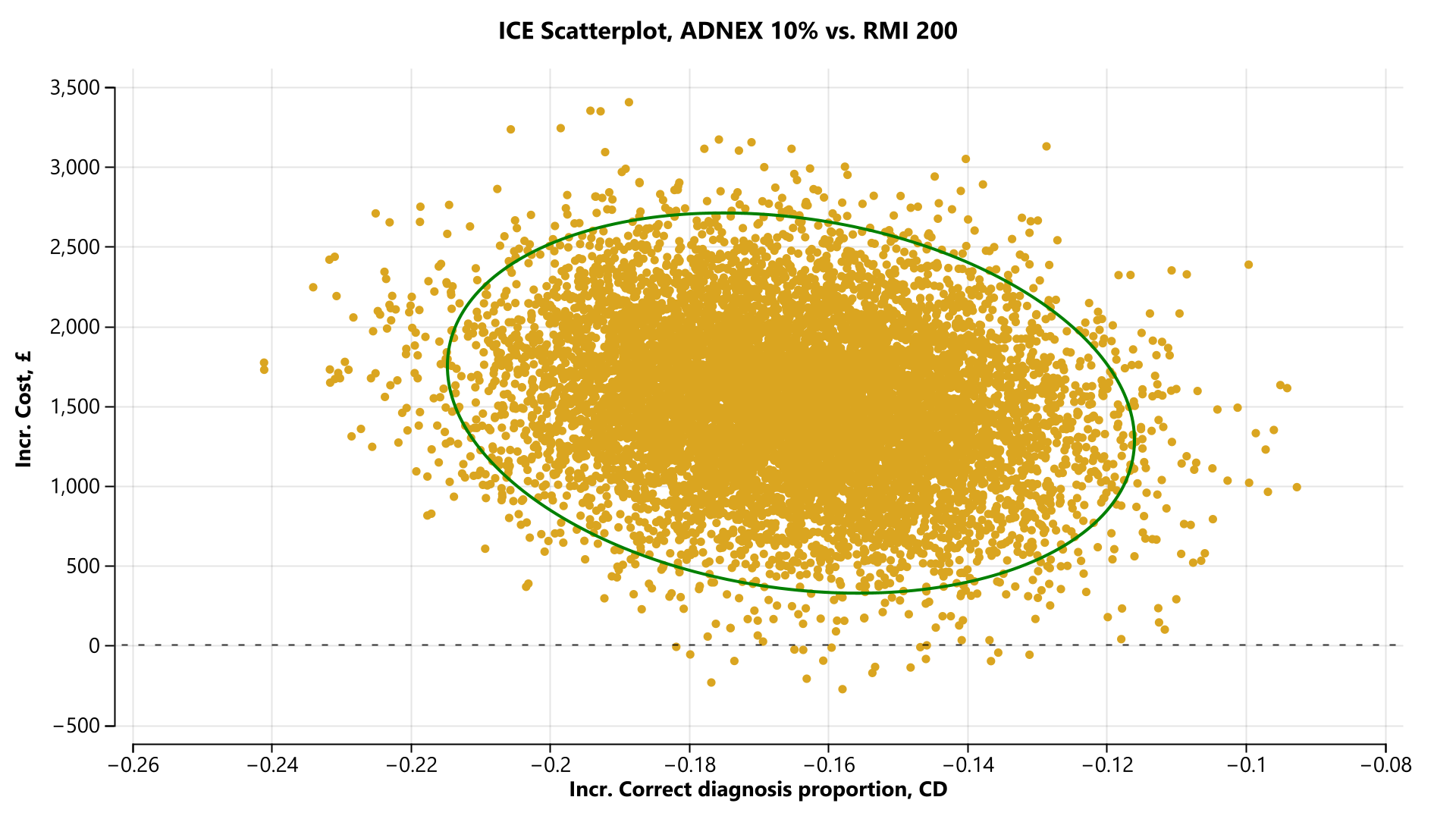
 **Supporting Information Figure S15 Pre-menopausal cohort incremental cost-effectiveness scatterplot (ADNEX 10% versus RMI 200) [correct diagnosis proportion]**

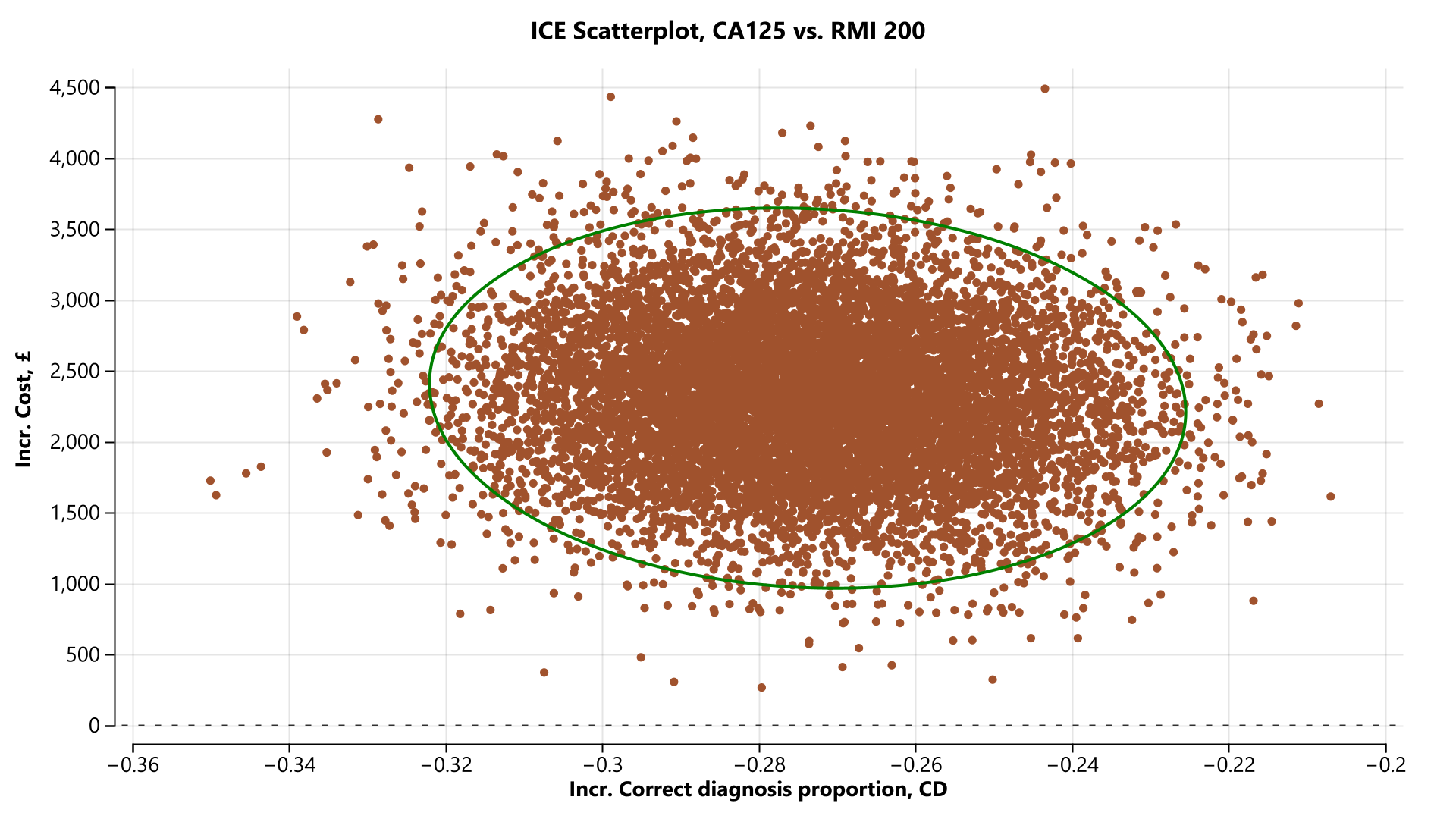
 **Supporting Information Figure S16 Pre-menopausal cohort incremental cost-effectiveness scatterplot (CA 125 versus RMI 200) [correct diagnosis proportion]**

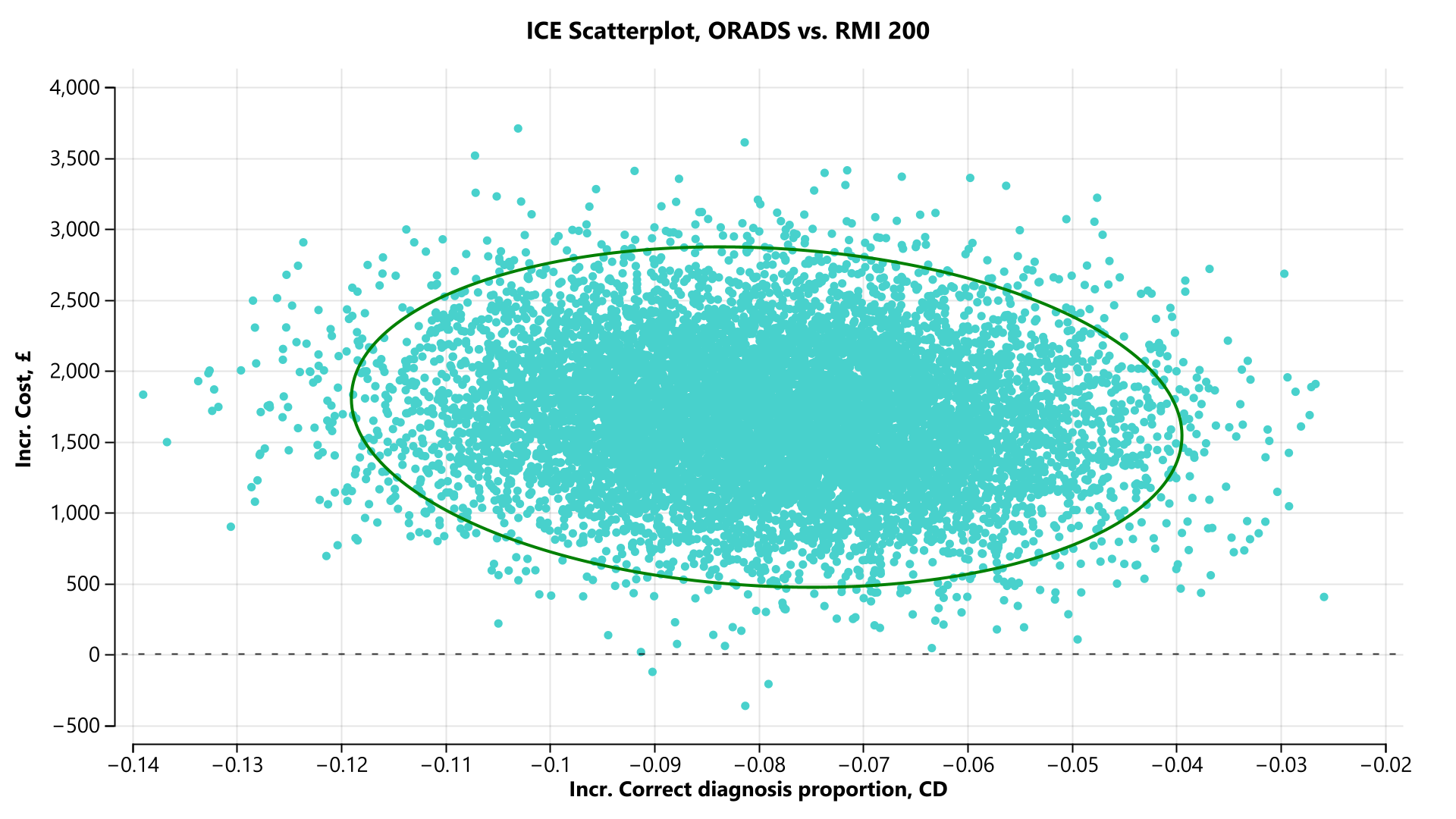

#### **Supporting Information Figure S17 Pre-menopausal cohort incremental cost-effectiveness scatterplot (ORADS versus RMI 200) [correct diagnosis proportion]**

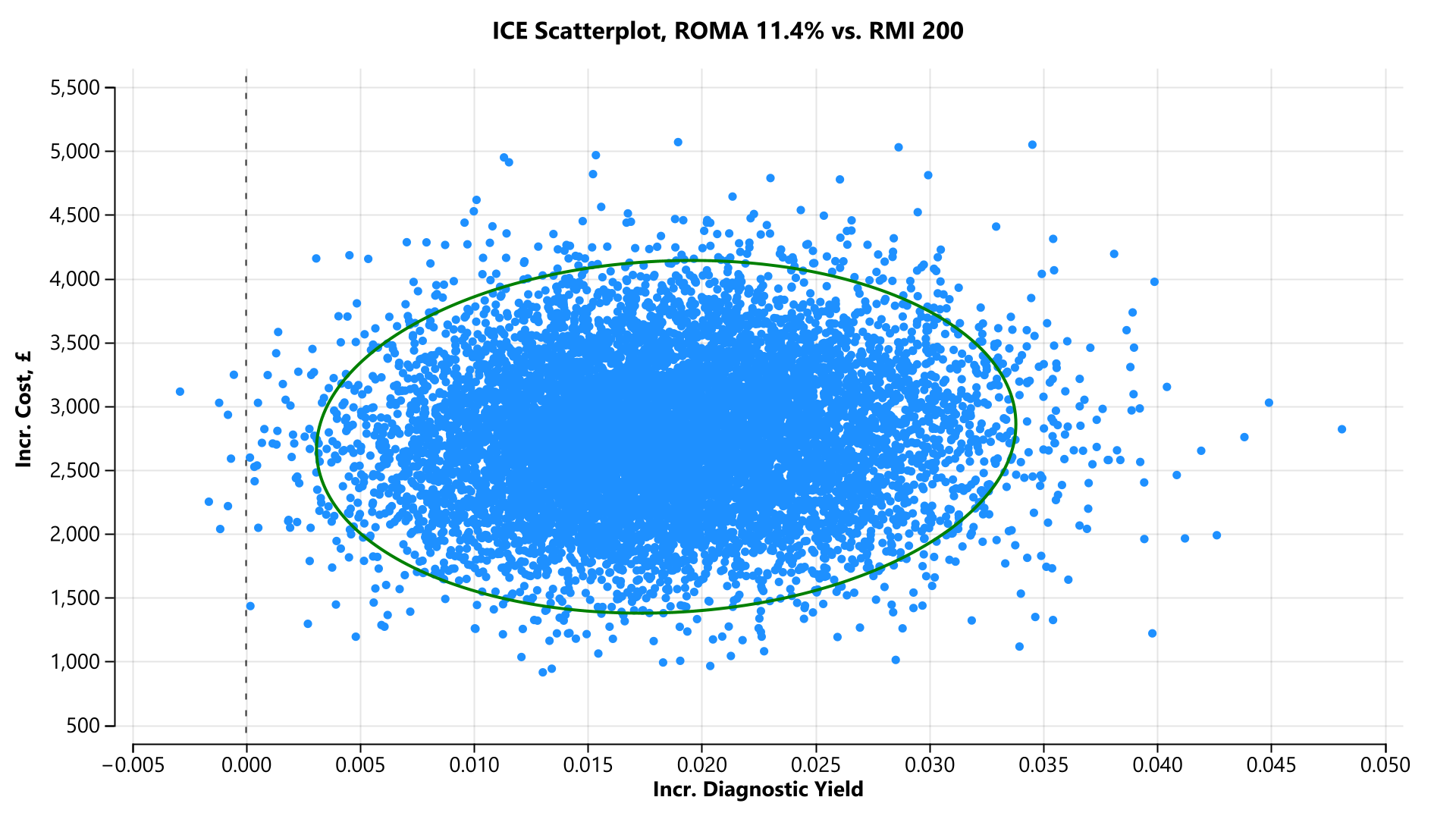
 **Supporting Information Figure S18 Pre-menopausal cohort incremental cost-effectiveness scatterplot (ROMA 11.4% versus RMI 200) [diagnostic yield]**

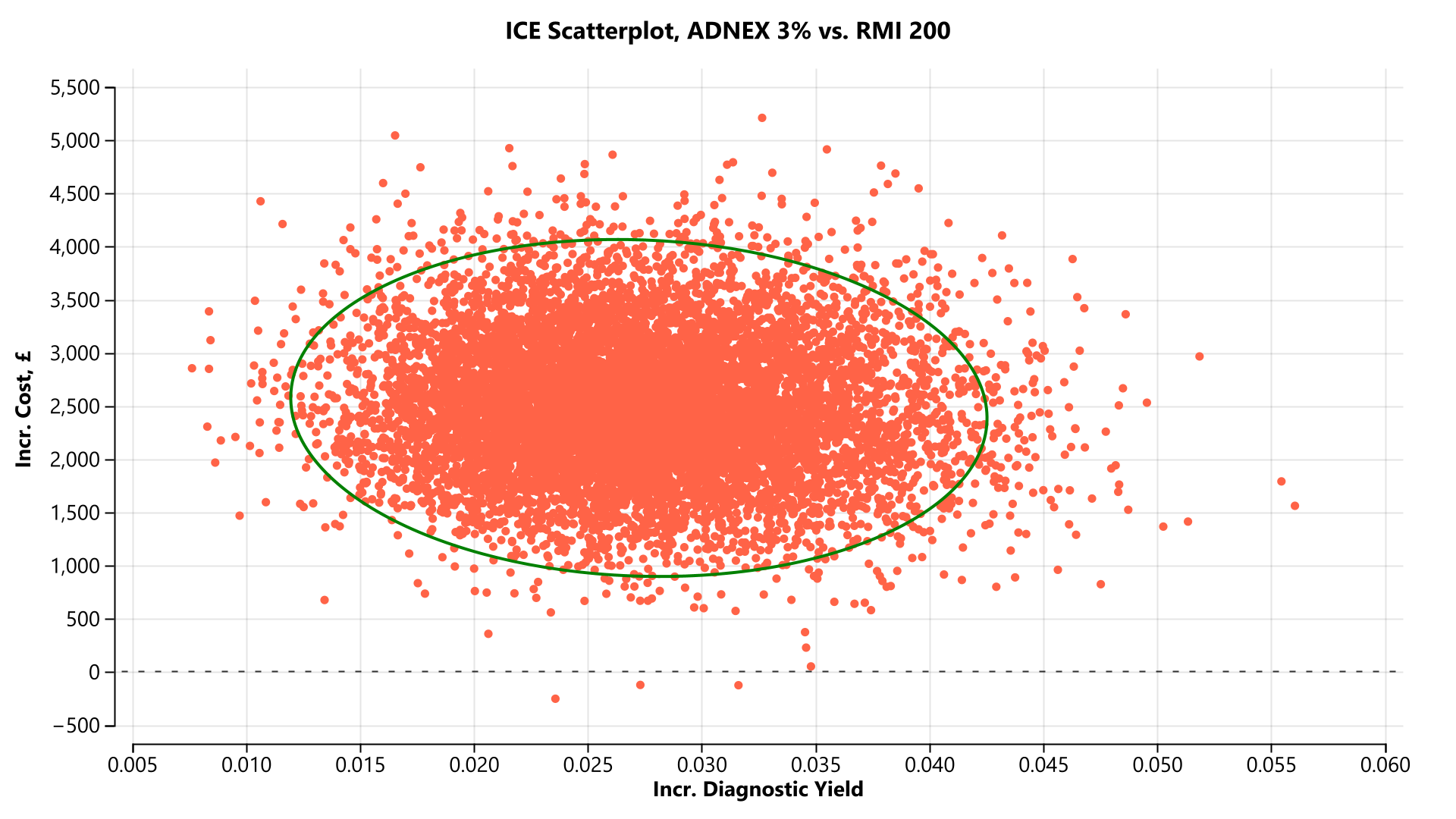
 **Supporting Information Figure S19 Pre-menopausal cohort incremental cost-effectiveness scatterplot (ADNEX 3% versus RMI 200) [diagnostic yield]**

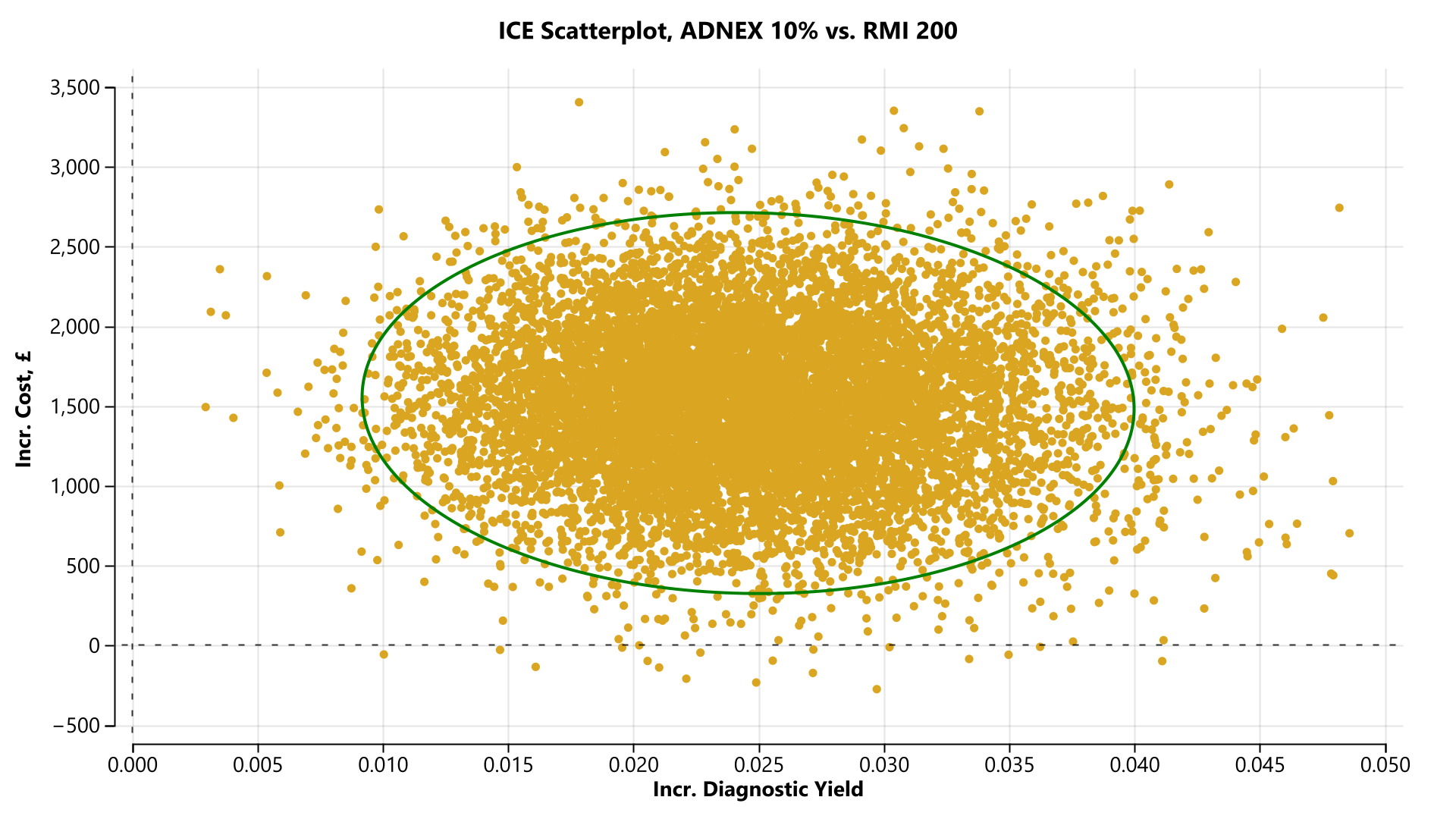
 **Supporting Information Figure S20 Pre-menopausal cohort incremental cost-effectiveness scatterplot (ADNEX 10% versus RMI 200) [diagnostic yield]**

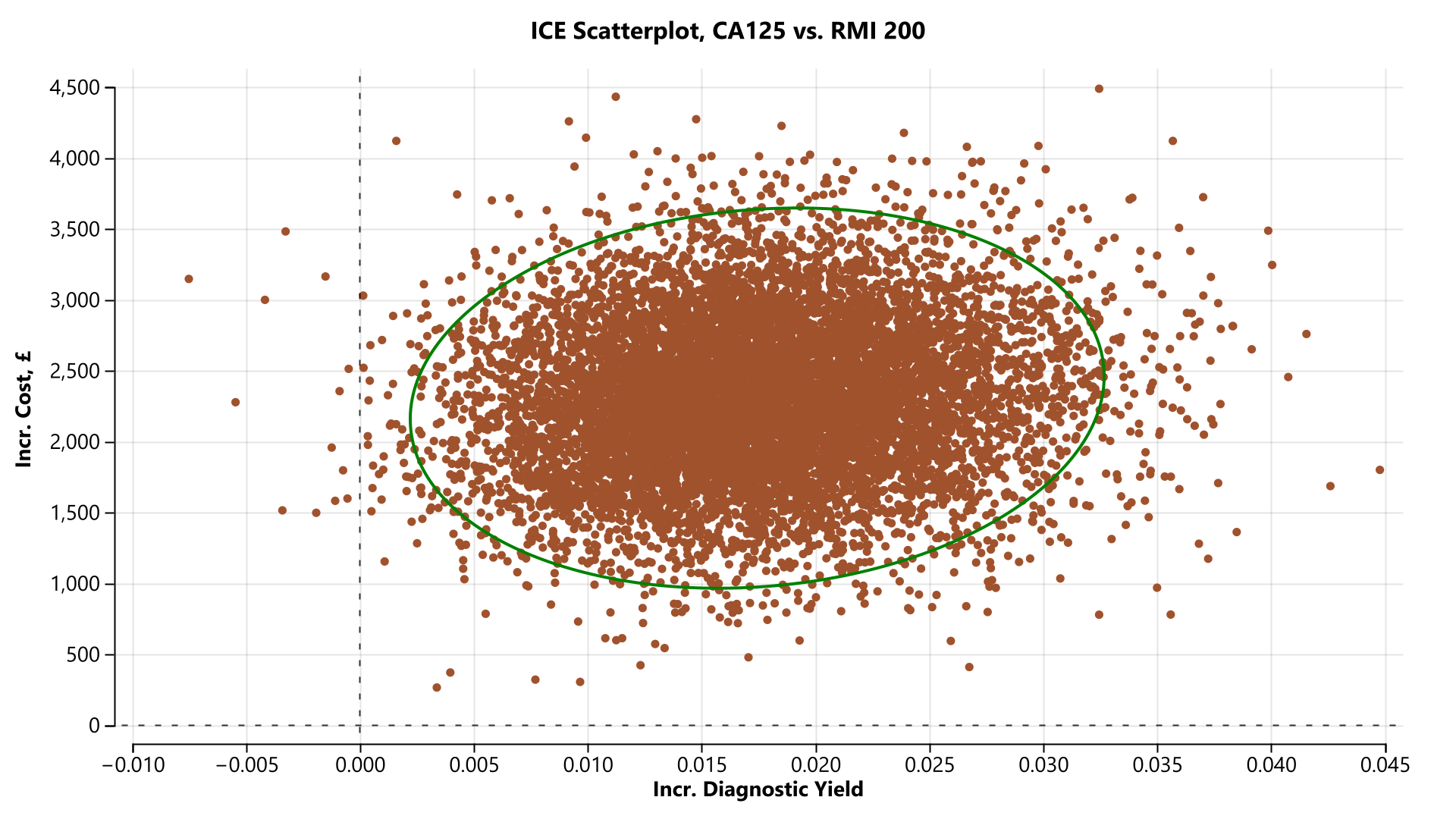
 **Supporting Information Figure S21 Pre-menopausal cohort incremental cost-effectiveness scatterplot (CA 125 versus RMI 200) [diagnostic yield]**

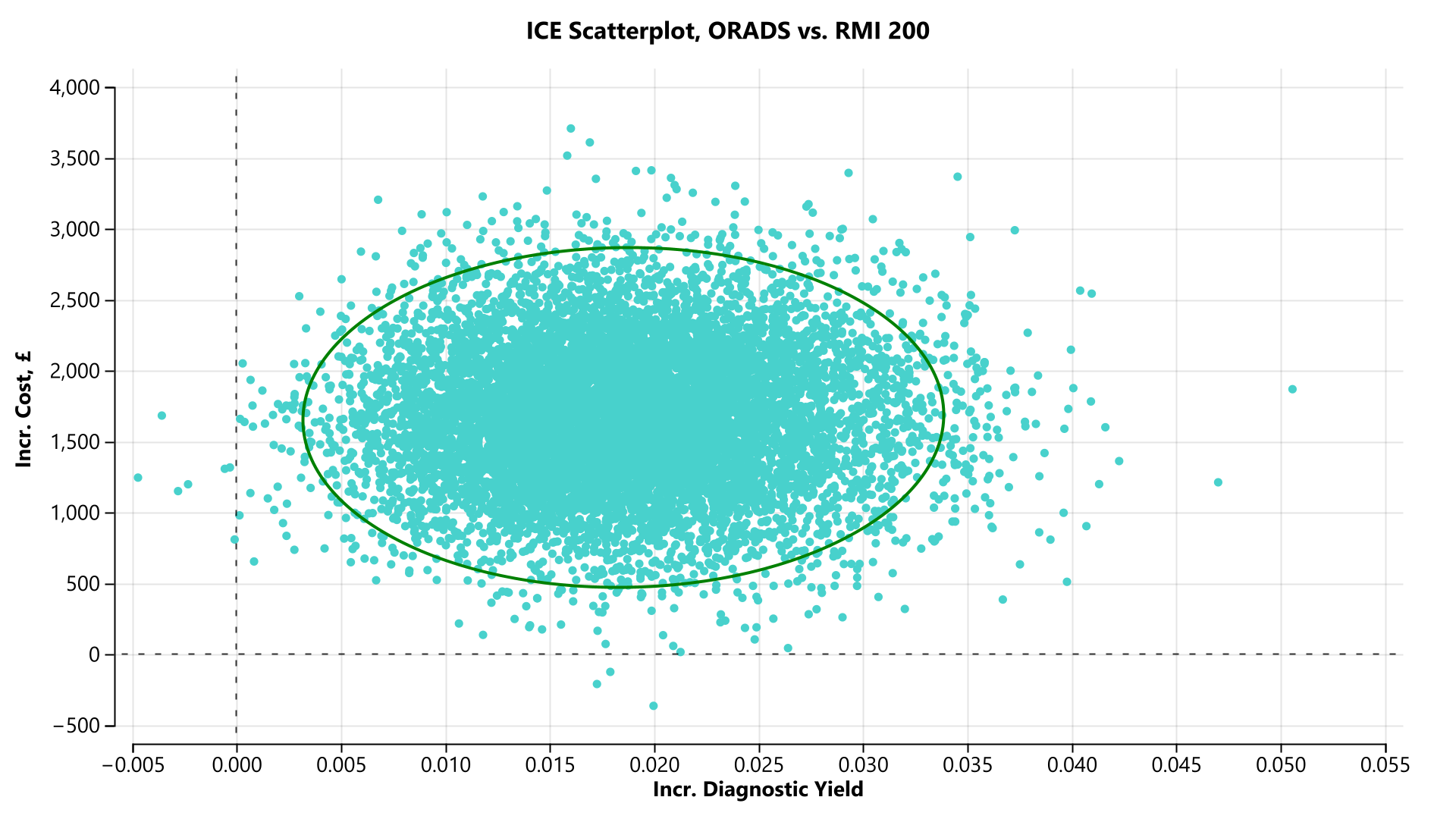

#### **Supporting Information Figure S22 Pre-menopausal cohort incremental cost-effectiveness scatterplot (ORADS versus RMI 200) [diagnostic yield]**

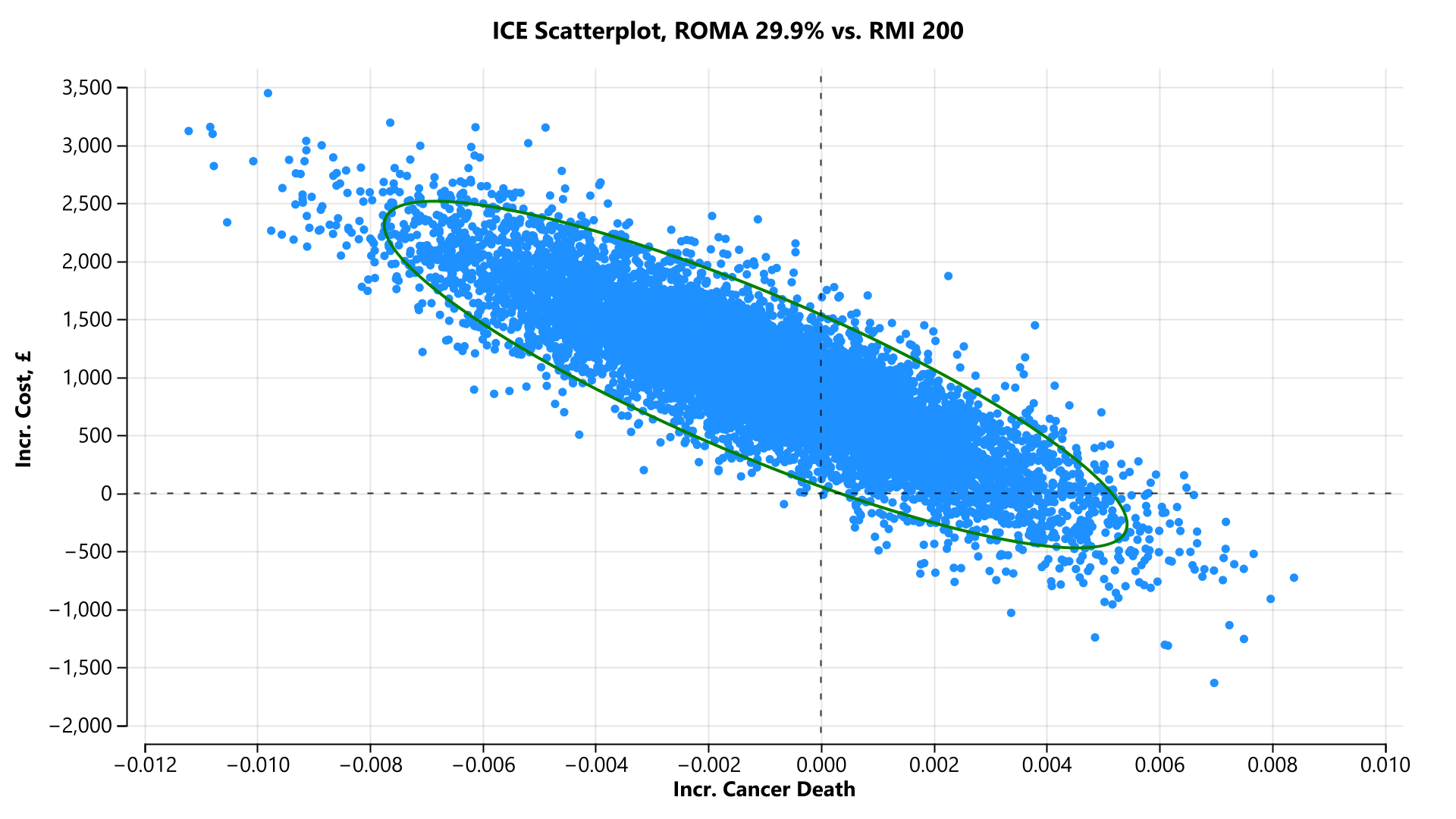

#### **Supporting Information Figure S23 Post-menopausal cohort incremental cost-effectiveness scatterplot (ROMA 29.9% versus RMI 200) [cancer death]**

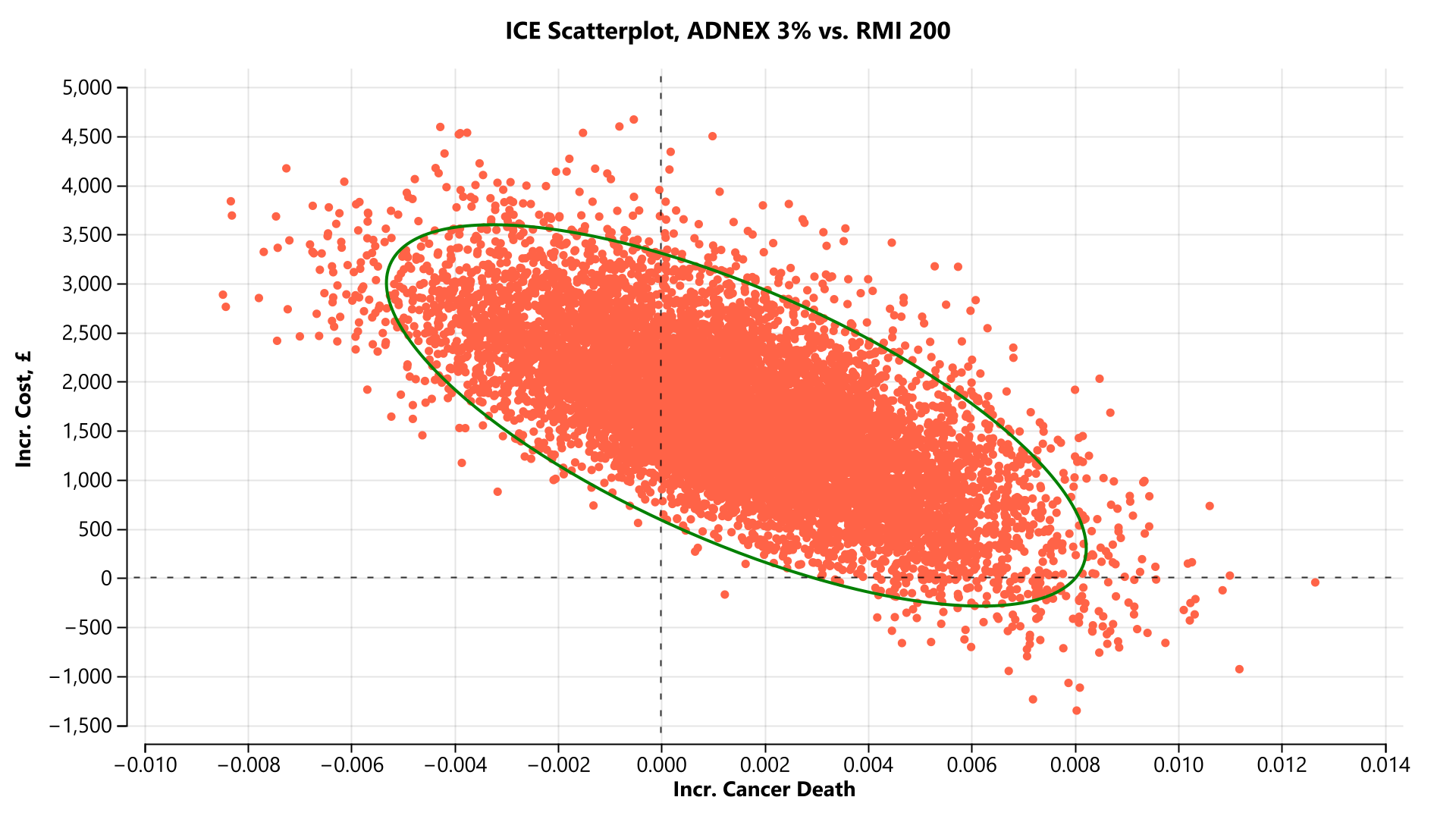

#### **Supporting Information Figure S24 Post-menopausal cohort incremental cost-effectiveness scatterplot (ADNEX 3% versus RMI 200) [cancer death]**

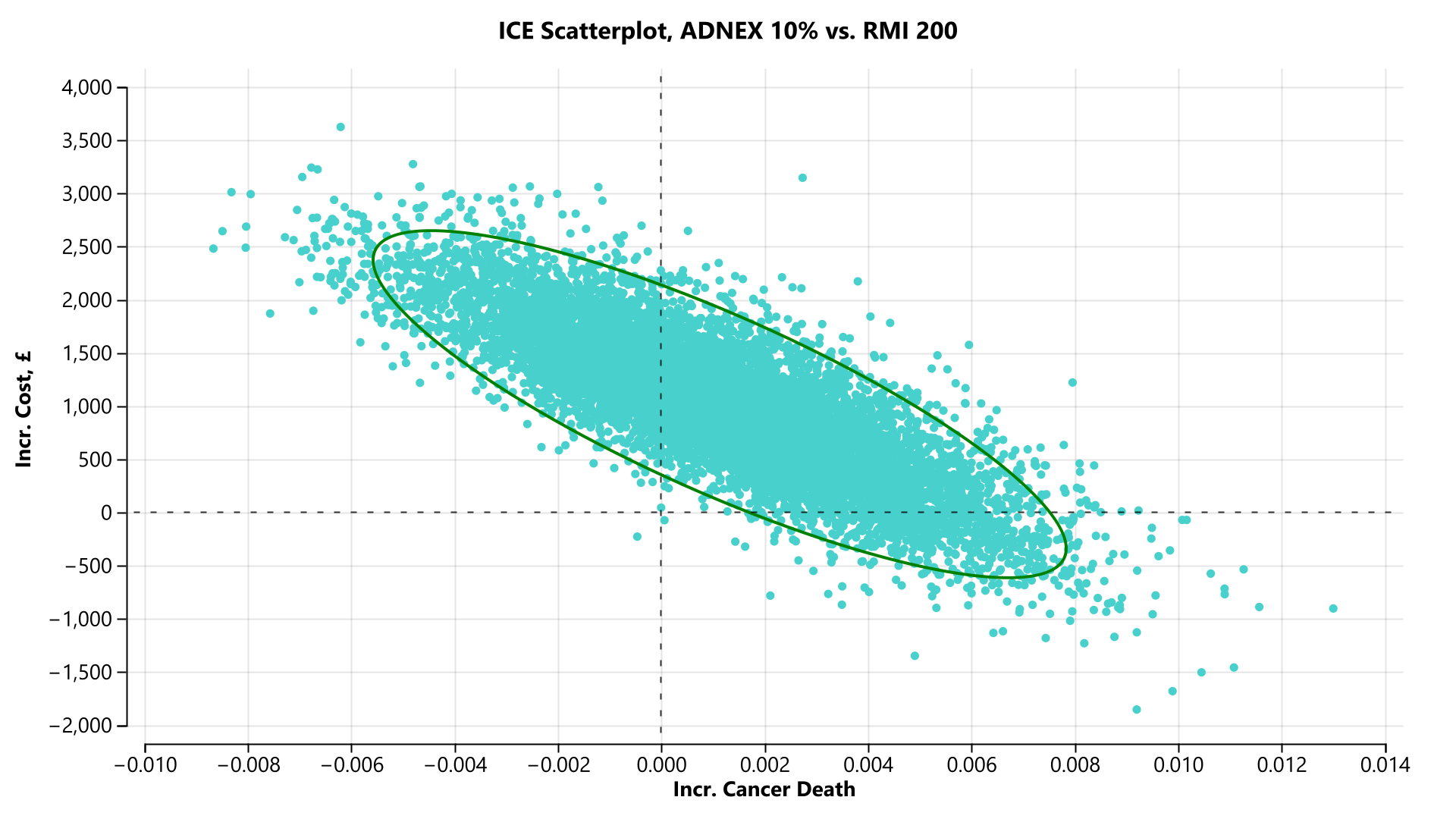

#### **Supporting Information Figure S25 Post-menopausal cohort incremental cost-effectiveness scatterplot (ADNEX 10% versus RMI 200) [cancer death]**

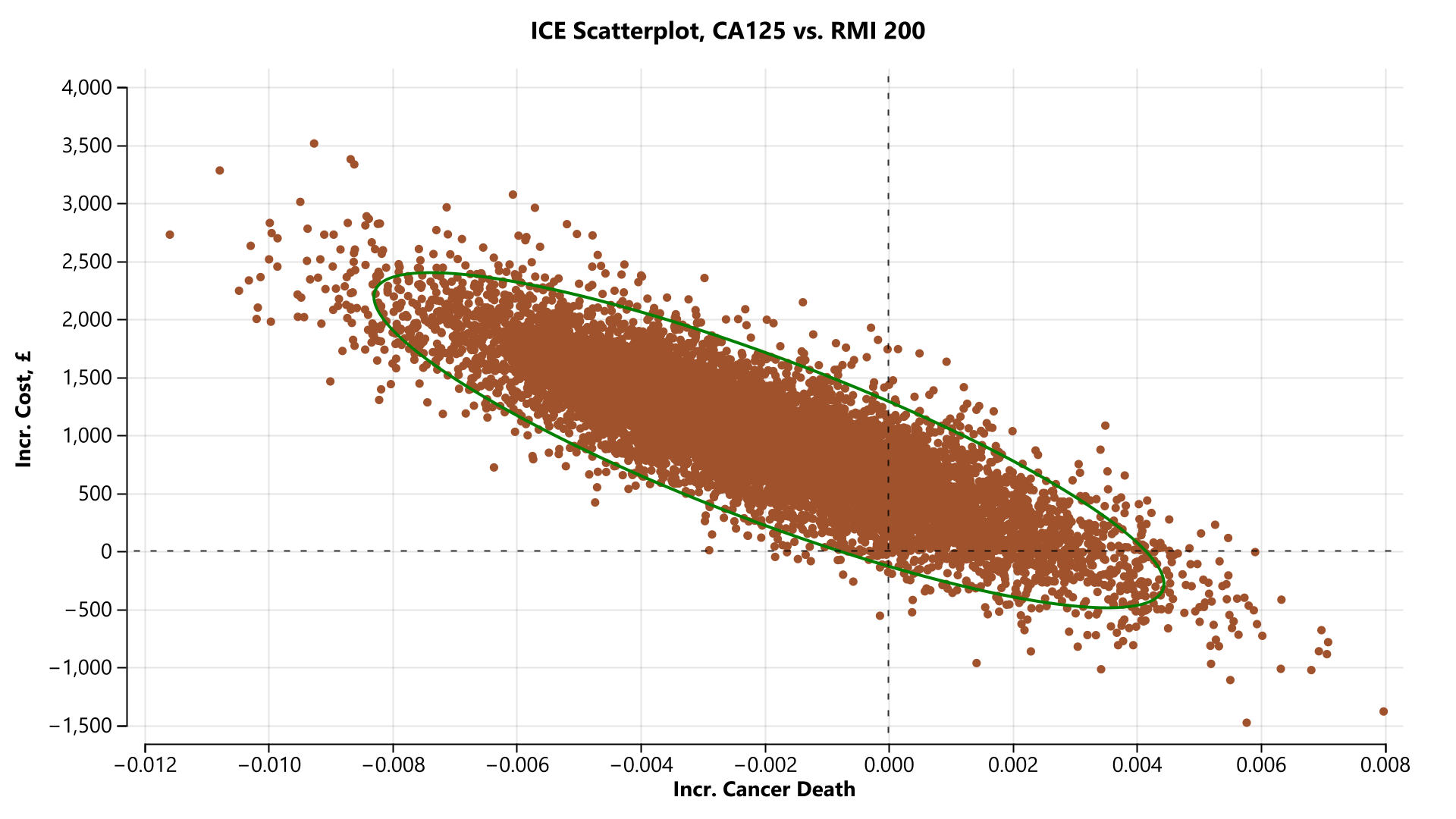

#### **Supporting Information Figure S26 Post-menopausal cohort incremental cost-effectiveness scatterplot (CA 125 versus RMI 200) [cancer death]**

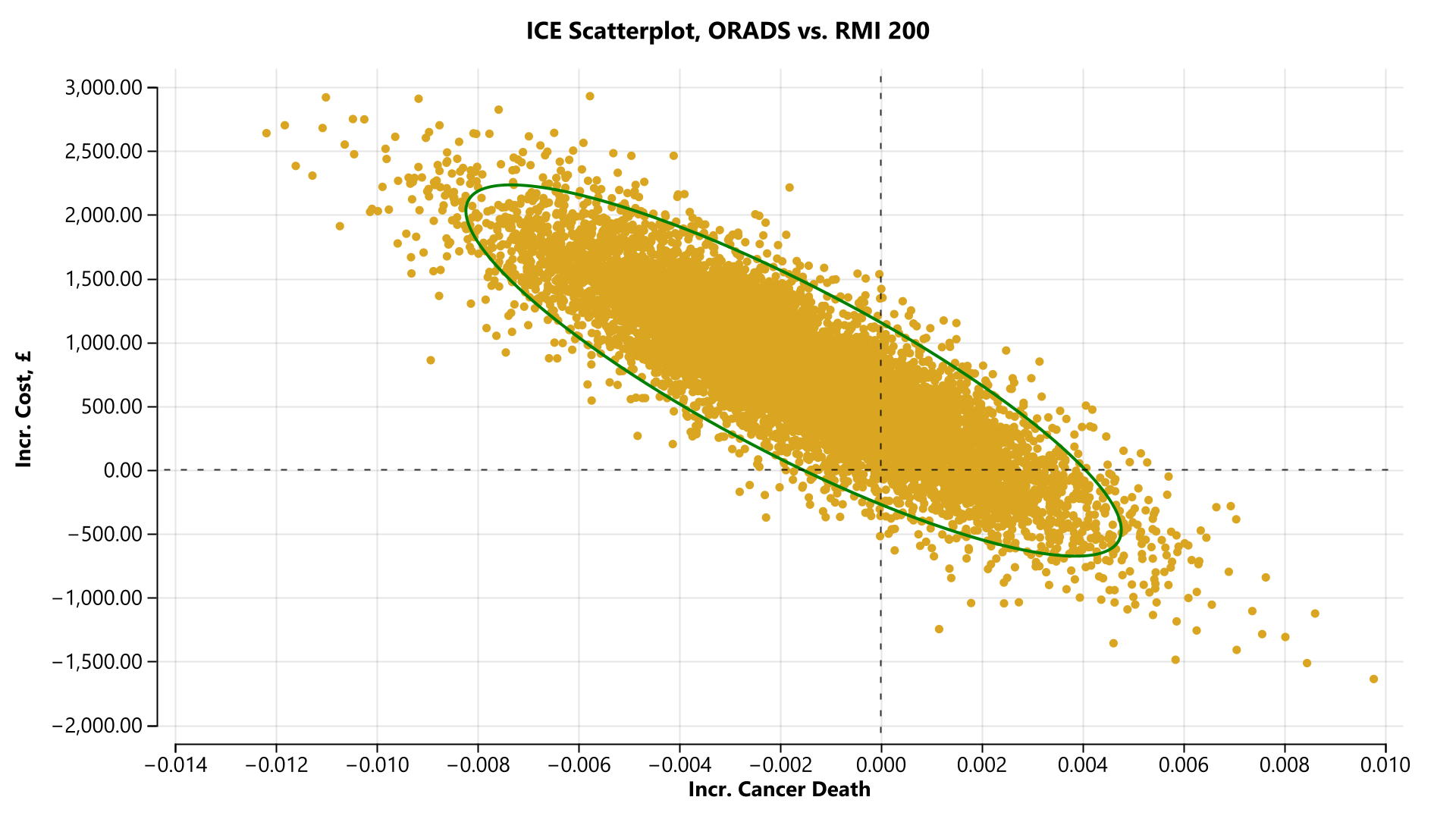

#### **Supporting Information Figure S27 Post-menopausal cohort incremental cost-effectiveness scatterplot (ORADS versus RMI 200) [cancer death]**

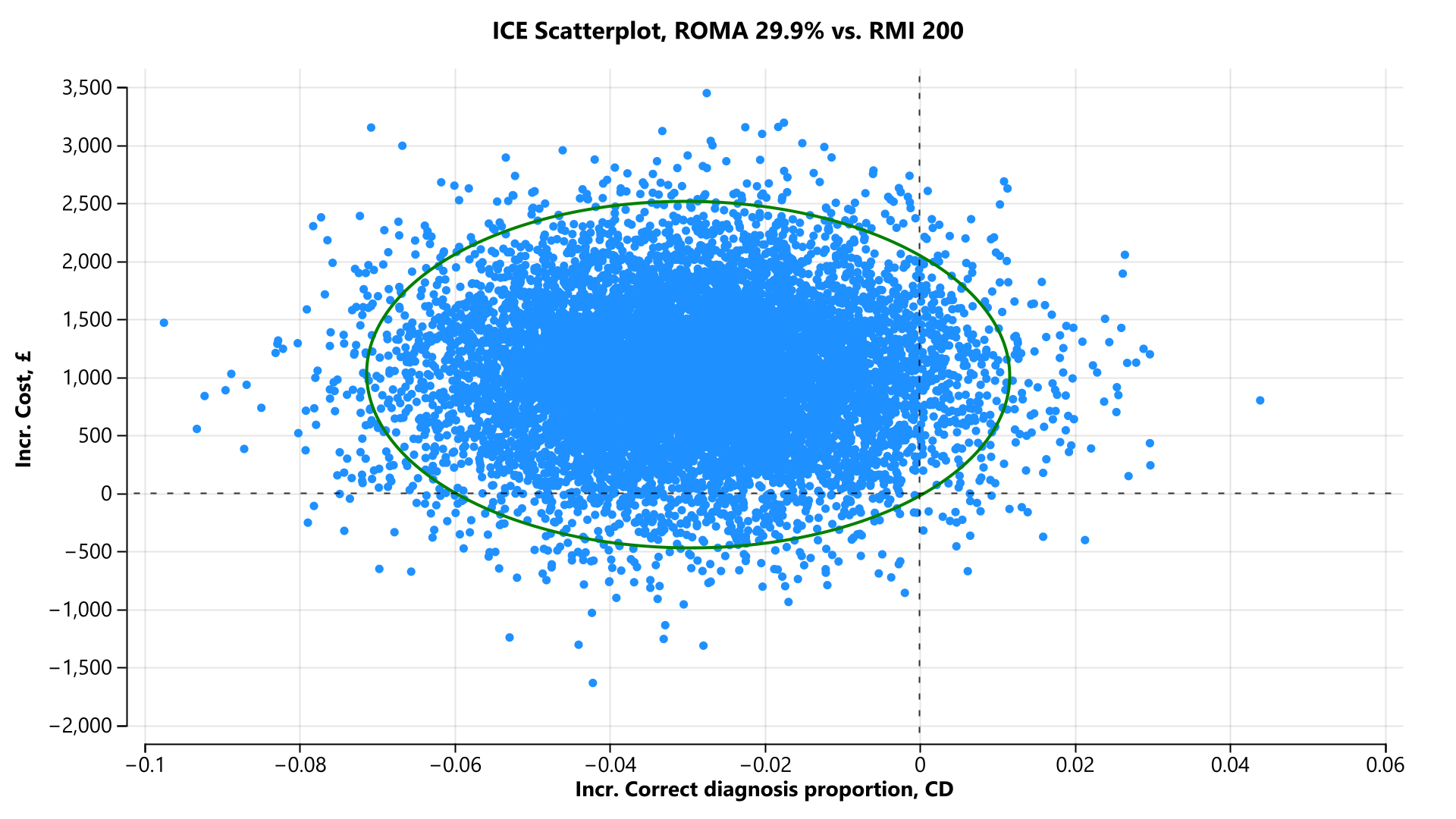

#### **Supporting Information Figure S28 Post-menopausal cohort incremental cost-effectiveness scatterplot (ROMA 29.9% versus RMI 200) [correct diagnosis]**

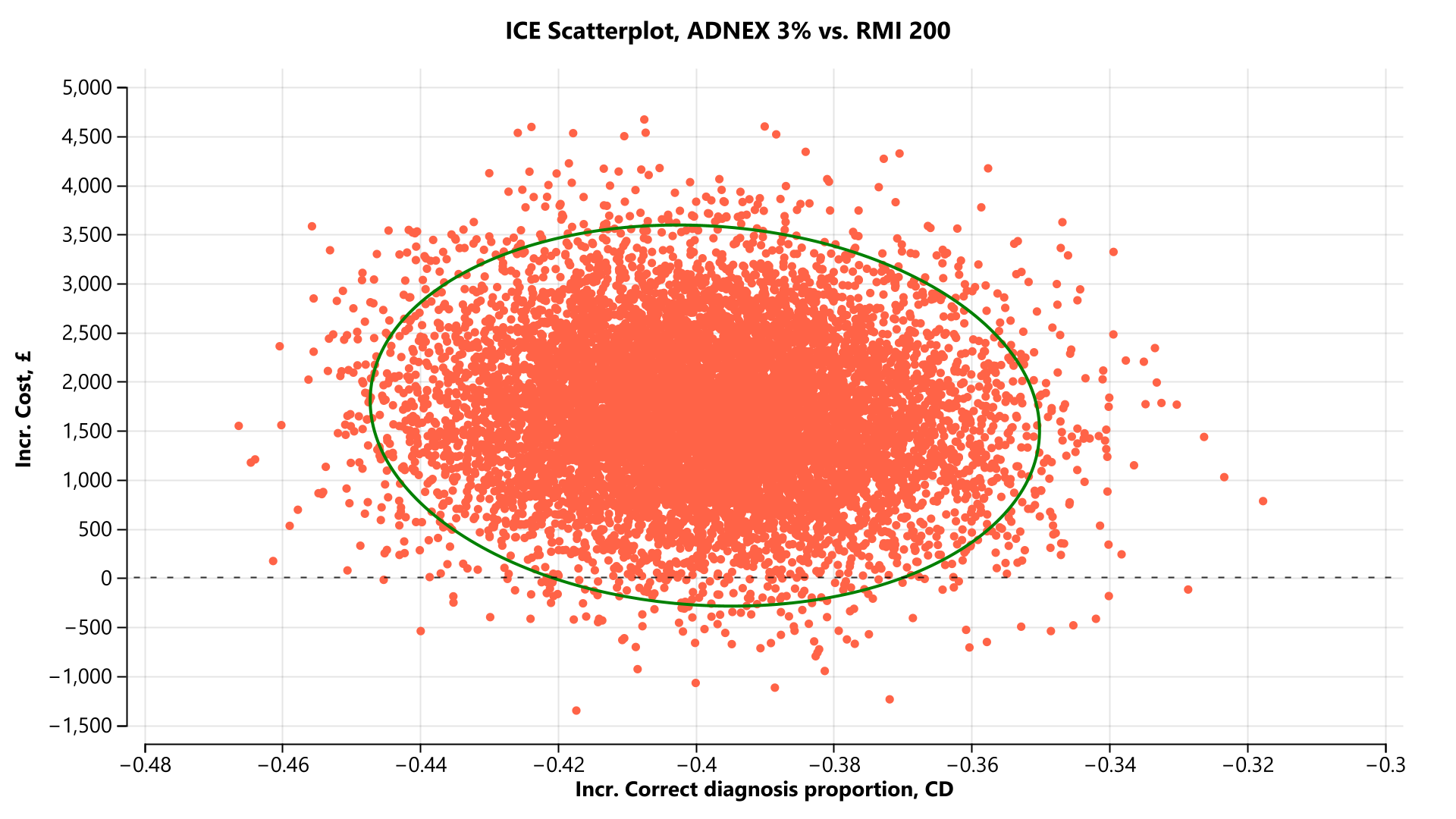

#### **Supporting Information Figure S29 Post-menopausal cohort incremental cost-effectiveness scatterplot (ADNEX 3% versus RMI 200) [correct diagnosis proportion]**

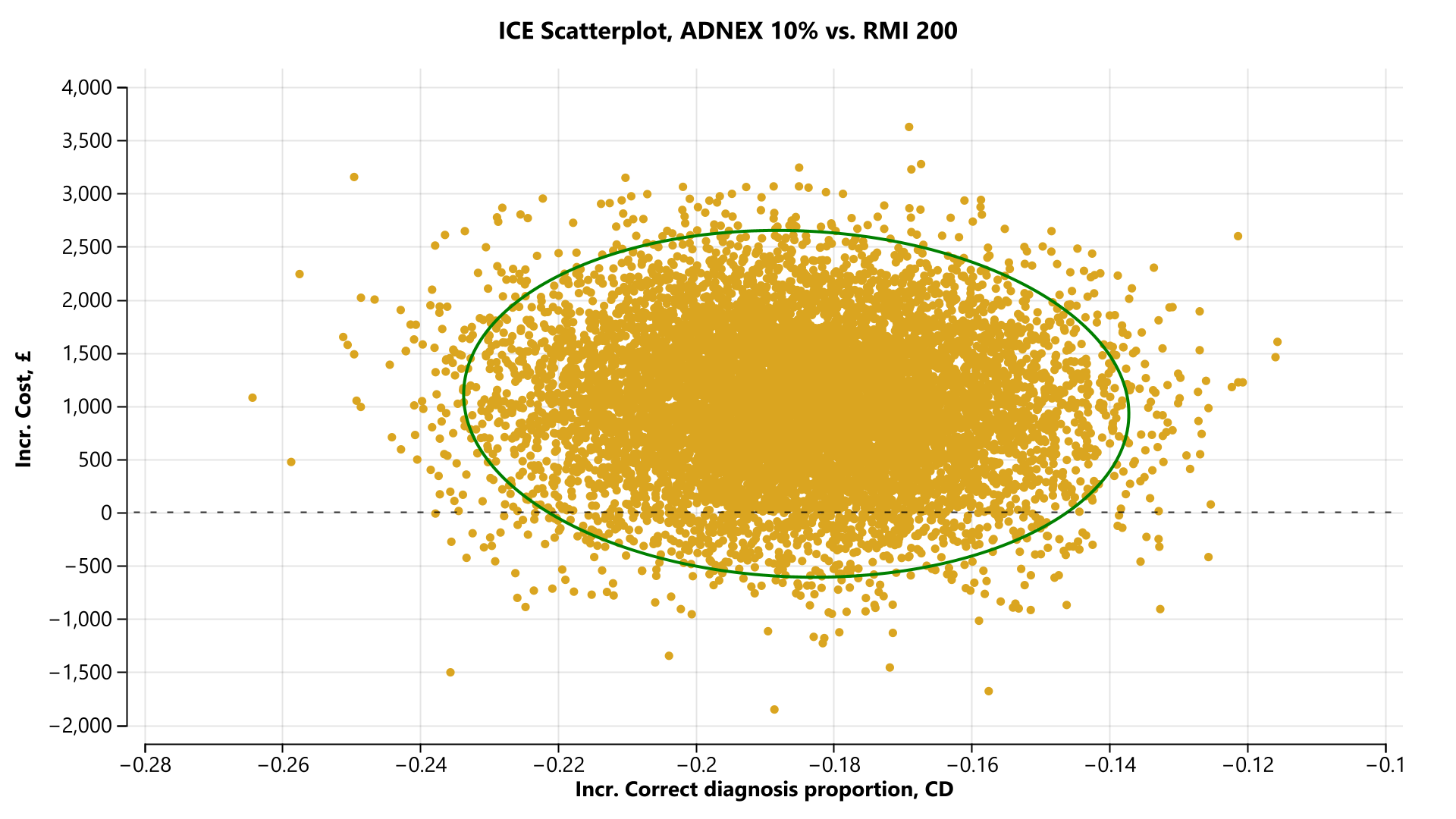

#### **Supporting Information Figure S30 Post-menopausal cohort incremental cost-effectiveness scatterplot (ADNEX 10% versus RMI 200) [correct diagnosis proportion]**

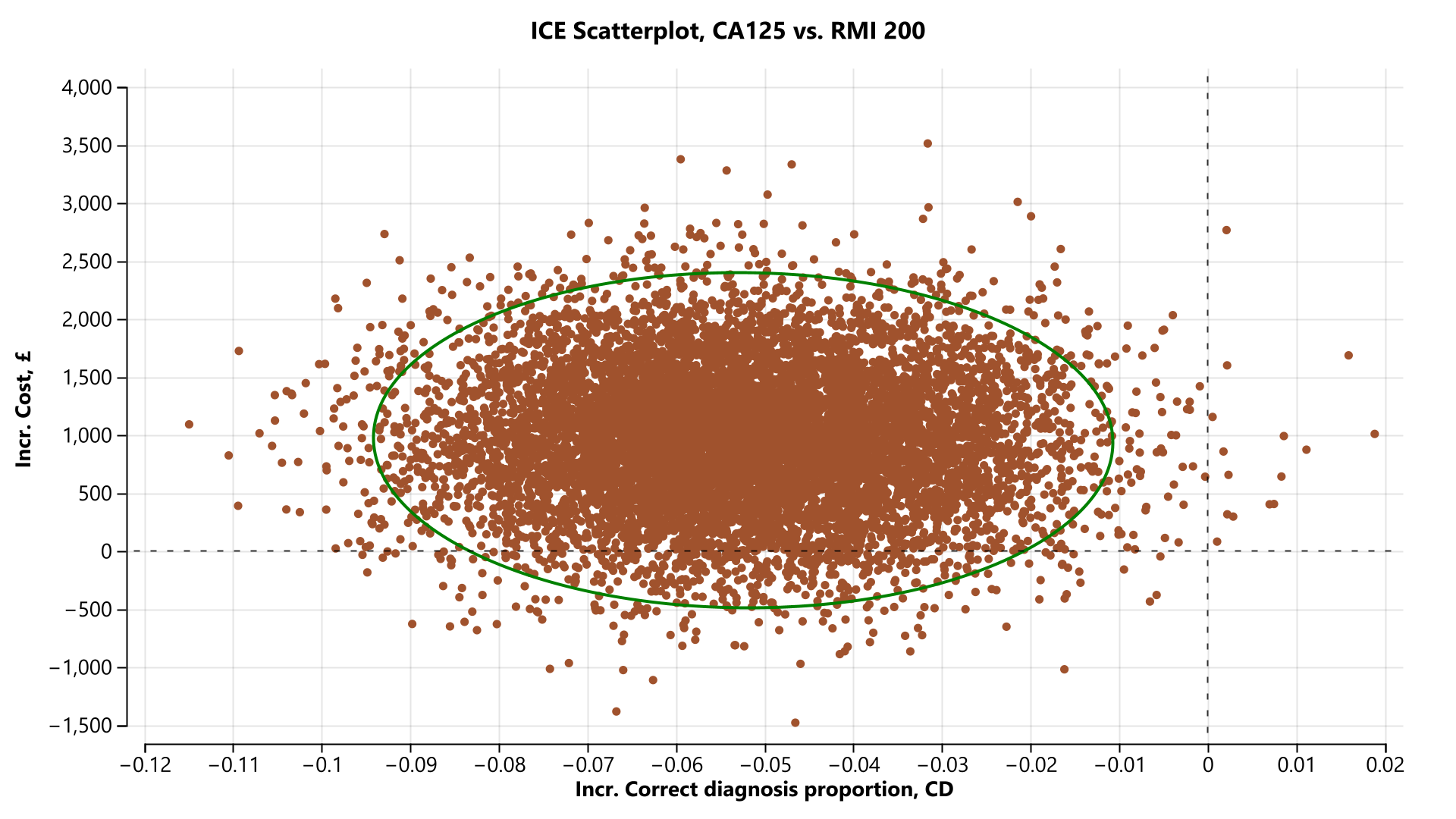

#### **Supporting Information Figure S31 Post-menopausal cohort incremental cost-effectiveness scatterplot (CA 125 versus RMI 200) [correct diagnosis proportion]**

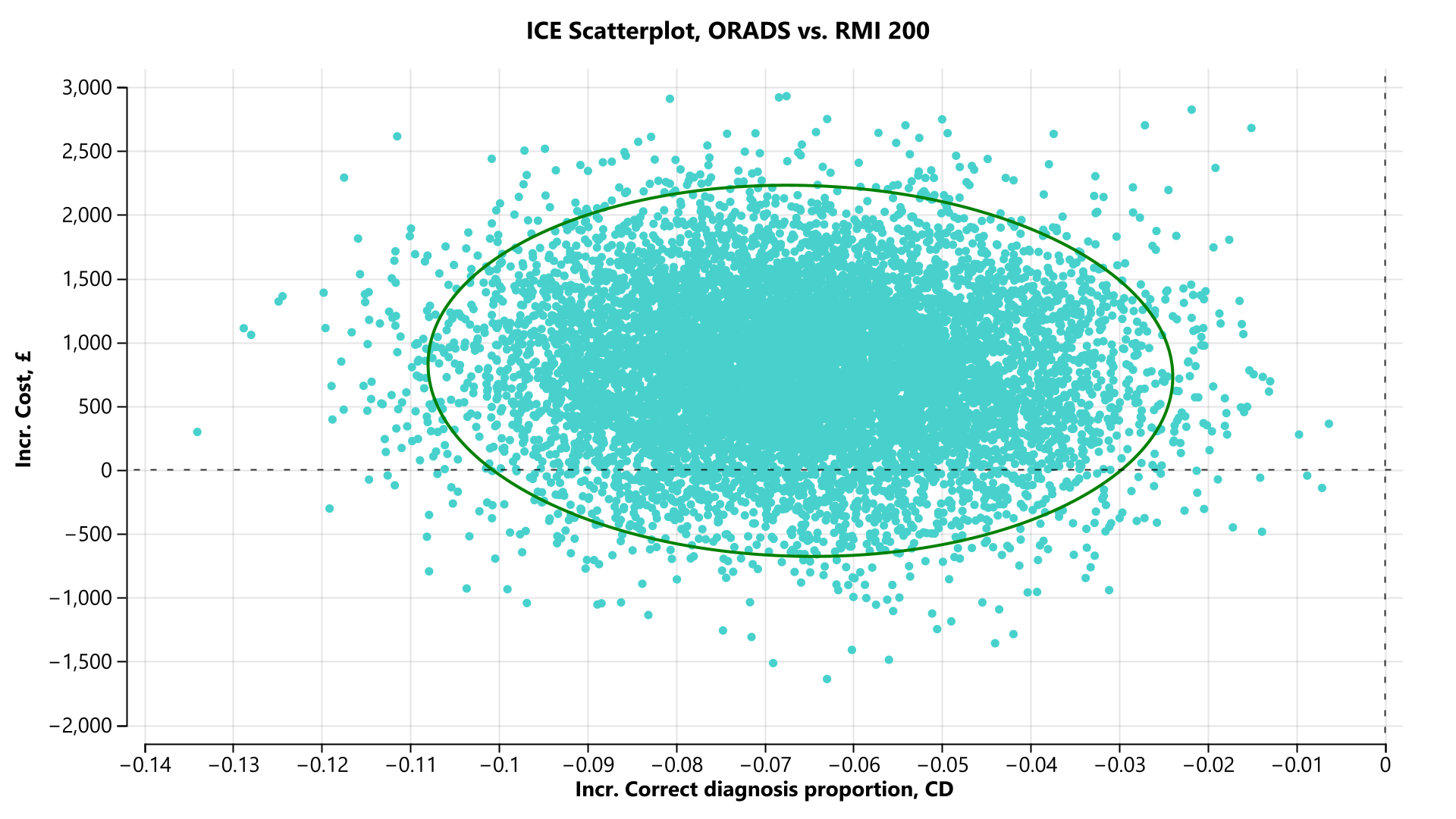

#### **Supporting Information Figure S32 Post-menopausal cohort incremental cost-effectiveness scatterplot (ORADS versus RMI 200) [correct diagnosis proportion]**

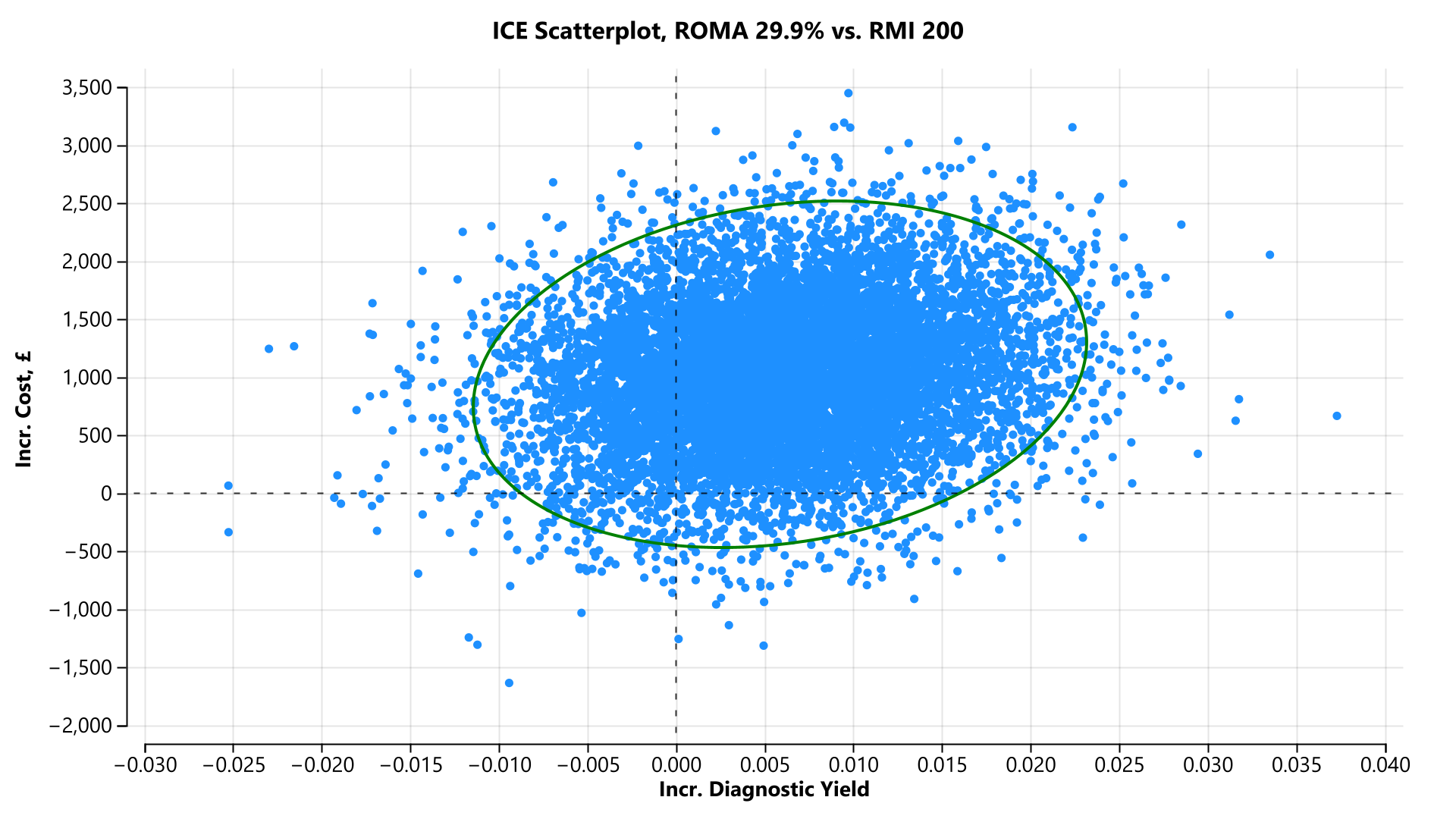

#### **Supporting Information Figure S33 Post-menopausal cohort incremental cost-effectiveness scatterplot (ROMA 29.9% versus RMI 200) [diagnostic yield]**

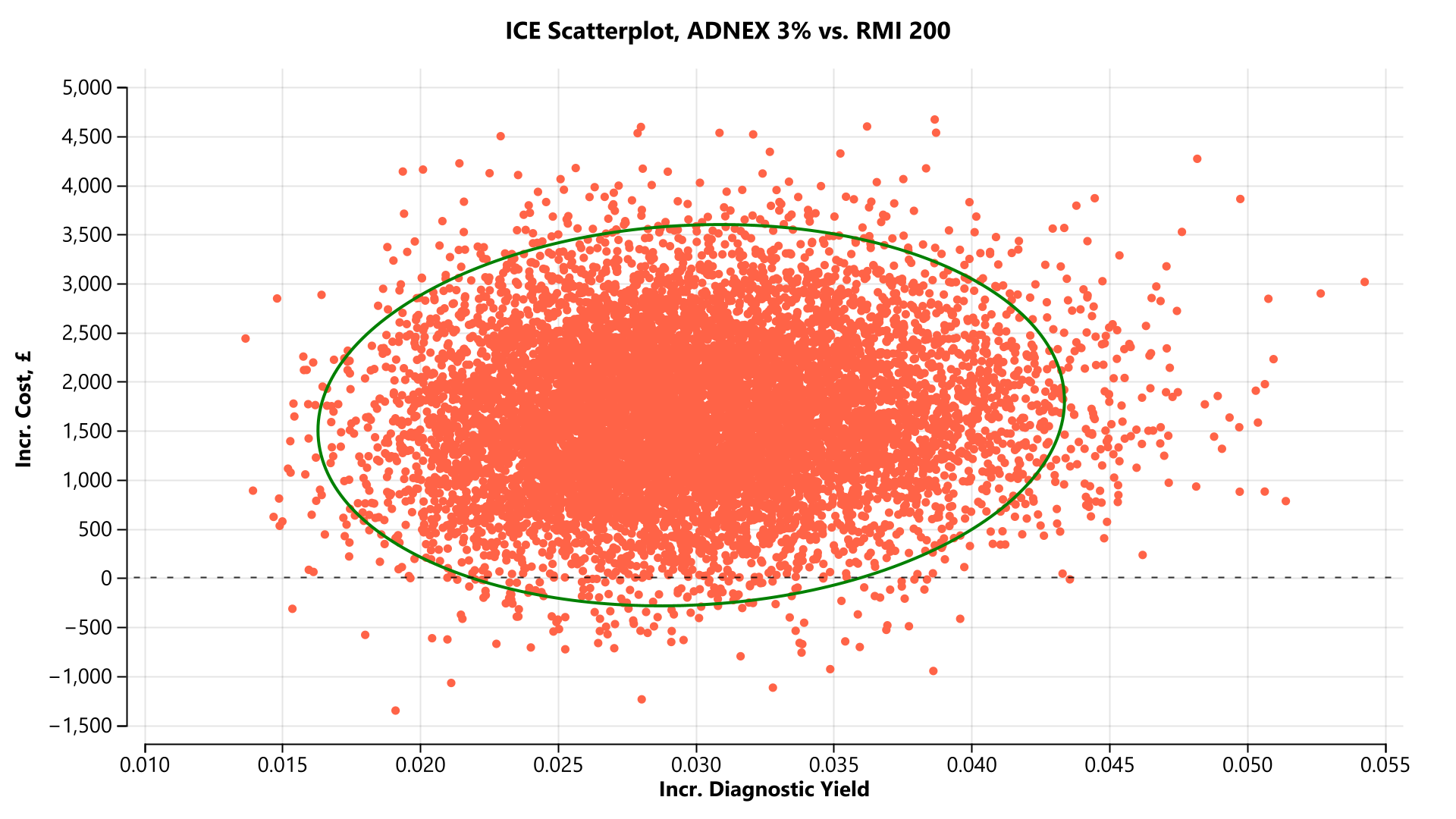

#### **Supporting Information Figure S34 Post-menopausal cohort incremental cost-effectiveness scatterplot (ADNEX 3% versus RMI 200) [diagnostic yield]**

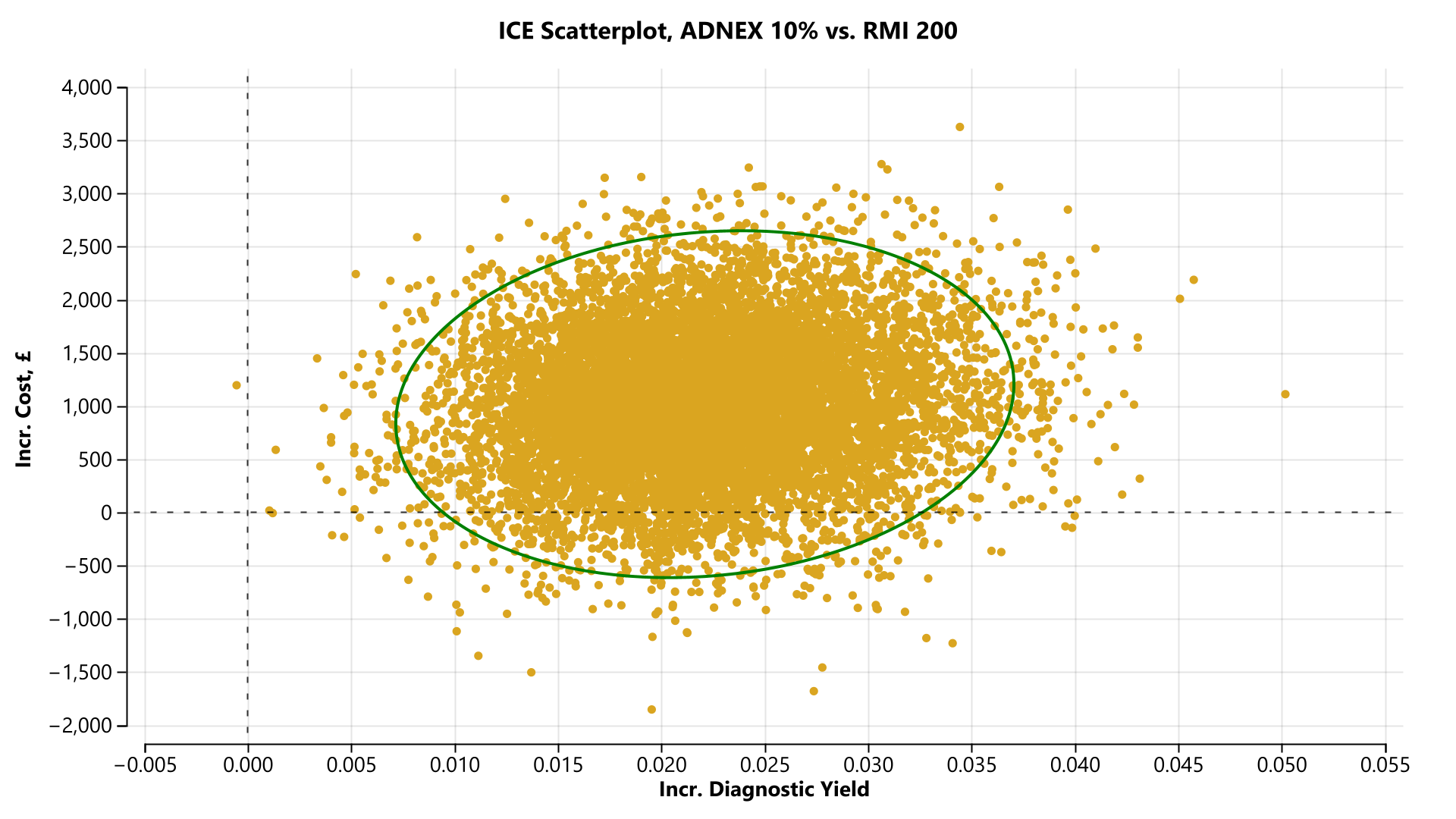
 **Supporting Information Figure S35 Post-menopausal cohort incremental cost-effectiveness scatterplot (ADNEX 10% versus RMI 200) [diagnostic yield]**

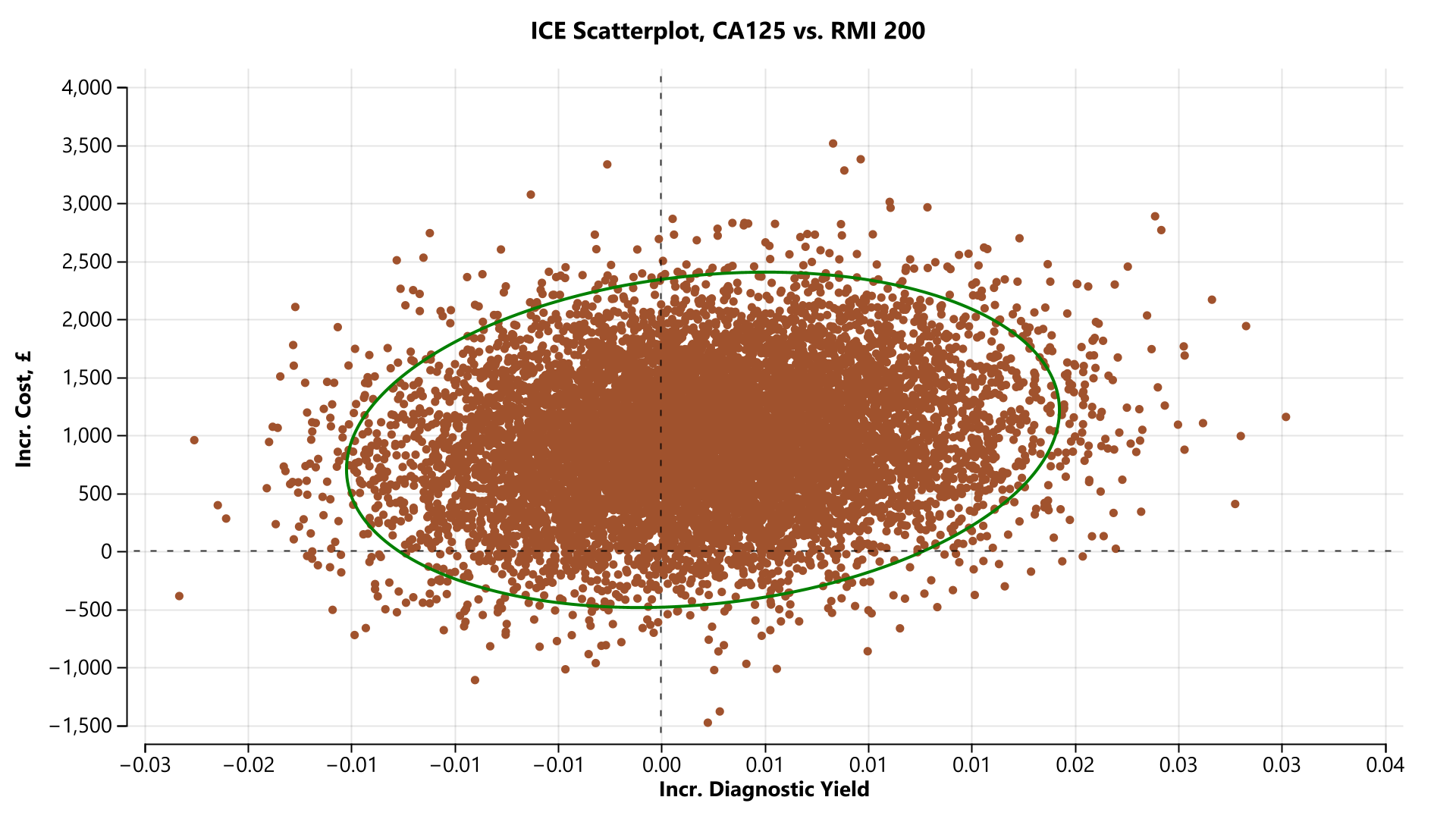
 **Supporting Information Figure S36 Post-menopausal cohort incremental cost-effectiveness scatterplot (CA 125 versus RMI 200) [diagnostic yield]**

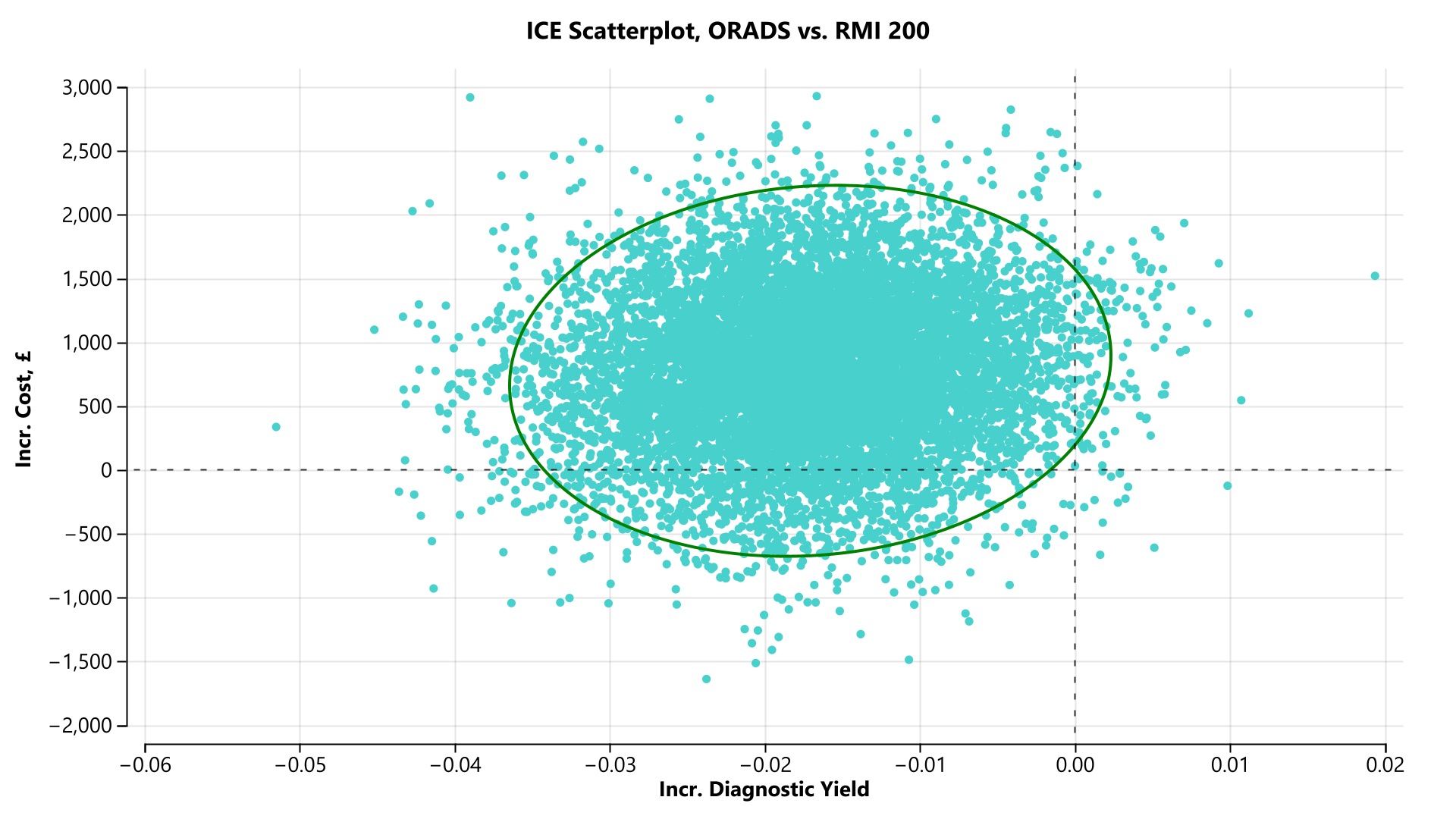

#### **Supporting Information Figure S37 Post-menopausal cohort incremental cost-effectiveness scatterplot (ORADS versus RMI 200) [diagnostic yield]**

**Table S6 CHEERS Checklist**

| **Topic** | **No.** | **Item** | **Location where item is reported** |
| --- | --- | --- | --- |
| **Title** |  |  |  |
|  | 1 | Identify the study as an economic evaluation and specify the interventions being compared. | Title, Page 1 |
| **Abstract** |  |  |  |
|  | 2 | Provide a structured summary that highlights context, key methods, results, and alternative analyses. | Abstract, Page 1 |
| **Introduction** |  |  |  |
| **Background and objectives** | 3 | Give the context for the study, the study question, and its practical relevance for decision making in policy or practice. | Introduction, Page 2 |
| **Methods** |  |  |  |
| **Health economic analysis plan** | 4 | Indicate whether a health economic analysis plan was developed and where available. | No HEAP - study was planned before this item was added to checklist |
| **Study population** | 5 | Describe characteristics of the study population (such as age range, demographics, socioeconomic, or clinical characteristics). | Methods, Page 3 |
| **Setting and location** | 6 | Provide relevant contextual information that may influence findings. | Methods, Page 3 |
| **Comparators** | 7 | Describe the interventions or strategies being compared and why chosen. | Methods, Page 4 |
| **Perspective** | 8 | State the perspective(s) adopted by the study and why chosen. | Methods, Page 4 |
| **Time horizon** | 9 | State the time horizon for the study and why appropriate. | Methods, Page 5 |
| **Discount rate** | 10 | Report the discount rate(s) and reason chosen. | Methods, Page 5 |
| **Selection of outcomes** | 11 | Describe what outcomes were used as the measure(s) of benefit(s) and harm(s). | Methods, Page 5 |
| **Measurement of outcomes** | 12 | Describe how outcomes used to capture benefit(s) and harm(s) were measured. | Methods, Page 5 |
| **Valuation of outcomes** | 13 | Describe the population and methods used to measure and value outcomes. | Methods, Page 5 |
| **Measurement and valuation of resources and costs** | 14 | Describe how costs were valued. | Methods, Page 5 |
| **Currency, price date, and conversion** | 15 | Report the dates of the estimated resource quantities and unit costs, plus the currency and year of conversion. | Methods, Page 5 |
| **Rationale and description of model** | 16 | If modelling is used, describe in detail and why used. Report if the model is publicly available and where it can be accessed. | Methods, Page 5 |
| **Analytics and assumptions** | 17 | Describe any methods for analysing or statistically transforming data, any extrapolation methods, and approaches for validating any model used. | Methods, Page 5 and Supporting Information |
| **Characterising heterogeneity** | 18 | Describe any methods used for estimating how the results of the study vary for subgroups. | Methods, Page 7 |
| **Characterising distributional effects** | 19 | Describe how impacts are distributed across different individuals or adjustments made to reflect priority populations. | Methods, Page 7 |
| **Characterising uncertainty** | 20 | Describe methods to characterise any sources of uncertainty in the analysis. | Methods, Page 7 |
| **Approach to engagement with patients and others affected by the study** | 21 | Describe any approaches to engage patients or service recipients, the general public, communities, or stakeholders (such as clinicians or payers) in the design of the study. | Not reported |
| **Results** |  |  |  |
| **Study parameters** | 22 | Report all analytic inputs (such as values, ranges, references) including uncertainty or distributional assumptions. | Results, Page 9 and Supporting Information |
| **Summary of main results** | 23 | Report the mean values for the main categories of costs and outcomes of interest and summarise them in the most appropriate overall measure. | Results, Page 8 |
| **Effect of uncertainty** | 24 | Describe how uncertainty about analytic judgments, inputs, or projections affect findings. Report the effect of choice of discount rate and time horizon, if applicable. | Results, Page 10 |
| **Effect of engagement with patients and others affected by the study** | 25 | Report on any difference patient/service recipient, general public, community, or stakeholder involvement made to the approach or findings of the study | Not reported |
| **Discussion** |  |  |  |
| **Study findings, limitations, generalisability, and current knowledge** | 26 | Report key findings, limitations, ethical or equity considerations not captured, and how these could affect patients, policy, or practice. | Discussion |
| **Other relevant information** |  |  |  |
| **Source of funding** | 27 | Describe how the study was funded and any role of the funder in the identification, design, conduct, and reporting of the analysis | End of manuscript |
| **Conflicts of interest** | 28 | Report authors conflicts of interest according to journal or International Committee of Medical Journal Editors requirements. | End of manuscript |

| ICMJE DISCLOSURE FORM | |
| --- | --- |
| **Date:** | 2/20/2025 |
| **Your Name:** | Samuel J. Perry |
| **Manuscript Title:** | A cost consequence analysis of six diagnostic strategies for ovarian cancer: A model-based economic evaluation |
| **Manuscript Number (if known):** | Click or tap here to enter text. |
| In the interest of transparency, we ask you to disclose all relationships/activities/interests listed below that are related to the content of your manuscript. “Related” means any relation with for-profit or not-for-profit third parties whose interests may be affected by the content of the manuscript. Disclosure represents a commitment to transparency and does not necessarily indicate a bias. If you are in doubt about whether to list a relationship/activity/interest, it is preferable that you do so.  The author’s relationships/activities/interests should be defined broadly. For example, if your manuscript pertains to the epidemiology of hypertension, you should declare all relationships with manufacturers of antihypertensive medication, even if that medication is not mentioned in the manuscript.  In item #1 below, report all support for the work reported in this manuscript without time limit. For all other items, the time frame for disclosure is the past 36 months. | |

|  | | | **Name all entities with whom you have this relationship or indicate none (add rows as needed)** | **Specifications/Comments (e.g., if payments were made to you or to your institution)** |
| --- | --- | --- | --- | --- |
| **Time frame: Since the initial planning of the work** | | | | |
| **1** | All support for the present manuscript (e.g., funding, provision of study materials, medical writing, article processing charges, etc.)  **No time limit for this item.** | | ☒ \| **None** \| Click the tab key to add additional rows. | |
| **Time frame: past 36 months** | | | | |
| **2** | | Grants or contracts from any entity (if not indicated in item #1 above). | ☒ \| **None** | |
| **3** | | Royalties or licenses | ☒ \| **None** | |
| **4** | | Consulting fees | ☒ \| **None** | |
| **5** | | Payment or honoraria for lectures, presentations, speakers bureaus, manuscript writing or educational events | ☒ \| **None** | |
| **6** | | Payment for expert testimony | ☐ \| **None** | |
| **7** | | Support for attending meetings and/or travel | ☐ \| **None** | |
| **8** | | Patents planned, issued or pending | ☒ \| **None** | |
| **9** | | Participation on a Data Safety Monitoring Board or Advisory Board | ☒ \| **None** | |
| **10** | | Leadership or fiduciary role in other board, society, committee or advocacy group, paid or unpaid | ☒ \| **None** | |
| **11** | | Stock or stock options | ☒ \| **None** | |
| **12** | | Receipt of equipment, materials, drugs, medical writing, gifts or other services | ☒ \| **None** | |
| **13** | | Other financial or non-financial interests | ☒ \| **None** | |
| **Please place an “X” next to the following statement to indicate your agreement:** | | | | |
| ☒ | | I certify that I have answered every question and have not altered the wording of any of the questions on this form. | | |

| ICMJE DISCLOSURE FORM \|; **Date:** \| 2/20/2025; **Your Name:** \| Donal Griffin; **Manuscript Title:** \| A cost consequence analysis of six diagnostic strategies for ovarian cancer: A model-based economic evaluation; **Manuscript Number (if known):** \| Click or tap here to enter text.; In the interest of transparency, we ask you to disclose all relationships/activities/interests listed below that are related to the content of your manuscript. “Related” means any relation with for-profit or not-for-profit third parties whose interests may be affected by the content of the manuscript. Disclosure represents a commitment to transparency and does not necessarily indicate a bias. If you are in doubt about whether to list a relationship/activity/interest, it is preferable that you do so. The author’s relationships/activities/interests should be defined broadly. For example, if your manuscript pertains to the epidemiology of hypertension, you should declare all relationships with manufacturers of antihypertensive medication, even if that medication is not mentioned in the manuscript. In item #1 below, report all support for the work reported in this manuscript without time limit. For all other items, the time frame for disclosure is the past 36 months. \| \| \| \| **Name all entities with whom you have this relationship or indicate none (add rows as needed)** \| **Specifications/Comments (e.g., if payments were made to you or to your institution)**; **Time frame: Since the initial planning of the work** \| \| \| \|; **1** \| All support for the present manuscript (e.g., funding, provision of study materials, medical writing, article processing charges, etc.) **No time limit for this item.** \| \| (☒ \| **None**) \| Click the tab key to add additional rows. \|; **Time frame: past 36 months** \| \| \| \|; **2** \| \| Grants or contracts from any entity (if not indicated in item #1 above). \| (☒ \| **None**) \|; **3** \| \| Royalties or licenses \| (☒ \| **None**) \|; **4** \| \| Consulting fees \| (☒ \| **None**) \|; **5** \| \| Payment or honoraria for lectures, presentations, speakers bureaus, manuscript writing or educational events \| (☒ \| **None**) \|; **6** \| \| Payment for expert testimony \| (☐ \| **None**) \|; **7** \| \| Support for attending meetings and/or travel \| (☐ \| **None**) \|; **8** \| \| Patents planned, issued or pending \| (☒ \| **None**) \|; **9** \| \| Participation on a Data Safety Monitoring Board or Advisory Board \| (☒ \| **None**) \|; **10** \| \| Leadership or fiduciary role in other board, society, committee or advocacy group, paid or unpaid \| (☒ \| **None**) \|; **11** \| \| Stock or stock options \| (☒ \| **None**) \|; **12** \| \| Receipt of equipment, materials, drugs, medical writing, gifts or other services \| (☒ \| **None**) \|; **13** \| \| Other financial or non-financial interests \| (☒ \| **None**) \|; **Please place an “X” next to the following statement to indicate your agreement:** \| \| \| \|; ☒ \| \| I certify that I have answered every question and have not altered the wording of any of the questions on this form. \| \| ICMJE DISCLOSURE FORM | |
| --- | --- |
| **Date:** | 2/20/2025 |
| **Your Name:** | Mark Monahan |
| **Manuscript Title:** | A cost consequence analysis of six diagnostic strategies for ovarian cancer: A model-based economic evaluation |
| **Manuscript Number (if known):** | Click or tap here to enter text. |
| In the interest of transparency, we ask you to disclose all relationships/activities/interests listed below that are related to the content of your manuscript. “Related” means any relation with for-profit or not-for-profit third parties whose interests may be affected by the content of the manuscript. Disclosure represents a commitment to transparency and does not necessarily indicate a bias. If you are in doubt about whether to list a relationship/activity/interest, it is preferable that you do so.  The author’s relationships/activities/interests should be defined broadly. For example, if your manuscript pertains to the epidemiology of hypertension, you should declare all relationships with manufacturers of antihypertensive medication, even if that medication is not mentioned in the manuscript.  In item #1 below, report all support for the work reported in this manuscript without time limit. For all other items, the time frame for disclosure is the past 36 months. | |

|  | | | **Name all entities with whom you have this relationship or indicate none (add rows as needed)** | **Specifications/Comments (e.g., if payments were made to you or to your institution)** |
| --- | --- | --- | --- | --- |
| **Time frame: Since the initial planning of the work** | | | | |
| **1** | All support for the present manuscript (e.g., funding, provision of study materials, medical writing, article processing charges, etc.)  **No time limit for this item.** | | ☒ \| **None** \| Click the tab key to add additional rows. | |
| **Time frame: past 36 months** | | | | |
| **2** | | Grants or contracts from any entity (if not indicated in item #1 above). | ☒ \| **None** | |
| **3** | | Royalties or licenses | ☒ \| **None** | |
| **4** | | Consulting fees | ☒ \| **None** | |
| **5** | | Payment or honoraria for lectures, presentations, speakers bureaus, manuscript writing or educational events | ☒ \| **None** | |
| **6** | | Payment for expert testimony | ☐ \| **None** | |
| **7** | | Support for attending meetings and/or travel | ☐ \| **None** | |
| **8** | | Patents planned, issued or pending | ☒ \| **None** | |
| **9** | | Participation on a Data Safety Monitoring Board or Advisory Board | ☒ \| **None** | |
| **10** | | Leadership or fiduciary role in other board, society, committee or advocacy group, paid or unpaid | ☒ \| **None** | |
| **11** | | Stock or stock options | ☒ \| **None** | |
| **12** | | Receipt of equipment, materials, drugs, medical writing, gifts or other services | ☒ \| **None** | |
| **13** | | Other financial or non-financial interests | ☒ \| **None** | |
| **Please place an “X” next to the following statement to indicate your agreement:** | | | | |
| ☒ | | I certify that I have answered every question and have not altered the wording of any of the questions on this form. | | |

| ICMJE DISCLOSURE FORM | |
| --- | --- |
| **Date:** | 2/20/2025 |
| **Your Name:** | Eleanor V. Williams |
| **Manuscript Title:** | A cost consequence analysis of six diagnostic strategies for ovarian cancer: A model-based economic evaluation |
| **Manuscript Number (if known):** | Click or tap here to enter text. |
| In the interest of transparency, we ask you to disclose all relationships/activities/interests listed below that are related to the content of your manuscript. “Related” means any relation with for-profit or not-for-profit third parties whose interests may be affected by the content of the manuscript. Disclosure represents a commitment to transparency and does not necessarily indicate a bias. If you are in doubt about whether to list a relationship/activity/interest, it is preferable that you do so.  The author’s relationships/activities/interests should be defined broadly. For example, if your manuscript pertains to the epidemiology of hypertension, you should declare all relationships with manufacturers of antihypertensive medication, even if that medication is not mentioned in the manuscript.  In item #1 below, report all support for the work reported in this manuscript without time limit. For all other items, the time frame for disclosure is the past 36 months. | |

|  | | | **Name all entities with whom you have this relationship or indicate none (add rows as needed)** | **Specifications/Comments (e.g., if payments were made to you or to your institution)** |
| --- | --- | --- | --- | --- |
| **Time frame: Since the initial planning of the work** | | | | |
| **1** | All support for the present manuscript (e.g., funding, provision of study materials, medical writing, article processing charges, etc.)  **No time limit for this item.** | | ☒ \| **None** \| Click the tab key to add additional rows. | |
| **Time frame: past 36 months** | | | | |
| **2** | | Grants or contracts from any entity (if not indicated in item #1 above). | ☒ \| **None** | |
| **3** | | Royalties or licenses | ☒ \| **None** | |
| **4** | | Consulting fees | ☒ \| **None** | |
| **5** | | Payment or honoraria for lectures, presentations, speakers bureaus, manuscript writing or educational events | ☒ \| **None** | |
| **6** | | Payment for expert testimony | ☐ \| **None** | |
| **7** | | Support for attending meetings and/or travel | ☐ \| **None** | |
| **8** | | Patents planned, issued or pending | ☒ \| **None** | |
| **9** | | Participation on a Data Safety Monitoring Board or Advisory Board | ☒ \| **None** | |
| **10** | | Leadership or fiduciary role in other board, society, committee or advocacy group, paid or unpaid | ☒ \| **None** | |
| **11** | | Stock or stock options | ☒ \| **None** | |
| **12** | | Receipt of equipment, materials, drugs, medical writing, gifts or other services | ☒ \| **None** | |
| **13** | | Other financial or non-financial interests | ☒ \| **None** | |
| **Please place an “X” next to the following statement to indicate your agreement:** | | | | |
| ☒ | | I certify that I have answered every question and have not altered the wording of any of the questions on this form. | | |
